# Using EEG to measure the neural effects of oxytocin administration: A meta-analysis and systematic review

**DOI:** 10.1101/2025.08.25.25334355

**Authors:** Elisabeth Deilhaug, Matthijs Moerkerke, Alina I. Sartorius, Heemin Kang, Emilie Smith-Meyer Kildal, Kjersti M. Walle, Torbjørn Elvsåshagen, Lars T. Westlye, Terje Nærland, Ole A. Andreassen, Daniel S. Quintana

**Affiliations:** NevSom, Department of Rare Disorders, Oslo University Hospital, Oslo, Norway; KG Jebsen Centre for Neurodevelopmental Disorders, University of Oslo, Oslo, Norway; Institute of Clinical Medicine, Faculty of Medicine, University of Oslo, Oslo, Norway; Department of Neurosciences, Center for Developmental Psychiatry, KU Leuven, Leuven, Belgium; Leuven Autism Research (LAuRes), KU Leuven, Leuven, Belgium; Department of Psychology, University of Oslo, Oslo, Norway; Centre for Precision Psychiatry, Division of Mental Health and Addiction, University of Oslo and Oslo University Hospital, Oslo, Norway; Department of Neurology, Division of Clinical Neuroscience, Oslo University Hospital, Oslo, Norway; Department of Behavioural Medicine, Institute of Basic Medical Sciences, University of Oslo, Oslo, Norway

**Keywords:** oxytocin, meta-analysis, systematic review, EEG, electroencephalography, neuroimaging

## Abstract

Electroencephalography (EEG) has emerged as a key method for investigating the neural mechanisms through which oxytocin influences cognition and behaviour. EEG is cost-effective, has excellent temporal precision, and may elucidate neural correlates of emotional and cognitive processes. EEG studies evaluating oxytocin’s electrophysiological effects have, however, yielded mixed results, which is likely driven by heterogeneity in EEG measures, study designs, dosages, and samples. To investigate the effect of oxytocin administration on EEG measures, we performed two multilevel random effects meta-analyses: The first meta-analysis synthesized studies investigating the effects of oxytocin administration on different neural correlates of social and cognitive processing; the second meta-analysis synthesized studies evaluating effects of oxytocin administration on exploratory, less task-specific neural activity measures, such as the modulation of microstates. Across both meta-analyses, we synthesized 161 effect sizes from 28 randomised controlled trials with a total of 1361 participants from different population groups. These multilevel meta-analyses yielded small effect sizes of oxytocin administration across different EEG measures reflecting social and cognitive processes (Hedges *g* = 0.14), and exploratory neural activity (Hedges *g* = 0.28) with significant heterogeneity estimates (*p* < 0.01 and *p* < 0.001, respectively). Moderator analyses revealed that the different EEG measurements of interest (e.g., event-related potentials) and the proportion of female participants were found to significantly moderate the effect of oxytocin on neural EEG activity. Altogether, these meta-analyses present tentative evidence for oxytocin administration modulating a wide range of neural activity. We observed substantial heterogeneity across studies - in terms of study designs, experimental paradigms and EEG measurements, and participant characteristics. More research is warranted to map out the context-specific effects of oxytocin administration on different neural markers, to better understand the neurobiological mechanisms of oxytocin.

## Introduction

Oxytocin is a hormone and neuromodulator that is predominantly produced in the supraoptic and paraventricular nuclei of the hypothalamus and is well known for its effects on lactation and childbirth (Gimpl & Fahrenholz, 2001; Jurek & Neumann, 2018). Oxytocin is released as a hormone into the peripheral blood stream but also acts as a neuromodulator locally in the brain. Since Kosfeld and colleagues’ (2005, but also see Declerck et al., 2020) reported that oxytocin may promote trust in humans, there has been a steady increase in oxytocin research on psychosocial functioning in humans (Jurek & Neumann, 2018; Leng & Leng, 2021). For example, studies have reported that oxytocin administration increases motivation to cooperate (Daughters et al., 2017; Lambert et al., 2017; Ten Velden et al., 2017), modulates the perception of social cues (Quintana et al., 2015) and increases empathetic accuracy (Bartz et al., 2010).

Moreover, oxytocin has also been reported to support social behaviour in some populations with psychiatric illnesses and neurodevelopmental differences (Anagnostou et al., 2012; Andari et al., 2010; Audunsdottir et al., 2024; Auyeung et al., 2015; Cochran et al., 2013; Guastella et al., 2010, 2015; Hollander et al., 2007; Ma et al., 2016; Rubin et al., 2014).

To complement the focus on social outcomes, emerging evidence highlights oxytocin’s role in plasticity and cognitive processes, pointing to broader mechanisms of behavioural regulation (Grace et al., 2018; Kang et al., 2025; Quintana et al., 2016, 2021; Wigton et al., 2015). Oxytocin receptors are present in all brain regions, particularly in the amygdala, hypothalamus, hippocampus and prefrontal cortex (Grinevich et al., 2015; Jurek & Neumann, 2018; Mitre et al., 2018), and are released from axon terminals, dendrites and soma of both pre-and post-synaptic neurons (Bakos et al., 2018; Mitre et al., 2018; van den Pol, 2012). Exogenous oxytocin has been linked to enhancing synaptic plasticity in the visual cortex, via modulations of astrocyte activity and an increase in excitatory synaptic protein expression (Sun et al., 2025), and recent research indicates that oxytocin has beneficial effects for neural processes that facilitate learning. Animal studies suggest that exogenous oxytocin may induce cortical plasticity in the auditory cortex by “turning up the volume” of social stimuli relevant for species survival, specifically by promoting maternal behaviour in response to pup calls for female mice (Carcea et al., 2021; Marlin et al., 2015).Recently, oxytocin administration has been found to increase motivational salience towards stimuli of personal relevance in humans, where neural and behavioural approach-avoidance of *both* social and non-social stimuli was enhanced (Alaerts et al., 2021). Additionally, oxytocin administration has been suggested to improve rats’ performance on non-social probabilistic reversal learning tasks (Roberts et al., 2019), and recent research in humans reports similar findings, such as a reduction in rigid behaviour in samples of autistic (Hollander et al., 2007) and anorexic participants (Russell et al., 2018). Oxytocin has also been shown to provide beneficial effects on working memory in populations with Huntington’s disease (Fisher et al., 2021) and schizophrenia (Michalopoulou et al., 2015). Findings of altered responses to both social and non-social stimuli could support the proposed allostatic theory of oxytocin, where oxytocin is thought to maintain stability by anticipating future changes in the environment and facilitate adaptive responses to these changes (Quintana & Guastella, 2020). Such findings highlight oxytocin’s potential role in promoting behavioural flexibility across a range of contexts.

Gaining a better understanding of the neurobiological mechanisms underlying oxytocin’s effects on cognition and behaviour is crucial for developing therapeutic interventions targeting oxytocin signalling in the brain, which may hold potential for alleviating social difficulties, anxiety disorders, and characteristics affiliated with various neuropsychiatric conditions (Quintana et al., 2021; Sippel et al., 2017; Weisman & Feldman, 2013). The number of studies using non-invasive neuroimaging techniques to better understand oxytocin’s neurobiological effects has increased exponentially over the past two decades (Grace et al., 2018; Leng & Leng, 2021; Quintana et al., 2016). With its high spatial resolution, functional magnetic resonance imaging (fMRI) is an often-utilised tool to measure neural activity. However, it does not reflect specific neuromodulatory mechanisms nor measure neuronal firing and has low temporal resolution (Ekstrom, 2010; Logothetis, 2008). Furthermore, not all experimental tasks are feasible inside the scanner, and fMRI experiments are costly and participants with contraindications are often excluded (Glover, 2011; Huettel et al., 2008). These constraints can be critical in oxytocin administration studies, where the effects often are transient and mediated by rapid neuromodulatory shifts that are not reflected in hemodynamic signals alone (Bethlehem et al., 2013; Meyer-Lindenberg et al., 2011; Quintana & Guastella, 2020). This underscores the value of integrating complementary neuroimaging methods, and electroencephalography (EEG) offers a valuable alternative by providing superior temporal resolution, as it directly records neural electrical activity as it unfolds (Michel & Murray, 2012). EEG detects voltage fluctuations from postsynaptic potentials in cortical pyramidal neurons. When receiving synaptic input, post-synaptic exhibitory and inhibitory potentials are generated, leading to intracellular currents along the dendrites (Holmes & Khazipov, 2007) that generate extracellular fields when summated, which can be recorded by scalp electrodes (Kirschstein & Köhling, 2009).While EEG signals reflect summed activity of large neuronal populations and cannot directly measure oxytocin receptor activity (Bewernitz & Derendorf, 2012; Michel & Brunet, 2019) EEG can investigate the functional consequences of oxytocin modulation in the brain via neural oscillations and connectivity patterns(Bewernitz & Derendorf, 2012). EEG can thus provide information on underlying neural populations associated with cognitive, emotional, motor and sensory processes (Cohen, 2017; Gkintoni et al., 2025).

The high temporal resolution, cost-effectiveness and task flexibility of EEG lend it to unique ways of investigating neural activity, supporting different research approaches: A task or hypothesis-driven approach may focus on neural patterns directly linked to particular psychological functions such as cognitive processing, social functioning or behaviour. In contrast, an exploratory approach investigates spontaneous or large-scale neural activity, aiming to index brain states or network properties that may not be tied to specific behaviours or stimuli. As these two approaches can capture different aspects of brain function and their results can differ markedly, this may indicate that findings from such divergent methodologies should be interpreted within their distinct conceptual frameworks (Niedermeyer, 2005). EEG studies may also employ a diverse range of measurements and analyses affecting cross-study interpretations. The often-utilised event-related potentials (ERPs) capture time and phase-locked activity following specific events, such as modulations of the N170 component, which typically reflects early stages of face processing (Rossion et al., 2003; Schindler et al., 2023). Another common measure is that of spectral power, which allow investigations of a wide range of oscillatory activity, such as alpha asymmetry to index approach-motivation, executive functioning and anxiety (Smith et al., 2017), or theta band activity related to cortico-hippocampal interactions (Başar et al., 2000). The versatility of EEG imposes critical considerations for the researches, as the different measures emphasise distinct temporal, spectral, or network-level brain properties, thus determining which neural phenomena can be detected and interpreted (Jackson & Bolger, 2014; Niedermeyer, 2005; H. Zhang et al., 2023).

The research on oxytocin administration has produced mixed results, and this heterogeneity extends across several domains (Audunsdottir et al., 2024; Kang et al., 2025; Keech et al., 2018; Mierop et al., 2020; Quintana et al., 2021; Winterton et al., 2021). Several variables have been mentioned to explain this heterogeneity. For example, different participants’ ages in a study cohort can moderate the efficacy of oxytocin administration (Audunsdottir & Quintana, 2022; Ebner et al., 2016; Horta et al., 2023; Huffmeijer et al., 2012), it remains to be clarified whether results from clinical populations are transferable to neurotypical populations and vice-versa (Quintana et al., 2021), and the sex-specific roles of oxytocin are well known (McCall & Singer, 2012; Procyshyn et al., 2024; Quintana et al., 2024; Williams, 2007) - yet most oxytocin studies have been conducted in males (Dumais & Veenema, 2016).Furthermore, dose, dosing scheme – chronic, long-term or single dose - and time between administration should be considered (Quintana et al., 2021). For example, while it is suggested that a lower dose of oxytocin may be more efficacious than a higher dose (Martins et al., 2022; Quintana et al., 2015, 2017), there is still uncertainty about the most effective dose across populations and tasks (Quintana et al., 2021). Additionally, as most intranasal oxytocin studies in the social and cognitive domain utilise between-participant designs (Walum et al., 2016), a potential influence of opting for a between-or within-participant design should be investigated. Within-participant designs reduce error variance by controlling for individual variability and enhances statistical power with less required participants than between-participant studies, but there are risks of carryover or habituation effects with repeated task exposure. These design considerations highlight the complexity of oxytocin research and underscore the need for careful methodological choices.

Relative to other modalities, EEG research on oxytocin is in its infancy, with great diversity in not only EEG measurements and equipment, but also populations, dose and experimental tasks (Wigton et al., 2015). To our knowledge, no meta-analysis has yet been published that synthesizes the findings of these studies. Given the variation in research and findings so far, there is a need for comprehensive meta-analyses that not only synthesize existing results but also account for heterogeneity effects across different populations and contexts, to clarify the nuanced role of oxytocin in various psychological and neurobiological processes. This paper addresses gaps in summarized evidence on oxytocin’s effects on neural activity measured by EEG in humans through meta-analysis and a systematic review. It examines a wide and diverse range of study designs, EEG paradigms and neural modulations following oxytocin administration, while accounting for potential moderating factors, in order to provide a comprehensive overview of the current literature, its limitations and potential clinical implications. Furthermore, the paper aims to inform future research by discussing challenges, important methodological considerations and highlighting best practices.

## Methods

The Statement for Reporting Literature Searches in Systematic Reviews, as outlined by the Preferred Reporting Items for Systematic Reviews and Meta-Analysis (PRISMA) (Page et al., 2021) was followed. The initial literature search was performed June 22^nd^, 2023, with an updated follow-up search July 23^rd^, 2024.

To be eligible for inclusion, the studies had to 1) administer oxytocin exogenously, 2) measure the effects of oxytocin versus placebo using EEG, 3) include human participants. There were no restrictions on the type of EEG measurement used, type of cap, number of electrodes nor recording time. The literature search was limited to studies published in English or in any of the Scandinavian languages and no restrictions in terms of publication year or publication status were imposed. The first stage of study eligibility evaluation was done independently by the first author of this study (ED) via screening the title and abstract of each article. If the article could not be included or excluded based on the title and abstract alone, the full copy of the article was reviewed.

### Data extraction

Data was extracted from each included study to calculate the standardised mean difference (between-participants) and standardised mean change (within-participants) between oxytocin and placebo conditions, as measured with EEG. Many of the studies included multiple outcomes and EEG measures, where the latter broadly denotes quantitative metrics such as time-locked voltage fluctuations (quantified in microvolts and milliseconds), frequency-domain metrics such as spectral power (measured in hertz and amplitude), as well as connectivity and complexity indices. Only the means and standard deviations for the main outcomes were extracted from each article, wherever this was clearly stated in the article, for example via the main hypothesis. In cases where the article did not report the raw means, but rather means such as estimated marginal means, the raw means were requested from the authors. In addition to means, standard deviations of the main outcome(s), and the number of participants, other study characteristics were extracted, such as mean age and mean age standard deviation, sex ratio, publication year, publication status, EEG paradigm and measurement, study design, oxytocin administration method and dosing schedule.

Not all eligible studies could be included in this meta-analysis due to missing data (e.g. means, standard deviations/errors). 22 of the eligible studies did not report the means and standard deviations of the main outcome for both the oxytocin and placebo conditions in the article or the supplementary materials. As we did not put any restrictions on the type of EEG measurement being used, there was a wide variety of not only measures, but also ways to present the data. In the cases where the scores were presented via plots, Web Plot Digitizer (Rohatgi, 2022) was used to extract the mean scores and standard deviations. The precision of Web Plot Digitizer was tested by extracting data from plots where means and standard deviations also were reported in the text or tables. For the cases where means and standard deviations were not reported, or data was not possible to extract from plots, we attempted to contact the corresponding authors.

First author ED extracted data from all eligible studies, while author MM extracted a random 25% sample (determined by a random number generator). No inconsistencies were found between the two independent raters.

### Grouping data and direction of effects

There was a wide range of different outcomes, measurements, study designs and EEG paradigms utilised in the eligible studies, collectively capturing diverse temporal, spectral, and spatial characteristics of brain activity. As a means to compare the effect sizes extracted from the studies more accurately, each study was placed in one out of two categories based on its main outcome: neural correlates of social and/or cognitive processing and behaviour, or explorations of broader, less task-specific neural activity. Due to the differences between these categories and to better understand the impact and potential clinical relevance of the synthesised evidence from the included studies, two separate meta-analyses were conducted: one meta-analysis for the studies in the social and non-social/cognitive categories, and another for the studies in the neural category, henceforth referred to as Social/Cognitive MA and Neural MA, respectively.

When interpreting the results for these meta-analyses, it is important to note that the results of these analyses should be interpreted differently. Larger effects indicate that oxytocin administration was generally considered more beneficial than placebo on the different social and cognitive process outcomes in the Social/Cognitive MA. To reflect this, a negative effect size estimate was thus re-coded as positive if a negative value, for example a reduction in amplitude or latency, indicated that OT administration was more beneficial for the study outcome. The studies in the Neural MAare more exploratory investigations of neural activity, as opposed to neural correlates connected to specific social or cognitive outcomes. However, the direction of the effects was still re-coded if a negative effect estimate indicated an expected modulation of the neural activity investigated. A study expecting decreased duration in a specific microstate after oxytocin administration, for example, would be re-coded as positive, as the decrease (and thus the negative effect estimates) was the expected outcome. Both meta-analyses therefore investigate the effectiveness of oxytocin administration when compared to placebo, but the interpretation and potential implications of these results differ.

### Publication and small study bias assessments

Robust Bayesian meta-analysis (RoBMA) was conducted to directly investigate the possibility of publication and small study bias, by implementing a model-averaging approach that integrates multiple meta-analytic models, including those that account for selection bias as well as no bias (Bartoš & Maier, 2020). Separate analyses were run on the studies with the least and largest variances due to the wide variety of effect sizes associated with each study in both MAs. Risk of publication was also assessed with the Egger’s regression test, which was also deployed to assess small study bias via testing potential linear relationships between effect sizes and the standard errors (Egger et al., 1997). The results from Egger’s test was visualised via a contour-enhanced funnel plot (Peters et al., 2008). The use of these two complementary approaches, Egger’s test from the frequentist paradigm and RoBMA from the Bayesian paradigm, allowed us to cross-examine the evidence for publication bias from distinct statistical perspectives, thereby strengthening the robustness and interpretability of our bias assessment.

## Statistical analyses

All statistical analyses were conducted in RStudio (Posit team, 2025) for R version 4.4.2 (R Core Team, 2024), except for statistical power analyses with the metameta package (Quintana, 2023) where R version 4.3.0 was used to ensure package compatibility.

Otherwise, the latest version of all packages was used. We calculated the effects sizes from each study using distinct methods for the within-subject versus between-subject studies: standardised mean change (Cohen’s *d* with Hedges’ correction for between-measures correlation) and standardised mean difference for within-measures, respectively (Gibbons et al., 1993; Hedges, 1981). Effect sizes for each eligible study were computed with the metafor R package (Viechtbauer, 2010), using reported means, standard deviations, and the number of participants in each tested group. In cases where standard errors were reported, these were converted to standard deviations before conducting the meta-analysis. As previously stated, in cases where a negative effect size was considered beneficial by the authors of the study, these were coded as such and transformed to positive values. Meta-analyses were subsequently conducted using the metafor package (Viechtbauer, 2010). A multilevel analysis was chosen to limit the risk of inflated effect sizes due to most studies reporting more than one effect size, as having more than one effect from a study violates the assumption of independent effects sizes when performing a conventional meta-analysis. Reporting multiple effect sizes are common in neuroscience research, where multiple dependent or independent outcomes from different brain regions, tasks, or timepoints are provided. A multilevel model estimates both within-study and between-study variance, providing more accurate pooled estimates compared to a standard random-effects model (Cheung, 2013; Harrer et al., 2021). Forest plots using the ggplot2 package (Wickham, 2016) were used for visualisations of the meta-analyses.

As non-significant intranasal oxytocin study results may not necessarily indicate an absence of effect (Quintana, 2018), equivalence testing was performed to assess whether the observed effect size was smaller than a meaningful effect, via the TOSTER package (Caldwell, 2022; Lakens, 2017). The Two One-Sided Tests (TOST) procedure was used to determine whether the overall effect lays entirely within a predefined range of *practically meaningful* effects. As it can be difficult to determine what would constitute the smallest effect sizes of interest (SESOI), a range of equivalence bounds were used: ±0.10, ±0.20, and ±0.25 (Lakens, 2017; Marshall et al., 2024; Quintana, 2018)

Study heterogeneity was indexed by calculating Cochran’s Q and *I*^2^ to test if the variability in effect sizes exceeds that expected from random sampling error, and to quantify the proportion of total variance to index inconsistency across studies, respectively. As we expected heterogeneity due to between-study variance, a “Graphical Display of Study Heterogeneity” (GOSH) (Olkin et al., 2012) analysis was conducted using the metafor package (Viechtbauer, 2010). GOSH analyses run a meta-analysis on all possible subsets of the *k* studies included in a meta-analysis. The GOSH plot was supplemented by GOSH diagnostics via the dmetar function source code (Harrer et al., 2019), which uses the cluster algorithms *k-*means (Hartigan & Wong, 1979), density reachability and connectivity clustering (DBSCAN) (Schubert et al., 2017) and Gaussian Mixture Models (GMM)(Fraley & and Raftery, 2002) to investigate and identify potential outliers that contribute to the patterns of heterogeneity revealed by the GOSH plots (Harrer et al., 2021).

Using the metafor package, Leave-One-Out Analyses (LOO) were computed investigate potential influential studies and outliers in the meta-analyses by demonstrating the range of effects, systematically excluding one study at a time and assess the potential impact of each study on the overall effect size (Viechtbauer & Cheung, 2010).

To assess the influence of studies with extreme variance on the overall results, influence diagnostics and accompanying plots were conducted via the metafor package, where studentised residuals, difference in fits (DFFITS) values, Cook’s distances, leverage values, hats, weights, covariance ratio and LOO-estimates (τ² and QE) were extracted (Viechtbauer, 2010). As these diagnostics assume that effect sizes are independent, two versions of these diagnostics were run: One set of diagnostics including the effects with the *largest* variance and one set of diagnostics including the effects with the *least* variance.

All scripts and data sets are uploaded to OSF (https://osf.io/ynsd7/).

### Moderator analyses

Moderator variables were defined prior to screening and data analysis and included in the pre-registration of this meta-analysis and systematic review (Prospero ID: CRD42023433479). Based on previous recommendations (Fu et al., 2011), moderator analyses were conducted if there were at least five effect sizes in each defined subgroup. Moderator analyses were conducted using the metafor package and visualised via forest plots from the ggplot2 package.

The following moderator analyses were run for at least one of the two meta-analyses:

- Categorical: Type of population (clinical versus healthy or neurotypical), experimental task type, outcome measures, oxytocin dosage (24 IU or more than 24 IU), study design (within versus between participants)
- Continuous: Mean age of participants, percentage of female participants, time (in minutes) between oxytocin administration and testing.

Experimental task types were sorted into different categories based on what the participants were instructed to do by the study team, where each category contained five or more effect sizes. For example, if the participants were asked to do any activity, this would be labelled as “behavioural”, whereas the participant passively being exposed to one type of visual stimuli would be labelled “visual”. The following categories for the Social/Cognitive MA: “Behavioural”, “visual” and “visual and behavioural”. For the Neural MA, the categories were: “resting-state” and “visual”. The different EEG outcome measures were grouped into different categories, where each category contained at least five effect sizes. For the Social/cognitive MA, the categories were: “Frequency bands”, “ERPs”, “symmetry/asymmetry”, “frequency domain” and “microstates”. For the Neural MA, there were only two EEG outcome measure categories: “microstates” and “frequency bands”. Note that these categories are umbrella categories: The studies in each of these categories still report a variety of different measures and units of measurements, e.g. investigating different ERP components and outcomes in both amplitudes and milliseconds in the “ERP” category.

Based on the literature pointing out integral differences between microstate analysis measurement outcomes (Michel & Koenig, 2018), we computed one post-hoc moderator analysis on outcome measures for the meta-analysis on neural outcomes. Here, the microstate analysis group was separated into four distinct groups. However, it must be noted that the number of effects in two of the groups were under the lower limit of 5. Therefore, these results should be interpreted as more exploratory and can be found in *Supplementary Materials*.

## Results

### Study selection

We identified 964 records from the initial systematic literature search (see *Figure 1*). After screening titles and abstracts, 42 studies were considered relevant for further evaluation. The subsequent literature search conducted one year later yielded an additional 75 records, of which 4 new studies were deemed relevant after title and abstract screening. This resulted in a total of 46 studies for full-text review.

**Figure 1:**
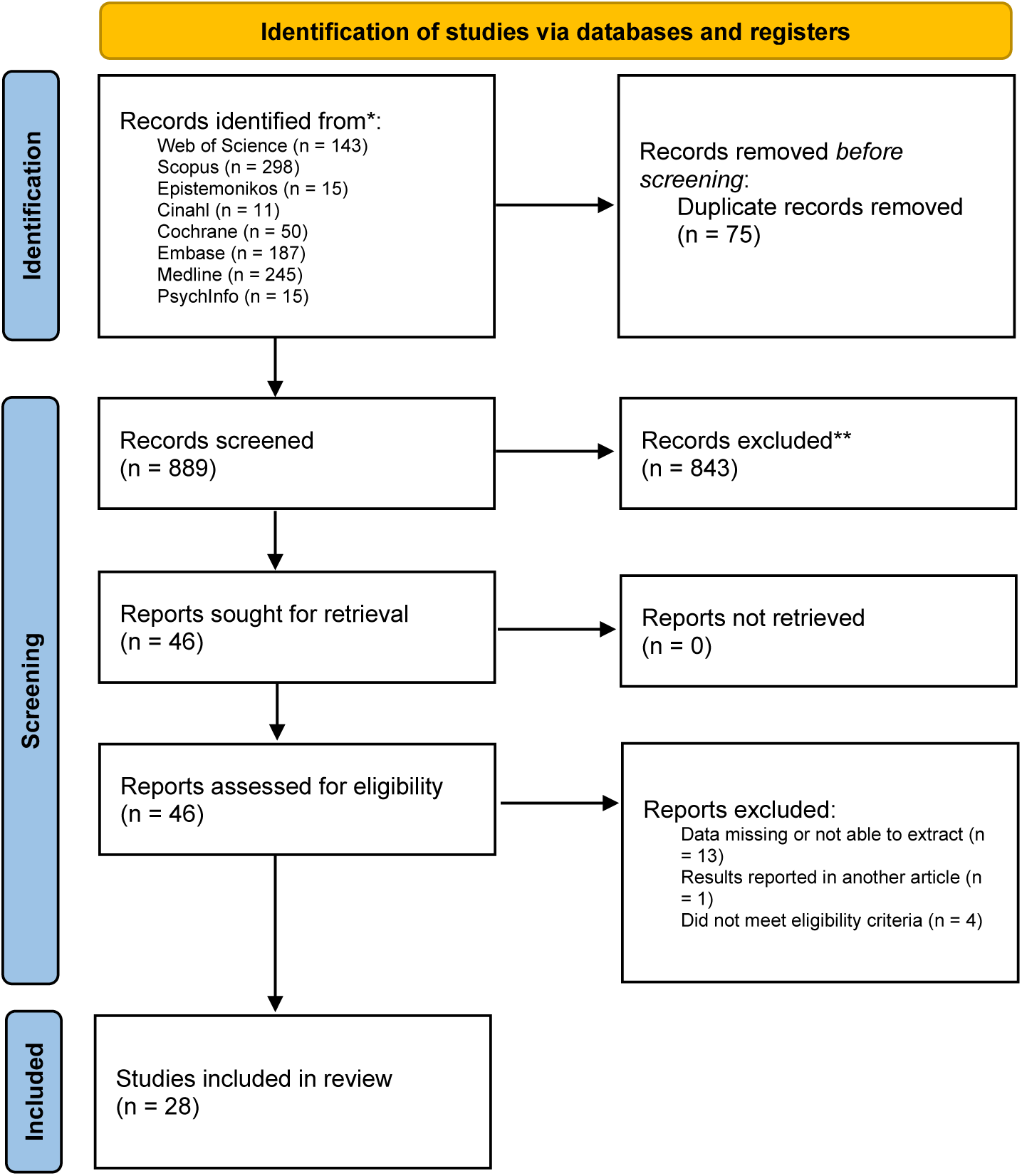
PRISMA flow diagram

Following full-text screening, 42 studies met the eligibility criteria. We contacted 17 authors to request additional data or extracted effect sizes from figures using WebPlotDigitizer where necessary (5 studies). 41.2% of contacted authors provided means and their variances. This resulted in 28 studies included in the final meta-analysis, with a total of 161 extracted outcome effect sizes. All eligible studies were peer-reviewed and published, double-blinded, randomized, controlled and compared intranasally administered oxytocin and placebo.

### Study characteristics

The total number of participants was 1361 (319 females and 199 belonging to a group with a neurodevelopmental differences or psychiatric diagnosis), and the mean age across all studies was 23.99 years (10.1-47.0 years). Only two studies (Alaerts et al., 2024; Moerkerke et al., 2023) employed a chronic dosing scheme design where the participants received two single 12 IU doses of oxytocin daily over a period of four weeks. The other studies gave the participants a single administration of oxytocin before testing, and the time between administration and testing ranged from 5 to 90 minutes.

The included studies used a wide variety of different EEG measurements, tasks and outcomes, and there were similarly diverse regions of interest (see *Supplementary Materials* for an overview of all included studies). The type of tasks employed in each study was labelled either visual, behavioural, or visual and behavioural. One study (Ochiai et al., 2021) used an auditory task, but this was placed in the behavioural category when conducting moderator analyses.

All studies in the Social/Cognitive MA included evoked EEG signals, even if their task and specific EEG measurements differed, and oxytocin administration was hypothesised to modulate specific EEG markers linked to social and/or cognitive processes. In the Neural MA, all but one study investigated spontaneous neural activity using resting-state tasks; one study (Perry et al., 2010) utilised a visual stimuli task. The study focused on neural modulations suppressed by biological motion, and specifically the involvement of the mirror neuron system (MNS) and broader brain structures. Although a potential link to social cognition was postulated, this was on a more exploratory level than the studies in the Social/Cognitive MA, with more focus on mechanisms than potential beneficial effects of the oxytocin administration – perhaps in part due to being one of the first studies to investigate the effects of oxytocin administration on modulations of EEG rhythms.

### Synthesis of results

As previously mentioned, we conducted two separate meta-analyses due to integral differences in outcomes. For the Social/Cognitive MA, studies in the “social” and “non-social” categories typically expected a modulation of EEG measurements linked to specific social/non-social processes, for example an increase in amplitude of the face-sensitive N170 component after oxytocin administration when viewing faces of adults and infants (Peltola et al., 2018). For the Neural MA, which contains the studies in the “neural” category, these did not explicitly link the outcomes to improvements in social or cognitive function and were typically more exploratory in nature. Moderator analyses were computed if the moderator variables contained at least five effect sizes each. The planned moderator analysis comparing chronic versus single dose administration did not fulfil these criteria and was thus not run in either meta-analysis. For a comprehensive overview of studies included in the two meta-analyses and their study characteristics, see *Overview of included articles* in *Supplementary Materials*.

### Meta analysis 1: Social and cognitive processing

A multilevel meta-analysis utilising the restricted maximum likelihood estimator (REML)(Viechtbauer, 2010) was performed on the studies with social and cognitive outcomes. This meta-analysis included a total of 113 different effect sizes across 21 studies and yielded a summary effect size of Hedge’s *g* = 0.14 [CI 0.05, 0.23] (*p* = 0.0016), indicating a beneficial effect of oxytocin administration compared to placebo on neural correlates of social and cognitive processing (Figure 2A and 2C). For a detailed forest plot, see *Supplementary Materials*.

**Figure 2.**
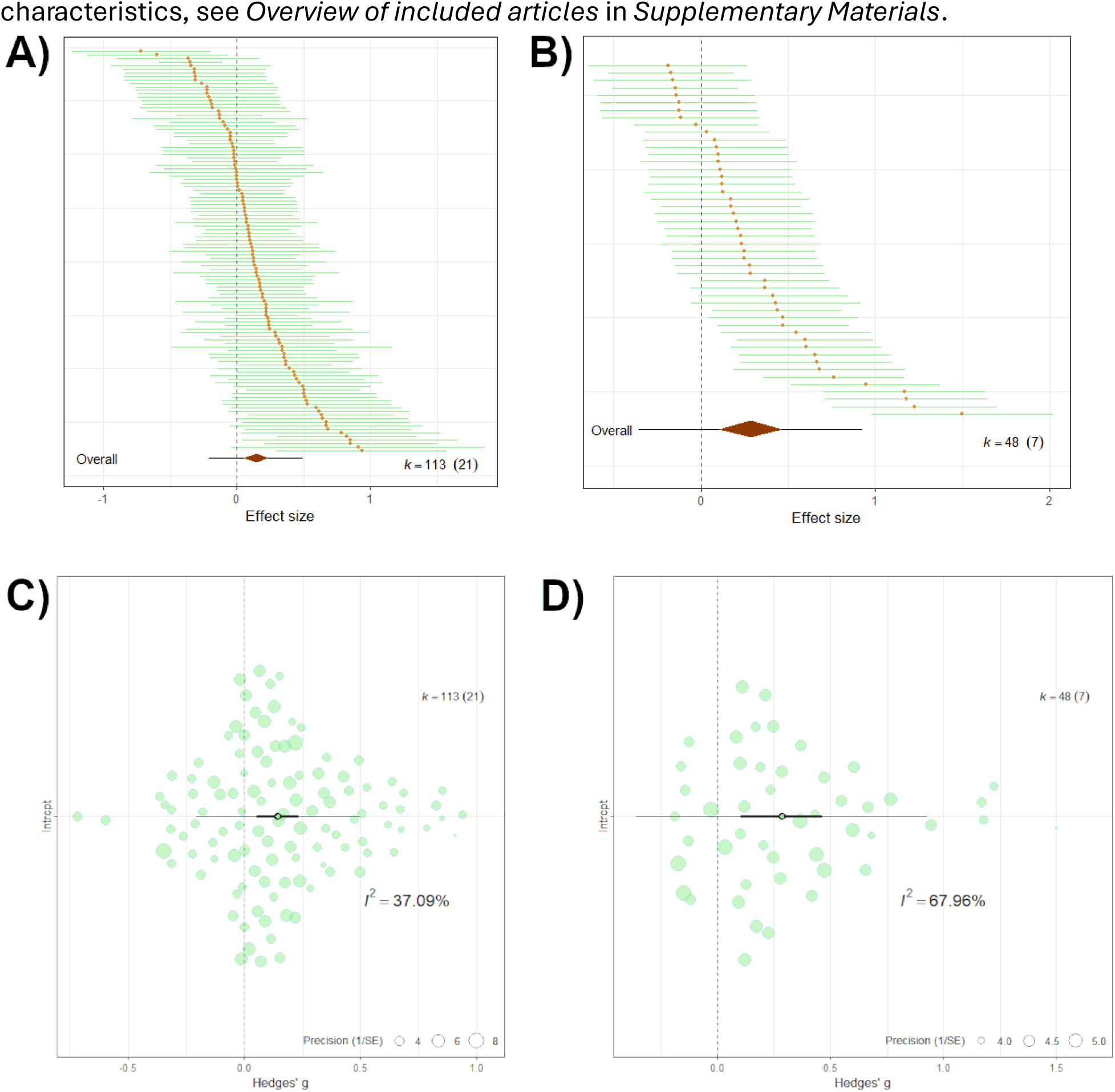
Top row (plots A and B): Caterpillar plots for Social/Cognitive MA (A) and Neural MA. **(B)** visualising individual study effect sizes (Hedges’ g) and their 95% confidence intervals. The solid horizontal line represents the pooled estimate from the random-effects meta-analysis model. The spread and overlap of confidence intervals reflect both the precision and heterogeneity among studies. Studies are ordered by effect size magnitude. K refers to the number of effect sizes in total for each meta-analysis. Numbers in parentheses refer to the number of studies in each meta-analysis. **Bottom row (plots C and D): Orchard plots for Social/Cognitive MA (C) and Neural MA (D)** visualising the overall effect size (Hedges’ g) from the meta-analyses. The central horizontal bar represents the intercept estimates, with the bold horizontal line representing the 95% confidence intervals. Jittered dots represent individual effect sizes, where the size of each dot reflects the inverse variance (precision) of the corresponding effect size. I²-statistics for each meta-analysis is included.

Equivalence testing revealed that the observed effect cannot be confidently interpreted as negligible with equivalence bounds set to *d* = 0.10 (*Z* = 1.06, *p* = 0.85) and *d* = 0.20 (*Z* =-1.05, *p* = 0.15), but not *d* = 0.25 (*Z* =-2.11, *p* = 0.017). Figures visualising the equivalence testing results are available in *Supplementary Materials*.

As expected, there was significant heterogeneity (*QE* = 149.98, *p* < 0.01), warranting additional heterogeneity analyses and moderator analyses to better understand the variance in the data.

### Publication and small study bias assessments

A Robust Bayesian meta-analysis revealed an inclusion Bayes factors for publication bias of *BF_least_* = 1.21 and *BF_largest_* = 9.57 for the studies with the least and largest variance respectively. This provides anecdotal to moderate evidence for the influence of small-study effects or other forms of bias, especially for the studies with most variance. Visual inspection of a funnel plot (*Figure 3A*) suggests some asymmetry, which can be indicative of small-study effects or publication bias. This observation was confirmed by Egger’s regression test for funnel plot asymmetry, which yielded a significant result (*t*(111) = 2.74, *p* = 0.0072), with a bias estimate of 18.5842 (*SE* = 6.7824), suggesting that there is a likelihood of systematic asymmetry.

**Figure 3.**
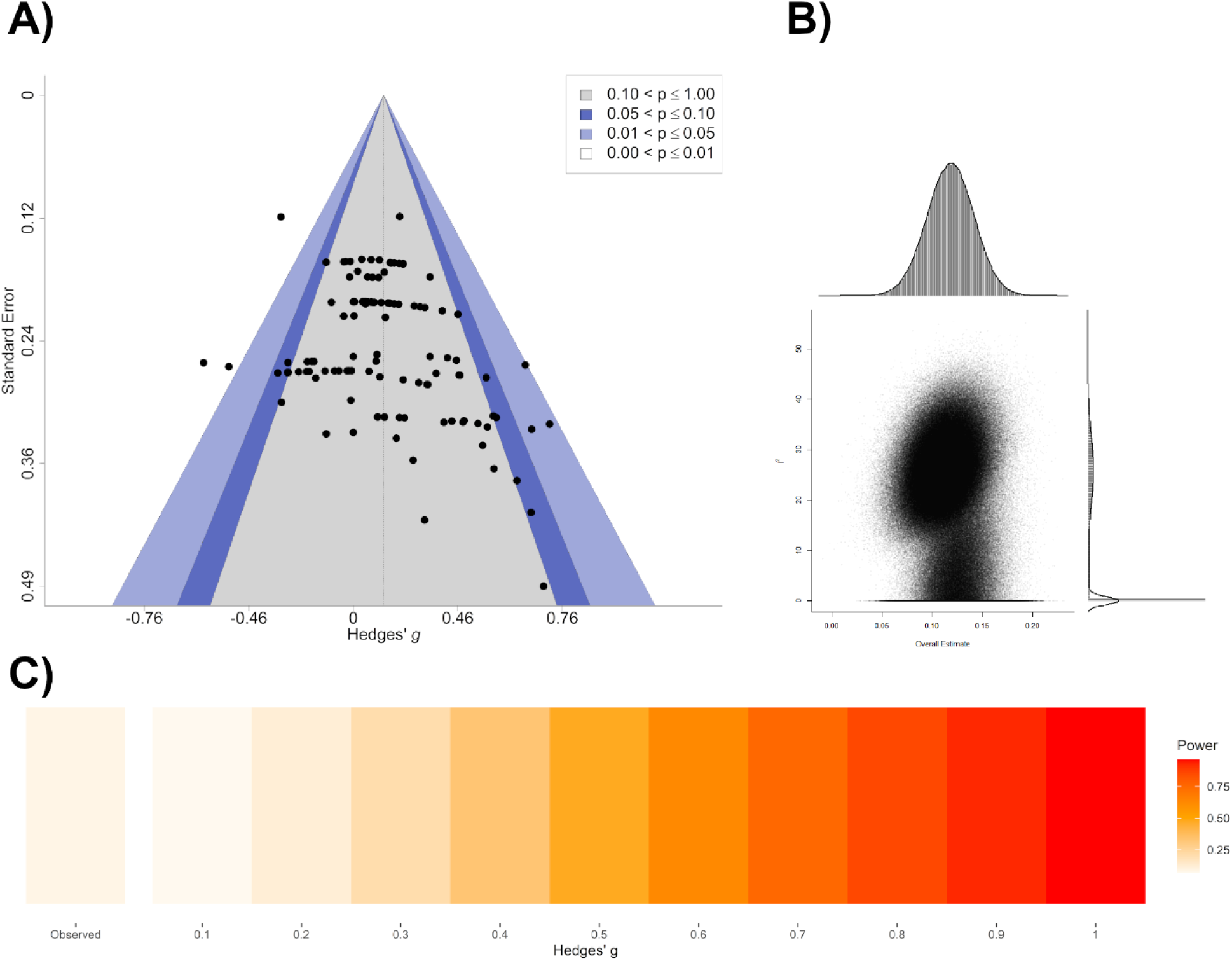
A) Funnel plot illustrating the relationship between the effect sizes (Hedges’ g) and their corresponding standard errors. The shaded areas represent regions of statistical significance: grey (p > 0.10), light blue (0.05 < p ≤ 0.10), medium blue (0.01 < p ≤ 0.05), and white (p ≤ 0.01). Visual inspection reveals slight asymmetry, suggesting potential publication bias or small-study effects. **B) GOSH plot** of the social and non-social outcomes, visualising overall effect size distributions and study heterogeneity. The x-axis represents the pooled effect size, and the y-axis represents the heterogeneity statistic, while each point corresponds to the results from a unique subset of studies. The central cluster around 0.10 to 0.15 reflects a small positive effect size that remains consistent across subsets, while the vertical spread indicates low-to-moderate heterogeneity (I² values ranging from 0% to 55%). The marginal histograms display the density of effect size estimates (top) and heterogeneity values (right). **C) Firepower plot** visualising of the median statistical power of the meta-analysis of the social and non-social outcomes, assuming the observed overall effect is the true effect size. The observed effect is seen to the far left. Lighter colours reflect lower statistical power, which increases as the colour darkens and become more red.

### Study heterogeneity and power

*I²*-statistics revealed that 32.93% and 4.16% of the total variance could be attributed to between-study heterogeneity and within-study heterogeneity, respectively. A robust Bayesian meta-analysis found a substantial degree of heterogeneity for the studies with the largest variance (*BF_rf_* = 414.438, model-averaged estimate *τ* = 0.351, CI [0.178, 0.572]). The studies with least variance, as expected, reported far more consistent and less heterogenous effect sizes across the studies *(BF_rf_* = 0.288, model-averaged estimate *τ* = 0.021, CI [0.000, 0.147])

*Figure 3B* displays the GOSH plot generated for the meta-analysis on social and non-social outcomes. While multilevel and Robust Bayesian meta-analyses indicated high heterogeneity, the GOSH analysis revealed that most study subsets exhibit low-to-moderate heterogeneity, suggesting that a subset of studies disproportionately contributes to the between-study variance (mean *QE* = 74.30 [*QE*_min_ = 21.42, *QE*_max_ = 126.25], mean τ^2^ = 0.011 [τ ^2^_min_ = 0, τ^2^_max_ = 0.067).

Power calculations revealed a median of statistical power to detect the effect size (Hedge’s g = 0.14) of only 8.5%. Assuming the effect size of 0.14 is the ‘true’ effect, the included studies’ statistical power to reliably detect an effect size of 0.1 or 0.2 ranged from 5.5% to 13.3% and 7% to 38.5%, respectively. See Figure 3C for a visualisation of median statistical power.

### Influence and sensitivity analysis

A leave-one-out analysis (LOO) was conducted to identify potential outliers. The overall effect size remained stable across all iterations, with no single study substantially altering the pooled estimate or heterogeneity statistics (*Hedges’ g* range: 0.135 to 0.159, *QE* range: 135.12 to 149.99). Influence diagnostics on the studies with the least and largest variances were subsequently computed to investigate studies exerting more subtle influence. By visual inspection, two studies (Moerkerke et al., 2023; Schiller et al., 2023) stood out as potential outliers. GOSH diagnostics listed outcome number 58 as a recurring outlier. This potential outlier (Schiller et al., 2023) had the highest estimate in the LOO and was also highlighted as an influential study when running influence diagnostics. Information on influence diagnostics and the remaining potential outliers identified by GOSH diagnostics are listed in *Supplementary Materials*.

Based on the results of the GOSH diagnostics and influence diagnostics, a meta-analysis with potential outliers removed was conducted with outcome 58 removed, assuming a conservative approach. The removal improved model fit and decreased residual heterogeneity (*QE* = 135.12, *p* = 0.06). The summary effect size increased slightly after outlier removal (Hedges’ *g* = 0.16, *p* = 0.0003). The differences between the original multilevel meta-analysis and the multilevel meta-analysis with the outlier removed, as well as accompanying figures, are summarised in *Supplementary Materials*.

### Moderator analyses

To investigate the influence of subgroups on the summary effect, moderator analyses were conducted. The moderator analyses were pre-specified and calculated if each subgroup contained at least five effects. Results from the statistically significant moderator analyses are summarised in *Table 1*. Plots for moderator analyses and non-significant moderator tests are available in *Supplementary Materials*.

**Table 1:**
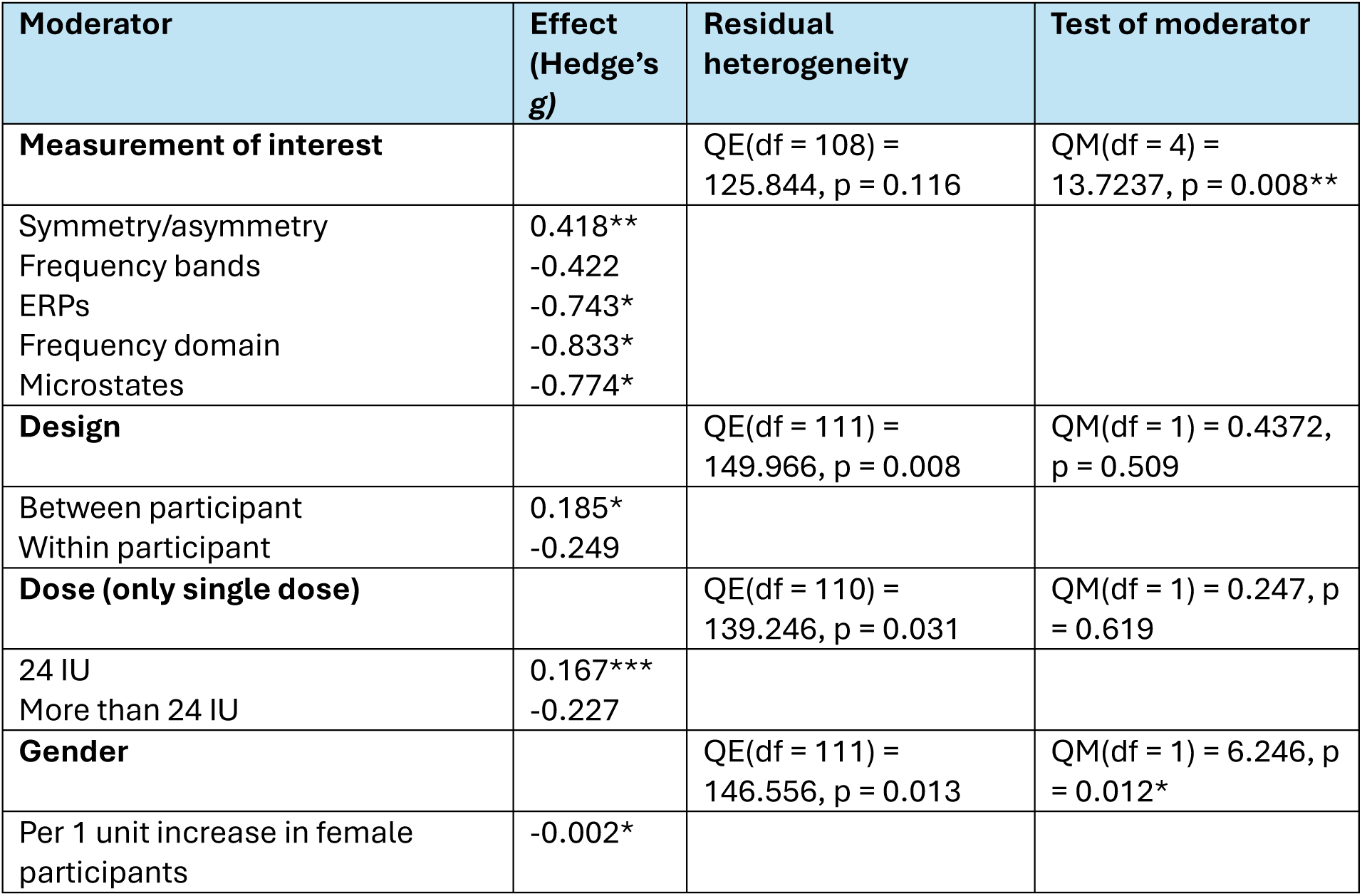
Overview of moderator analyses for social and non-social outcomes. Listed effects for all variables are real effects, i.e. the sum after subtracting the intercept value from the moderator. For each categorical moderator analysis in the table, the first listed effect is the intercept. For continuous moderators, the intercept is not listed. Significant moderators are highlighted with an asterisk (* = 0.05, ** = 0.01, *** = 0.001). Significant residual heterogeneity is not highlighted. QE =residual heterogeneity statistic, QM = model heterogeneity statistic.

| Moderator | Effect (Hedge's $g$) | Residual heterogeneity | Test of moderator |
| --- | --- | --- | --- |
| **Measurement of interest** | | QE(df = 108) = 125.844, $p = 0.116$ | QM(df = 4) = 13.7237, $p = 0.008^{**}$ |
| Symmetry/asymmetry | 0.418<sup>**</sup> |  |  |
| Frequency bands | -0.422 |  |  |
| ERPs | -0.743<sup>*</sup> |  |  |
| Frequency domain | -0.833<sup>*</sup> |  |  |
| Microstates | -0.774<sup>*</sup> |  |  |
| **Design** | | QE(df = 111) = 149.966, $p = 0.008$ | QM(df = 1) = 0.4372, $p = 0.509$ |
| Between participant | 0.185<sup>*</sup> |  |  |
| Within participant | -0.249 |  |  |
| **Dose (only single dose)** | | QE(df = 110) = 139.246, $p = 0.031$ | QM(df = 1) = 0.247, $p = 0.619$ |
| 24 IU | 0.167<sup>***</sup> |  |  |
| More than 24 IU | -0.227 |  |  |
| **Gender** | | QE(df = 111) = 146.556, $p = 0.013$ | QM(df = 1) = 6.246, $p = 0.012^{*}$ |
| Per 1 unit increase in female participants | -0.002<sup>*</sup> |  |  |

As there was a very wide variety of specific EEG outcome analyses and measurements conducted, the extracted effects from each study were sorted into one of the following groups, *frequency bands (N= 14), ERPs (N = 76), symmetry/asymmetry (N = 10), frequency domain (N = 7), and microstates (N = 6)*.

The “gender” (i.e. percentage of female participants), mean “age”, and “time in minutes between administration and testing” moderators were treated as continuous variables, where the listed effect is per 1 unit increase.

The “gender” moderator was statistically significant (p < 0.05), where percentage increases in female participants lead to decreases in the effect of oxytocin administration on the outcomes. The “measurement of interest” moderator was also statistically significant (p < 0.01), inferring that the type of EEG measurement utilised in the study influenced the observed effect. In this moderator analysis, the symmetry/asymmetry, ERP, frequency domain and microstates measurements were statistically significant. While the moderator analyses for study design and dose were not statistically significant, the “between-participant design” (*g* = 0.18, *p <* 0.05) and “24IU dose” (*g* = 0.17, *p* < 0.001) subgroups, respectively, were statistically significant. Neither explained a significant amount of the variance in the data.

### Meta analysis 2: Exploratory neural outcomes

A multilevel meta-analysis utilising REML was also performed on the studies in the Neural MA, which included a total of 48 different effect sizes across 7 studies. A summary effect size of Hedge’s *g* = 0.28 [CI 0.10, 0.46] (*p* = 0.0021) was found. As previously mentioned, the results of this meta-analysis must be understood differently than the Social/Cognitive MA. These studies hypothesised increases or decreases in neural activity after oxytocin administration compared to placebo on a more exploratory basis: predominantly spontaneous EEG activity related to brain states or networks not explicitly connected to improvements in social or cognitive processing or behaviour. As shown in the orchard and caterpillar plots (Figure 2B *and* figure 2D), these results are therefore listed as a “decrease” vs “increase” in neural activity. For a detailed forest plot, see *Supplementary Materials*.

Equivalence testing with a range of equivalence bounds were non-significant, *d* = 0.10 (*Z* = 2.008, *p* = 0.978), *d* = 0.20 (*Z* = 0.951, *p* = 0.829) and *d* = 0.25 (*Z* = 0.423, *p* = 0.664), indicating that the observed effect is not negligible.

There was significant heterogeneity (*QE* = 139.8, *p* < 0.001), suggesting that heterogeneity analyses and moderator analyses were warranted.

### Publication and small study bias assessments

Conducting a Robust Bayesian meta-analysis on the studies with the least and most variance revealed an inclusion Bayes factor for bias of *BFleast* = 1.17 and *BFlargest* = 1.29, providing minimal support for a bias model in both the data with the least and largest variances. Assessing publication bias was also done by calculating Egger’s linear regression and visualising this via a funnel plot (Figure 4A). Egger’s regression test for funnel plot asymmetry revealed no asymmetry indicative of publication bias or small-study effects, *t*(46) = 0.98, *p* = 0.332 with a bias estimate of 34.186 (*SE* = 32.89).

**Figure 4:**
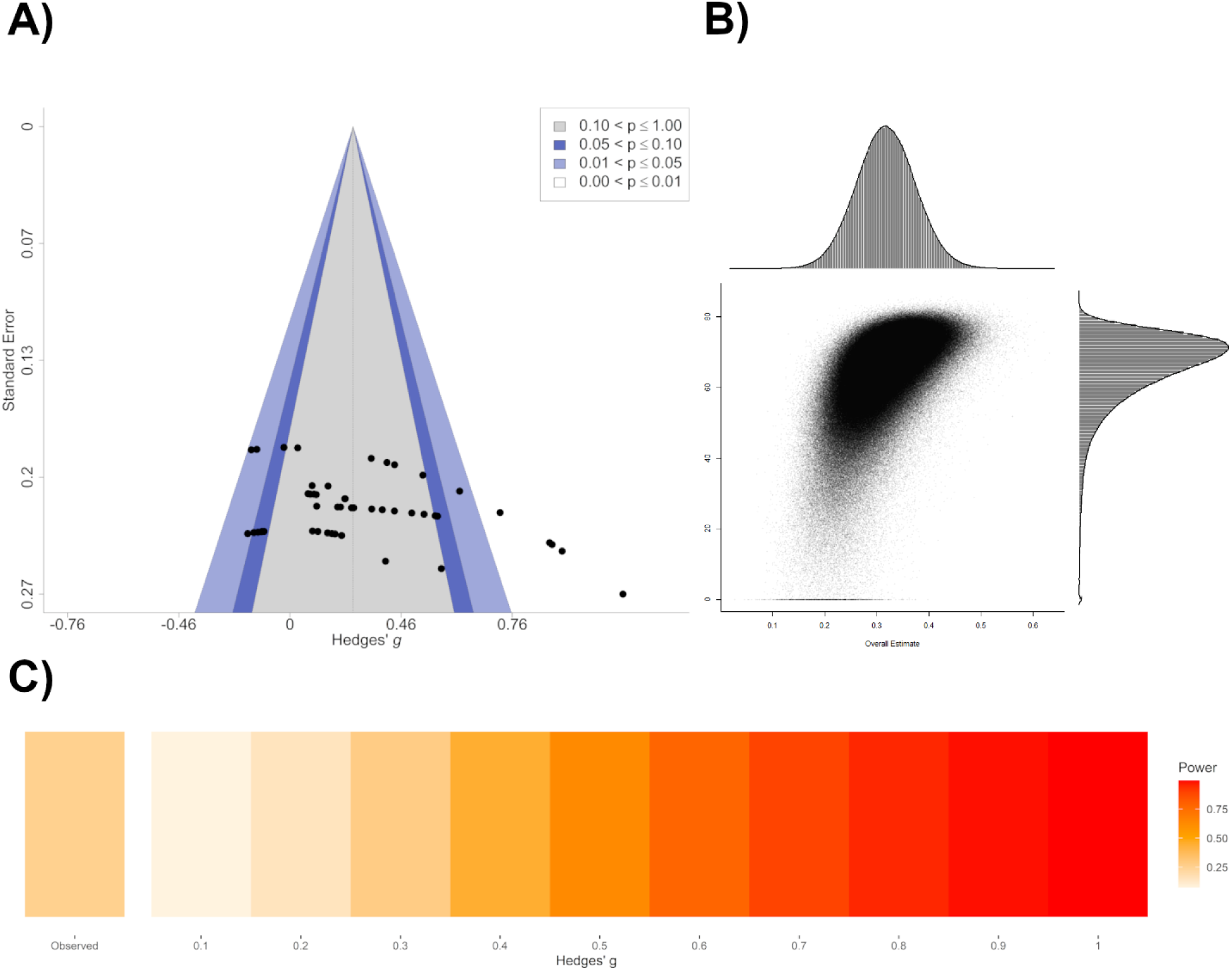
A) Funnel plot illustrating the relationship between the effect sizes (Hedges’ g) and their corresponding standard errors. The shaded areas represent regions of statistical significance: grey (p > 0.10), light blue (0.05 < p ≤ 0.10), medium blue (0.01 < p ≤ 0.05), and white (p ≤ 0.01). Visual inspection reveals asymmetry, suggesting potential publication bias or small-study effects. **B) GOSH plot** of neural outcomes, showing the distribution of effect sizes and study heterogeneity. The x-axis represents the pooled effect size, and the y-axis shows the heterogeneity statistic, with each point reflecting a unique study subset. Most estimates ranged between approximately 0.20 and 0.50 on the x-axis, indicating a generally positive and moderately small effect size. I² values in the 40% to 80% range indicate substantial variation. Marginal histograms display the density of effect sizes (top) and heterogeneity values (right). **C) Firepower plot** visualising of the median statistical power of the meta-analysis of the social and non-social outcomes, assuming the observed overall effect is the true effect size. The observed effect is seen to the far left. Lighter colours reflect lower statistical power, which increases as the colour darkens and become more red.

However, the Egger’s linear regression revealed a τ² value of 3455.56, indicating that a substantial amount of heterogeneity in effect sizes across studies may not only be explained by inverse variance but may also be due to the presence of bias or extreme values or outliers.

### Study heterogeneity and power

*I²*-statistics revealed 22.89% between-study heterogeneity and 41.41% within-study heterogeneity. Robust Bayesian meta-analysis on the studies with the least and the largest variance revealed a moderately high level of heterogeneity for the most-variance studies (Least: *BF_rf_* = 0.38, model-averaged estimate *τ* = 0.32; Largest: *BF_rf_* = 9.15, model-averaged estimate *τ* = 0.37), albeit far smaller than that seen in the meta-analysis on social and non-social outcomes.

The GOSH plot (Figure 4B) revealed a concentrated, elliptical-shaped cluster of generally positive effect estimates and a high level of heterogeneity, *QE* = 74.30 [*QE*_min_ = 21.42, *QE*_max_ = 126.25] and mean τ^2^ = 0.098 [τ ^2^_min_ = 0, τ^2^_max_ = 0.029).

A power calculation for the individual studies in the neural outcomes provided a median of statistical power to detect the effect size (Hedge’s g = 0.31) of 25.5%. The included studies’ statistical power to detect an effect size of 0.3 reliably ranged from 20.4% to 37.6%. A visualisation of median statistical power in the meta-analysis is presented in Figure 4C.

### Influence and sensitivity analysis

Computing a LOO analysis did not identify any outliers. To detect potentially more subtle influences, we ran influence and GOSH diagnostics. Influence diagnostics for studies with the smallest and largest variances are shown in *Supplementary Materials*. Visually, the study by Tomescu et al. (2024) appeared as a potential outlier, but it did not boast large leverage values, which indicate that the study doesn’t heavily influence the model parameter estimate, warranting further investigation. GOSH diagnostics were subsequently performed, listing outcomes 34, 40, 41 and 43 (see *Supplementary Materials)* from Tomescu et al. (2024) as outliers. While all these potential outliers have effect sizes larger than 1, but neither of these individual outcomes stood out when investigating leverage and influence at an outcome level, suggesting that these effect sizes may not have much influence on the overall meta-analysis result.

A new multilevel meta-analysis without the potential outlier outcomes 34, 40, 41 and 43 substantially altered model fit and heterogeneity, although there was still statistically significant heterogeneity at *QE* = 73.68, *p* = 0.003. The summary effect size was only marginally altered (Hedges’ *g* = 0.23, *p* = 0.0008). A summary of the differences between the two models and accompanying figures can be found in *Supplementary Materials*.

### Moderator analyses

Moderator analyses were conducted as pre-specified. *Table 3* summarises significant results from the moderator analyses. None of the moderator analyses reached statistical significance, and neither were able to significantly explain the variation in the data. Two moderators were able to reach statistical significance, namely the gender intercept at 0.01 level (Hedges’ *g* = 0.30), i.e. when there are no female participants, and the between-participant moderator at the 0.05 level (Hedges’ *g* = 0.37). All moderator analysis plots and tests can be found in *Supplementary Materials*.

**Table 3:** Overview of moderator analyses for neural outcomes. Listed effects for all moderator variables are real effects, i.e. the sum after subtracting the intercept value from the moderator. For each categorical moderator analysis in the table, the first listed effect is the intercept. Intercepts are not listed for continuous moderators. Significant moderators are highlighted with an asterisk (* = 0.05, ** = 0.01, *** = 0.001). Significant residual heterogeneity is not highlighted. QE =residual heterogeneity statistic, QM = model heterogeneity statistic.

| Moderator | Effect (Hedges' $g$) | Residual heterogeneity | Test of moderator |
| --- | --- | --- | --- |
| **Design** | | $QE(df = 46) = 124.822$,<br>$p < .0001$ | $QM(df = 1) = 0.311$, $p = 0.577$ |
| Between participant | 0.368* |  |  |
| Within participant | 0.231 |  |  |
| **Gender** | | $QE(df = 46) = 138.442$,<br>$p < .0001$ | $QM(df = 1) = 0.113$, $p = 0.737$ |
| Per 1 unit increase in female participants | -0.001 |  |  |

## Discussion

In this article, we performed two multilevel random-effects meta-analyses with a total of 161 effect sizes across 28 studies. We found some tentative evidence that oxytocin administration had a significant effect on EEG correlates related to social and cognitive processing [*g* = 0.14 (0.05, 0.23)] and on exploratory neural modulations [*g* = 0.28 (0.10, 0.46)]. Equivalence tests with different smallest effect sizes of interest were performed to assess whether the observed effects were negligible. In all but one cases did we find them to be non-significant, revealing that even though the observed effect sizes are small, they may still hold practical relevance. However, it should be noted that oxytocin’s effects on behavioural and cognitive outcomes are, on average, smaller than these effects (Audunsdottir et al., 2024; Kang et al., 2025). Running Robust Bayesian meta-analyses on the effect sizes with the least and the largest variances per study, the estimated summary effect sizes were anecdotal, especially for the studies in the Social/Cognitive MA. The discrepancy between the Bayesian and frequentist effect size estimates may arise from the models’ different approaches to estimating uncertainty.

The frequentist model provides a point estimate of the effect size with confidence intervals, which also widen with smaller sample sizes or higher variance to reflect increased uncertainty. However, these intervals may sometimes underestimate variability arising from differences in study design or data characteristics, potentially inflating confidence in the effect size. Conversely, Bayesian models incorporate prior information, and thus generally produce more cautious estimates with broader credible intervals, especially for studies with lower power (Sadeghirad et al., 2023), which better reflect underlying uncertainty in the data. Conversely, frequentist models provide more definitive estimates of the effect size but may not fully account for the underlying variability in study designs or data characteristics.

It was assumed that there would be a high degree of heterogeneity for both meta-analyses, considering the wide variety of study designs, EEG paradigms and outcome measures. This was confirmed not only via the frequentist multilevel meta-analyses but also via Robust Bayesian meta-analyses. Computing a GOSH plot revealed that heterogeneity varied greatly across study subsets, with wide *I*^2^ and τ² ranges for both meta-analyses, indicating that some studies contribute more to influence heterogeneity than others. The *I*^2^ statistic was computed to elucidate the variance in observed effects: true effects ratio. For the Neural MA, the *I²*-statistics revealed that within-study variability was the largest contributor to overall heterogeneity in the meta-analysis.

While this could be interpreted as being due to sampling error rather than a difference in true effects, most studies in the Neural MA produced a variety of different outcome measures that were not necessarily dependent on each other. Therefore, this could indicate that the within-study heterogeneity reflects a true difference in effects reported from the same studies, compared to the studies in the Social/Cognitive MA, where between-study heterogeneity was higher. This difference may in part be due to all microstate studies (Schiller et al., 2019; Tomescu et al., 2024; Zelenina et al., 2022) including effect sizes from different microstate metrics, such as duration, coverage, contribution and occurrence of the different microstates. Microstates reflect quasi-stable brain map configurations transitioning to different, quasi stable map configurations (Lehmann et al., 1987), which can be more heterogeneous depending on segmentation approaches, which capture different aspects of large-scale neural activity that are not necessarily correlated (Koenig et al., 2024; Michel & Koenig, 2018). However, a post-hoc moderator analysis investigating potential moderating effects of different EEG measurements in the Neural MA did not find any significant moderating effects, nor did it explain residual heterogeneity.

A significant moderator effect of measurements of interest was found in the Social/Cognitive MA, revealing that effect sizes varied systematically based on type of measurement. This explained a substantial portion of the observed variance. Studies in this meta-analysis using symmetry/asymmetry measures reported larger and more positive effects, whereas the studies relying on ERPs, frequency domain analyses, and microstates reported smaller effect sizes in comparison. It is important to note that all these studies used different tasks, the study designs varied, and the study outcomes differed. Moderator analyses on type of outcome (social versus non-social) and task type (behavioural, visual, visual and behavioural) did however not yield any significant results. One cannot draw the conclusion that specific EEG measurements are better suited than others to capture effects of oxytocin administration on neural responses to social or cognitive outcomes. Instead, it may suggest that different EEG measures are more or less sensitive to distinct neurophysiological processes influenced by oxytocin, or that certain methods tend to produce larger or more consistent effect sizes. ERPs showed smaller effect sizes and are often used in task-based designs to measure stimulus-locked brain activity, whereas symmetry/asymmetry measures, which showed larger effects, may reflect more sustained changes in neural activity in response to social or cognitive tasks as these measures capture more stable and large-scale neural patterns. Similarly, microstates and frequency-domain analyses capture different aspects of spontaneous or task-independent neural dynamics, where microstates studies may, as previously outlined, also display more heterogeneity depending on microstate mapping methodologies and measurement metrics (Koenig et al., 2024). In contrast, ERPs and frequency-domain measures may both be more susceptible to variability in preprocessing, trial averaging, and task conditions. Different methods and EEG measurements capture different neural processes, which may lead to variation in effect sizes. The heterogeneity in EEG measures and study tasks make interpretation complex, underscoring the need for careful consideration of methodological choices in EEG research. Future research would benefit from systematic comparison of different EEG measurements within the same experimental paradigms to better determine which measures are the most reliable and sensitive to capture the different effects of oxytocin administration on neural activity.

For the Social/Cognitive MA, the moderator *gender* was significant, where the effect size decreased as the number of female participants increased. The eligible studies in the present meta-analysis primarily included male participants as is typical in the field. In fact, in the Social/Cognitive MA, only 8 studies (Moerkerke et al., 2023; Ochiai et al., 2021; Peltola et al., 2018; Petereit et al., 2019; Rutherford et al., 2017; Schiller et al., 2023; Singh et al., 2016; X. Zhang et al., 2021) included female participants at all, with a total of 47 effect sizes with female participants across these studies. Half of these studies included *both* female and male participants (Moerkerke et al., 2023; Ochiai et al., 2021; Schiller et al., 2023; Singh et al., 2016; X. Zhang et al., 2021). This highlights the need for more studies with female participants, both to improve our understanding of the interplay between oxytocin and sex hormones, and to better understand the mechanisms of oxytocin overall (Quintana et al., 2024). The importance of including both male and female participants to elucidate sex specific effects is further exemplified in Schiller et al. (2023): The authors found specific effects of exogenous oxytocin when investigating neurophysiological correlates of attention in an approach or avoidance scenario. While the percentage of female participants was not a significant moderator in the Neural MA, the effect indicated a slight decrease in effect size when the number of female participants increased. This could potentially support previous findings of oxytocin affecting men and women differently, not only in context-dependent social tasks (Procyshyn et al., 2024), but also in neural responses in resting-state tasks as seen in fMRI studies (e.g. Coenjaerts et al., 2023; Ebner et al., 2016). However, considering the small sample sizes and small effect sizes in this meta-analysis, more research is warranted to investigate sex differences in broader neural activation after oxytocin administration.

While there is evidence that oxytocin receptor expression in the brain fluctuates throughout the lifespan (Audunsdottir & Quintana, 2022) and also plays a role in response to exogenous oxytocin (Ebner et al., 2016; Horta et al., 2023; Huffmeijer et al., 2012), we were not able to detect any moderating effects of age in any of the meta-analyses. However, among the eligible studies, only four tested a population with a mean age outside the range of 20 to 30 years (Alaerts et al., 2024 [*M*_age_ = 10.45]; Moerkerke et al., 2003 [*M*_age_ = 10.1]; Singh et al., 2016 [Healthy participants: *M*_age_ = 37, schizophrenia participants: *M*_age_ = 47]; Peltola et al., 2018, *M*_age_ = 31.9). More research is therefore needed to investigate potential moderating effects of age in a study sample with more diverse ranges.

Neither oxytocin administration dosage (24 IU or more than 24 IU) nor time between administration and testing yielded significant results for either of the meta-analyses. However, sub-group analysis of studies utilising 24 IU reached statistical significance in the Cognitive MA. Interestingly, meta-analysis with studies employing higher dosages (>24IU) had smaller effects on social/non-social outcomes than studies employing 24 IU. This can provide some tentative support for the notion that lower dosages are more efficient than higher dosages (Martins et al., 2022; Quintana et al., 2015, 2017). One of the included studies found dose-dependent effects of oxytocin that were moderated by both gender and diagnosis, where higher doses (48 IU) compared to lower doses (24 IU) led to more beneficial effects in males, while the opposite was true for females. In healthy participants, this dose-response pattern was reversed (Singh et al., 2016).

However, more research is needed to better tell how generalisable this is to different contexts and populations. Conclusions about the most effective dose should also be made with caution due to the wide variation of experimental tasks utilised in the studies, considering oxytocin’s task specificity (Wigton et al., 2015). As this meta-analysis did not include studies using dosages lower than 24 IU, future research should aim to investigate if this could be a more efficient dosage in some populations and across contexts than the commonly used 24 IU.

The eligible studies in this paper tended to use the within-participant design more frequently than the between-participant design (N_within_ = 15, N_between_ = 13). Therefore, potential moderating effects of employing a between-participant or within-participant study design were assessed. While the summary moderator effects did not reach statistical significance for neither of the two meta-analyses, both yielded a significant between-participant moderator variable. This indicates that the effects tended to be larger in studies employing a between-participant design compared to the studies with a within-participant design. Taken together, these findings suggest that the choice of experimental design in oxytocin research may influence the magnitude of observed effects.

We found no evidence of moderating effects of psychiatric conditions or disorders in the Social/Cognitive MA. However, it is important to note that the number of effect sizes from clinical populations was small (N_clinical_ = 5, N_neurotypical_ = 108), so any interpretations should be made with caution. Most studies on clinical populations tend to find beneficial effects on social cognition and behaviour after oxytocin administration (e.g. Anagnostou et al., 2012; Cochran et al., 2013; Guastella et al., 2010, 2015; Ma et al., 2016; Rubin et al., 2014). For example, a study by Singh et al. (2016) suggested that oxytocin administration increases mirror neuron system (MNS) activity. MNS dysfunction may contribute to the social deficits observed in schizophrenia, indicating that oxytocin administration may facilitate neural processes underlying social cognition in individuals with schizophrenia. Conversely, a recent meta-analysis and systematic review by Audunsdottir et al. (2024) found modest and inconclusive support for oxytocin administration’s effect on social outcomes and routinised behaviours respectively, but the authors noted that most of the included studies were statistically underpowered to detect small-to-medium effects. In this paper’s meta-analyses, two of the included studies found inconsistent effects of chronic oxytocin administration in autistic children. Moerkerke et al. (2023) reported reduced neural sensitivity to expressive faces during oxytocin administration. This was interpreted as inhibition of learning effects in neural sensitivity to emotionally evocative faces due to oxytocin’s anxiolytic and stress regulatory effects. Here, the authors found that an increase only occurred after cessation. Alaerts et al. (2024) found a reduction in noisy oscillatory coupling at a follow-up session, which was more similar to brain state patterns seen in non-autistic children, suggesting long lasting modulation of neural architecture even after oxytocin administration cessation. As only two studies used a chronic administration schedule, moderator analyses comparing single and chronic dosing of oxytocin was not computed. There is emerging evidence for different mechanisms of effect for chronic and single doses of oxytocin (Benner et al., 2021; Horta et al., 2020). Chronic administration may target the general stress system as opposed to increased neural activity related to processing and boosting the salience of social cues (S. Zhang et al., 2023). Chronic oxytocin administration is for example suggested to suppress hypothalamic–pituitary–adrenal (HPA) axis activity during stress by modulating receptor expression and binding in stress-related brain regions (Horta et al., 2020). These studies highlight the need to investigate both single dose effects and more chronic, long-term, effects of oxytocin administration. Both the existing literature and our findings suggest that more research is needed to understand oxytocin’s mechanisms of action and before considering oxytocin as a potential individualised support in different populations.

### Potential limitations

While there are limitations to EEG, such as spatial ambiguity especially in deeper brain structures (Michel & Brunet, 2019; Zorzos et al., 2021) and indirect measurements of oxytocin receptor activity (Bewernitz & Derendorf, 2012), advancements in EEG research and source localisation challenge some of these limitations (Fahimi Hnazaee et al., 2020; Jobert & Wilson, 2016; Marino & Mantini, 2024; Michel & Brunet, 2019; Yektaeian Vaziri & Makkiabadi, 2025; Yen et al., 2023). EEG provides millisecond level sensitivity to detect modulations of neural oscillations and connectivity while being entirely non-invasive and cost-effective (Light et al., 2010; Marino & Mantini, 2024). EEG further supports repeated measurements over time to capture both acute and long-term changes (Gevins et al., 2012; Knott et al., 2002; Liu et al., 2024). Investigating neural correlates of oxytocin administration via EEG is still a new and growing field, and as the research literature expands, it would be of interest to comprehensively investigate and compare specific EEG recording specifics (e.g. type of cap or electrodes), preprocessing methods, measurements, paradigms, or outcomes to get a better understanding of the effects of oxytocin. Various concerns have been raised in oxytocin research about different biases (Lane et al., 2016): Pre-registration has been suggested as potential means to reduce researcher biases and remedy some of the selective reporting that has been seen (Hardwicke & Wagenmakers, 2023). Summaries of alternative analyses conducted by independent experts, “synchronous robustness reports”, have also been proposed to boast robustness (Bartoš et al., 2025). Here, publication bias and small study bias were investigated by computing Egger’s regression and funnel plots, where both revealed some evidence for the presence of bias for the meta-analysis on social and non-social outcomes. Furthermore, statistical power medians for both meta-analyses fell within the estimated low median statistical power of neuroscience studies, i.e. in the 8-31% range (Button et al., 2013). This can indicate that in both meta-analyses the studies are – on average – statistically too underpowered to detect a wide range of effect sizes, revealing a risk of exaggerated effect size estimates and bias.

A limitation of this study is the restricted availability of data that was needed to conduct a meta-analysis. Out of all potentially eligible articles from the literature search, descriptive statistics such as means and standard deviations were rarely reported in the main article, or even in the supplementary materials. More available data would increase the number of studies in these meta-analyses, and provide more evidence for, or against, specific effects, and more detailed moderator analyses could be conducted.

## Conclusion

In conclusion, we present evidence of tentative support for oxytocin administration significantly modulating a wide range of neural activity as measured by EEG, compared to placebo. The results from the two meta-analyses indicate that oxytocin administration generally have beneficial effects on neural correlates of cognition and social processing and behaviour, and also significantly modulates EEG measures connected to a wide range of neural mechanisms, brain states and networks. These findings support a growing body of literature pointing to oxytocin’s role in multiple neural processes, where its mechanisms of action largely are context-dependent. However, most studies were statistically underpowered to reliably detect a wide range of effect sizes and observed heterogeneity both within and between studies was high. The variation in tasks, study designs, and EEG paradigms limits the ability to directly compare results across studies and precludes drawing strong conclusions about oxytocin’s specific mechanisms of action - even though several moderator analyses were conducted to explore potential sources of variability. To our knowledge, this is the first study to systematically synthesize findings on the effects of oxytocin administration in on neural activity in humans as measured by EEG. As more research accumulates, it will be important for future meta-analyses to focus on more narrowly defined subsets of studies to clarify context-specific effects and improve comparability. Likewise, continued efforts toward methodological consistency and transparency in experimental studies will further strengthen the field, particularly as important sources of variability remain to be fully explored. Open science practices, including pre-registration and data sharing, when possible, are encouraged to support reproducibility and lower the risk of bias.

## Data Availability

All data produced are available online at https://osf.io/ynsd7/

## Acknowledgements and financial support

The literature search was performed by librarian Skjalg Tønnesen Kalvik at the Medical Library at Rikshospitalet.

This work was supported by the Research Council of Norway (PI DQ: 301767; 324783), South-Eastern Norway Regional Health Authority (PI DQ: 2022059), and the Kavli Trust (PI DQ).

## References

Alaerts, K., Moerkerke, M., Daniels, N., Zhang, Q., Grazia, R., Steyaert, J., Prinsen, J., & Boets, B. (2024). Chronic oxytocin improves neural decoupling at rest in children with autism: An exploratory RCT. Journal of Child Psychology and Psychiatry, 65(10), 1311–1326. 10.1111/jcpp.13966

Alaerts, K., Taillieu, A., Daniels, N., Soriano, J. R., & Prinsen, J. (2021). Oxytocin enhances neural approach towards social and non-social stimuli of high personal relevance. Scientific Reports, 11(1), 23589. 10.1038/s41598-021-02914-8

Anagnostou, E., Soorya, L., Chaplin, W., Bartz, J., Halpern, D., Wasserman, S., Wang, A. T., Pepa, L., Tanel, N., Kushki, A., & Hollander, E. (2012). Intranasal oxytocin versus placebo in the treatment of adults with autism spectrum disorders: A randomized controlled trial. Molecular Autism, 3(1), 16. 10.1186/2040-2392-3-16

Andari, E., Duhamel, J.-R., Zalla, T., Herbrecht, E., Leboyer, M., & Sirigu, A. (2010). Promoting social behavior with oxytocin in high-functioning autism spectrum disorders. Proceedings of the National Academy of Sciences, 107(9), 4389–4394. 10.1073/pnas.0910249107

Audunsdottir, K., & Quintana, D. S. (2022). Oxytocin’s dynamic role across the lifespan. Aging Brain, 2, 100028. 10.1016/j.nbas.2021.100028

Audunsdottir, K., Sartorius, A. M., Kang, H., Glaser, B. D., Boen, R., Nærland, T., Alaerts, K., Kildal, E. S. M., Westlye, L. T., Andreassen, O. A., & Quintana, D. S. (2024). The effects of oxytocin administration on social and routinized behaviors in autism: A preregistered systematic review and meta-analysis. Psychoneuroendocrinology, 167, 107067. 10.1016/j.psyneuen.2024.107067

Auyeung, B., Lombardo, M. V., Heinrichs, M., Chakrabarti, B., Sule, A., Deakin, J. B., Bethlehem, R. a. I., Dickens, L., Mooney, N., Sipple, J. a. N., Thiemann, P., & Baron-Cohen, S. (2015). Oxytocin increases eye contact during a real-time, naturalistic social interaction in males with and without autism. Translational Psychiatry, 5(2), e507–e507. 10.1038/tp.2014.146

Bakos, J., Srancikova, A., Havranek, T., & Bacova, Z. (2018). Molecular Mechanisms of Oxytocin Signaling at the Synaptic Connection. Neural Plasticity, 2018(1), 4864107. 10.1155/2018/4864107

Bartoš, F., Sarafoglou, A., Aczel, B., Hoogeveen, S., Chambers, C. D., & Wagenmakers, E.-J. (2025). Introducing synchronous robustness reports. Nature Human Behaviour, 9(4), 635–637. 10.1038/s41562-025-02129-1

Bartz, J. A., Zaki, J., Bolger, N., Hollander, E., Ludwig, N. N., Kolevzon, A., & Ochsner, K. N. (2010). Oxytocin Selectively Improves Empathic Accuracy. Psychological Science, 21(10), 1426–1428. 10.1177/0956797610383439

Başar, E., Başar-Eroğlu, C., Karakaş, S., & Schürmann, M. (2000). Brain oscillations in perception and memory. International Journal of Psychophysiology, 35(2), 95–124. 10.1016/S0167-8760(99)00047-1

Benner, S., Aoki, Y., Watanabe, T., Endo, N., Abe, O., Kuroda, M., Kuwabara, H., Kawakubo, Y., Takao, H., Kunimatsu, A., Kasai, K., Bito, H., Kakeyama, M., & Yamasue, H. (2021). Neurochemical evidence for differential effects of acute and repeated oxytocin administration. Molecular Psychiatry, 26(2), 710–720. 10.1038/s41380-018-0249-4

Bethlehem, R. A. I., van Honk, J., Auyeung, B., & Baron-Cohen, S. (2013). Oxytocin, brain physiology, and functional connectivity: A review of intranasal oxytocin fMRI studies. Psychoneuroendocrinology, 38(7), 962–974. 10.1016/j.psyneuen.2012.10.011

Bewernitz, M., & Derendorf, H. (2012). Electroencephalogram-based pharmacodynamic measures: A review. International Journal of Clinical Pharmacology and Therapeutics, 50(3), 162–184. 10.5414/cp201484

Button, K. S., Ioannidis, J. P. A., Mokrysz, C., Nosek, B. A., Flint, J., Robinson, E. S. J., & Munafò, M. R. (2013). Power failure: Why small sample size undermines the reliability of neuroscience. Nature Reviews Neuroscience, 14(5), 365–376. 10.1038/nrn3475

Caldwell, A. R. (2022). Exploring Equivalence Testing with the Updated TOSTER R Package. PsyArXiv. 10.31234/osf.io/ty8de

Carcea, I., Caraballo, N. L., Marlin, B. J., Ooyama, R., Riceberg, J. S., Mendoza Navarro, J. M., Opendak, M., Diaz, V. E., Schuster, L., Alvarado Torres, M. I., Lethin, H., Ramos, D., Minder, J., Mendoza, S. L., Bair-Marshall, C. J., Samadjopoulos, G. H., Hidema, S., Falkner, A., Lin, D.,… Froemke, R. C. (2021). Oxytocin neurons enable social transmission of maternal behaviour. Nature, 596(7873), 553–557. 10.1038/s41586-021-03814-7

Cheung, M. (2013). Modeling Dependent Effect Sizes With Three-Level Meta-Analyses: A Structural Equation Modeling Approach. Psychological Methods, 19. 10.1037/a0032968

Cochran, D. M., Fallon, D., Hill, M., & Frazier, J. A. (2013). The Role of Oxytocin in Psychiatric Disorders: A Review of Biological and Therapeutic Research Findings. Harvard Review of Psychiatry, 21(5), 219. 10.1097/HRP.0b013e3182a75b7d

Coenjaerts, M., Adrovic, B., Trimborn, I., Philipsen, A., Hurlemann, R., & Scheele, D. (2023). Effects of exogenous oxytocin and estradiol on resting-state functional connectivity in women and men. Scientific Reports, 13(1), 3113. 10.1038/s41598-023-29754-y

Cohen, M. X. (2017). Where Does EEG Come From and What Does It Mean? Trends in Neurosciences, 40(4), 208–218. 10.1016/j.tins.2017.02.004

Daughters, K., Manstead, A. S. R., Ten Velden, F. S., & De Dreu, C. K. W. (2017). Oxytocin modulates third-party sanctioning of selfish and generous behavior within and between groups. Psychoneuroendocrinology, 77, 18–24. 10.1016/j.psyneuen.2016.11.039

Declerck, C. H., Boone, C., Pauwels, L., Vogt, B., & Fehr, E. (2020). A registered replication study on oxytocin and trust. Nature Human Behaviour, 4(6), 646–655. 10.1038/s41562-020-0878-x

Dumais, K. M., & Veenema, A. H. (2016). Vasopressin and oxytocin receptor systems in the brain: Sex differences and sex-specific regulation of social behavior. Frontiers in Neuroendocrinology, 40, 1–23. 10.1016/j.yfrne.2015.04.003

Ebner, N. C., Chen, H., Porges, E., Lin, T., Fischer, H., Feifel, D., & Cohen, R. A. (2016). Oxytocin’s effect on resting-state functional connectivity varies by age and sex. Psychoneuroendocrinology, 69, 50–59. 10.1016/j.psyneuen.2016.03.013

Egger, M., Smith, G. D., Schneider, M., & Minder, C. (1997). Bias in meta-analysis detected by a simple, graphical test. BMJ, 315(7109), 629. 10.1136/bmj.315.7109.629

Ekstrom, A. (2010). How and when the fMRI BOLD signal relates to underlying neural activity: The danger in dissociation. Brain Research Reviews, 62(2), 233–244. 10.1016/j.brainresrev.2009.12.004

Fahimi Hnazaee, M., Wittevrongel, B., Khachatryan, E., Libert, A., Carrette, E., Dauwe, I., Meurs, A., Boon, P., Van Roost, D., & Van Hulle, M. M. (2020). Localization of deep brain activity with scalp and subdural EEG. NeuroImage, 223, 117344. 10.1016/j.neuroimage.2020.117344

Fisher, E. R., Rocha, N. P., Morales-Scheihing, D. A., Venna, V. R., Furr-Stimming, E. E., Teixeira, A. L., & Rossetti, M. A. (2021). The Relationship Between Plasma Oxytocin and Executive Functioning in Huntington’s Disease: A Pilot Study. Journal of Huntington’s Disease, 10(3), 349–354. 10.3233/JHD-210467

Fraley, C., & and Raftery, A. E. (2002). Model-Based Clustering, Discriminant Analysis, and Density Estimation. Journal of the American Statistical Association, 97(458), 611–631. 10.1198/016214502760047131

Fu, R., Gartlehner, G., Grant, M., Shamliyan, T., Sedrakyan, A., Wilt, T. J., Griffith, L., Oremus, M., Raina, P., Ismaila, A., Santaguida, P., Lau, J., & Trikalinos, T. A. (2011). Conducting quantitative synthesis when comparing medical interventions: AHRQ and the Effective Health Care Program. Journal of Clinical Epidemiology, 64(11), 1187–1197. 10.1016/j.jclinepi.2010.08.010

Gevins, A., McEvoy, L. K., Smith, M. E., Chan, C. S., Sam-Vargas, L., Baum, C., & Ilan, A. B. (2012). Long-term and within-day variability of working memory performance and EEG in individuals. Clinical Neurophysiology, 123(7), 1291–1299. 10.1016/j.clinph.2011.11.004

Gibbons, R. D., Hedeker, D. R., & Davis, J. M. (1993). Estimation of Effect Size from a Series of Experiments Involving Paired Comparisons. Journal of Educational Statistics, 18(3), 271–279. JSTOR. 10.2307/1165136

Gimpl, G., & Fahrenholz, F. (2001). The Oxytocin Receptor System: Structure, Function, and Regulation. Physiological Reviews, 81(2), 629–683. 10.1152/physrev.2001.81.2.629

Gkintoni, E., Aroutzidis, A., Antonopoulou, H., & Halkiopoulos, C. (2025). From Neural Networks to Emotional Networks: A Systematic Review of EEG-Based Emotion Recognition in Cognitive Neuroscience and Real-World Applications. Brain Sciences, 15(3). 10.3390/brainsci15030220

Glover, G. H. (2011). Overview of Functional Magnetic Resonance Imaging. Functional Imaging, 22(2), 133–139. 10.1016/j.nec.2010.11.001

Grace, S. A., Rossell, S. L., Heinrichs, M., Kordsachia, C., & Labuschagne, I. (2018). Oxytocin and brain activity in humans: A systematic review and coordinate-based meta-analysis of functional MRI studies. Psychoneuroendocrinology, 96, 6–24. 10.1016/j.psyneuen.2018.05.031

Grinevich, V., Desarménien, M. G., Chini, B., Tauber, M., & Muscatelli, F. (2015). Ontogenesis of oxytocin pathways in the mammalian brain: Late maturation and psychosocial disorders. Frontiers in Neuroanatomy, 8. 10.3389/fnana.2014.00164

Guastella, A. J., Einfeld, S. L., Gray, K. M., Rinehart, N. J., Tonge, B. J., Lambert, T. J., & Hickie, I. B. (2010). Intranasal Oxytocin Improves Emotion Recognition for Youth with Autism Spectrum Disorders. Biological Psychiatry, 67(7), 692–694. 10.1016/j.biopsych.2009.09.020

Guastella, A. J., Ward, P. B., Hickie, I. B., Shahrestani, S., Hodge, M. A. R., Scott, E. M., & Langdon, R. (2015). A single dose of oxytocin nasal spray improves higher-order social cognition in schizophrenia. Schizophrenia Research, 168(3), 628–633. 10.1016/j.schres.2015.06.005

Hardwicke, T. E., & Wagenmakers, E.-J. (2023). Reducing bias, increasing transparency and calibrating confidence with preregistration. Nature Human Behaviour, 7(1), 15–26. 10.1038/s41562-022-01497-2

Harrer, M., Cuijpers, P., Furukawa, T. A., & Ebert, D. D. (2021). Doing Meta-Analysis With R: A Hands-On Guide (1st ed.). Chapman & Hall/CRC Press. https://www.routledge.com/Doing-Meta-Analysis-with-R-A-Hands-On-Guide/Harrer-Cuijpers-Furukawa-Ebert/p/book/9780367610074

Harrer, M., Cuijpers, P., Furukawa, T., & Ebert, D. D. (2019). dmetar: Companion R Package For The Guide’Doing Meta-Analysis in R (Version R package version 0.1.0) [Computer software]. http://dmetar.protectlab.org/

Hartigan, J. A., & Wong, M. A. (1979). A K-Means Clustering Algorithm. Journal of the Royal Statistical Society Series C: Applied Statistics, 28(1), 100–108. 10.2307/2346830

Hedges, L. V. (1981). Distribution Theory for Glass’s Estimator of Effect Size and Related Estimators. Journal of Educational Statistics, 6(2), 107–128. JSTOR. 10.2307/1164588

Hollander, E., Bartz, J., Chaplin, W., Phillips, A., Sumner, J., Soorya, L., Anagnostou, E., & Wasserman, S. (2007). Oxytocin Increases Retention of Social Cognition in Autism. Biological Psychiatry, 61(4), 498–503. 10.1016/j.biopsych.2006.05.030

Holmes, G. L., & Khazipov, R. (2007). Basic Neurophysiology and the Cortical Basis of EEG. In A. S. Blum & S. B. Rutkove (Eds.), The Clinical Neurophysiology Primer (pp. 19–33). Humana Press. 10.1007/978-1-59745-271-7_2

Horta, M., Kaylor, K., Feifel, D., & Ebner, N. C. (2020). Chronic oxytocin administration as a tool for investigation and treatment: A cross-disciplinary systematic review. Neuroscience & Biobehavioral Reviews, 108, 1–23. 10.1016/j.neubiorev.2019.10.012

Horta, M., Polk, R., & Ebner, N. C. (2023). Single dose intranasal oxytocin administration: Data from healthy younger and older adults. Data in Brief, 51, 109669. 10.1016/j.dib.2023.109669

Huettel, S., Song, A., & McCarthy, G. (2008). Functional Magnetic Resonance Imaging, Second Edition.

Huffmeijer, R., van IJzendoorn, M. H., & Bakermans-Kranenburg, M. J. (2012). Ageing and Oxytocin: A Call for Extending Human Oxytocin Research to Ageing Populations – A Mini-Review. Gerontology, 59(1), 32–39. 10.1159/000341333

Jackson, A. F., & Bolger, D. J. (2014). The neurophysiological bases of EEG and EEG measurement: A review for the rest of us. Psychophysiology, 51(11), 1061–1071. 10.1111/psyp.12283

Jobert, M., & Wilson, F. J. (2016). Advanced Analysis of Pharmaco-EEG Data in Humans. Neuropsychobiology, 72(3–4), 165–177. 10.1159/000431096

Jurek, B., & Neumann, I. D. (2018). The Oxytocin Receptor: From Intracellular Signaling to Behavior. Physiological Reviews, 98(3), 1805–1908. 10.1152/physrev.00031.2017

Kang, H., Glaser, B. D., Sartorius, A. I., Audunsdottir, K., Kildal, E. S.-M., Nærland, T., Andreassen, O. A., Westlye, L. T., & Quintana, D. S. (2025). Effects of oxytocin administration on non-social executive functions in humans: A preregistered systematic review and meta-analysis. Molecular Psychiatry, 30(5), 2239–2251. 10.1038/s41380-024-02871-4

Keech, B., Crowe, S., & Hocking, D. R. (2018). Intranasal oxytocin, social cognition and neurodevelopmental disorders: A meta-analysis. Psychoneuroendocrinology, 87, 9–19. 10.1016/j.psyneuen.2017.09.022

Kirschstein, T., & Köhling, R. (2009). What is the Source of the EEG? Clinical EEG and Neuroscience, 40(3), 146–149. 10.1177/155005940904000305

Klopfer, P. H., & Klopfer, M. S. (1968). Maternal “Imprinting” in Goats: Fostering of alien young. Zeitschrift Für Tierpsychologie, 25(7), 862–866. 10.1111/j.1439-0310.1968.tb00048.x

Knott, V., Mahoney, C., Kennedy, S., & Evans, K. (2002). EEG correlates of acute and chronic paroxetine treatment in depression. Journal of Affective Disorders, 69(1), 241–249. 10.1016/S0165-0327(01)00308-1

Koenig, T., Diezig, S., Kalburgi, S. N., Antonova, E., Artoni, F., Brechet, L., Britz, J., Croce, P., Custo, A., Damborská, A., Deolindo, C., Heinrichs, M., Kleinert, T., Liang, Z., Murphy, M. M., Nash, K., Nehaniv, C., Schiller, B., Smailovic, U.,… Michel, C. M. (2024). EEG-Meta-Microstates: Towards a More Objective Use of Resting-State EEG Microstate Findings Across Studies. Brain Topography, 37(2), 218–231. 10.1007/s10548-023-00993-6

Kosfeld, M., Heinrichs, M., Zak, P. J., Fischbacher, U., & Fehr, E. (2005). Oxytocin increases trust in humans. Nature, 435(7042), 673–676. 10.1038/nature03701

Lakens, D. (2017). Equivalence Tests: A Practical Primer for t Tests, Correlations, and Meta-Analyses. Social Psychological and Personality Science, 8(4), 355–362. 10.1177/1948550617697177

Lambert, B., Declerck, C. H., Boone, C., & Parizel, P. M. (2017). A functional MRI study on how oxytocin affects decision making in social dilemmas: Cooperate as long as it pays off, aggress only when you think you can win. Hormones and Behavior, 94, 145–152. 10.1016/j.yhbeh.2017.06.011

Lane, A., Luminet, O., Nave, G., & Mikolajczak, M. (2016). Is there a Publication Bias in Behavioural Intranasal Oxytocin Research on Humans? Opening the File Drawer of One Laboratory. Journal of Neuroendocrinology, 28(4). 10.1111/jne.12384

Lehmann, D., Ozaki, H., & Pal, I. (1987). EEG alpha map series: Brain micro-states by space-oriented adaptive segmentation. Electroencephalography and Clinical Neurophysiology, 67(3), 271–288. 10.1016/0013-4694(87)90025-3

Leng, G., & Leng, R. I. (2021). Oxytocin: A citation network analysis of 10 000 papers. Journal of Neuroendocrinology, 33(11), e13014. 10.1111/jne.13014

Light, G. A., Williams, L. E., Minow, F., Sprock, J., Rissling, A., Sharp, R., Swerdlow, N. R., & Braff, D. L. (2010). Electroencephalography (EEG) and Event-Related Potentials (ERPs) with Human Participants. Current Protocols in Neuroscience, 52(1), 6.25.1-6.25.24. 10.1002/0471142301.ns0625s52

Liu, Q., Jia, S., Tu, N., Zhao, T., Lyu, Q., Liu, Y., Song, X., Wang, S., Zhang, W., Xiong, F., Zhang, H., Guo, Y., & Wang, G. (2024). Open access EEG dataset of repeated measurements from a single subject for microstate analysis. Scientific Data, 11(1), 379. 10.1038/s41597-024-03241-z

Logothetis, N. K. (2008). What we can do and what we cannot do with fMRI. Nature, 453(7197), 869–878. 10.1038/nature06976

Ma, Y., Shamay-Tsoory, S., Han, S., & Zink, C. F. (2016). Oxytocin and Social Adaptation: Insights from Neuroimaging Studies of Healthy and Clinical Populations. Trends in Cognitive Sciences, 20(2), 133–145. 10.1016/j.tics.2015.10.009

Marino, M., & Mantini, D. (2024). Human brain imaging with high-density electroencephalography: Techniques and applications. The Journal of Physiology, n/a(n/a). 10.1113/JP286639

Marlin, B. J., Mitre, M., D’amour, J. A., Chao, M. V., & Froemke, R. C. (2015). Oxytocin enables maternal behaviour by balancing cortical inhibition. Nature, 520(7548), 499–504. 10.1038/nature14402

Marshall, A. D., Occhipinti, S., & Loxton, N. J. (2024). Tipping the analytical scales, investigating the use of frequentist equivalence analyses in psychology: A scoping review. Quality & Quantity, 58(3), 2929–2955. 10.1007/s11135-023-01758-w

Martins, D., Brodmann, K., Veronese, M., Dipasquale, O., Mazibuko, N., Schuschnig, U., Zelaya, F., Fotopoulou, A., & Paloyelis, Y. (2022). “Less is more”: A dose-response account of intranasal oxytocin pharmacodynamics in the human brain. Progress in Neurobiology, 211, 102239. 10.1016/j.pneurobio.2022.102239

McCall, C., & Singer, T. (2012). The animal and human neuroendocrinology of social cognition, motivation and behavior. Nature Neuroscience, 15(5), 681–688. 10.1038/nn.3084

Meyer-Lindenberg, A., Domes, G., Kirsch, P., & Heinrichs, M. (2011). Oxytocin and vasopressin in the human brain: Social neuropeptides for translational medicine. Nature Reviews Neuroscience, 12(9), 524–538. 10.1038/nrn3044

Michalopoulou, P. G., Averbeck, B. B., Kalpakidou, A. K., Evans, S., Bobin, T., Kapur, S., & Shergill, S. S. (2015). The effects of a single dose of oxytocin on working memory in schizophrenia. Schizophrenia Research, 162(1), 62–63. 10.1016/j.schres.2014.12.029

Michel, C. M., & Brunet, D. (2019). EEG Source Imaging: A Practical Review of the Analysis Steps. Frontiers in Neurology, 10. 10.3389/fneur.2019.00325

Michel, C. M., & Koenig, T. (2018). EEG microstates as a tool for studying the temporal dynamics of whole-brain neuronal networks: A review. Brain Connectivity Dynamics, 180, 577–593. 10.1016/j.neuroimage.2017.11.062

Michel, C. M., & Murray, M. M. (2012). Towards the utilization of EEG as a brain imaging tool. NeuroImage, 61(2), 371–385. 10.1016/j.neuroimage.2011.12.039

Mierop, A., Mikolajczak, M., Stahl, C., Béna, J., Luminet, O., Lane, A., & Corneille, O. (2020). How Can Intranasal Oxytocin Research Be Trusted? A Systematic Review of the Interactive Effects of Intranasal Oxytocin on Psychosocial Outcomes. Perspectives on Psychological Science, 15(5), 1228–1242. 10.1177/1745691620921525

Mitre, M., Minder, J., Morina, E. X., Chao, M. V., & Froemke, R. C. (2018). Oxytocin Modulation of Neural Circuits. In R. Hurlemann & V. Grinevich (Eds.), Behavioral Pharmacology of Neuropeptides: Oxytocin (pp. 31–53). Springer International Publishing. 10.1007/7854_2017_7

Moerkerke, M., Daniels, N., Van der Donck, S., Tibermont, L., Tang, T., Debbaut, E., Bamps, A., Prinsen, J., Steyaert, J., Alaerts, K., & Boets, B. (2023). Can repeated intranasal oxytocin administration affect reduced neural sensitivity towards expressive faces in autism? A randomized controlled trial. Journal of Child Psychology and Psychiatry, 64(11), 1583–1595. 10.1111/jcpp.13850

Niedermeyer, E. (2005). Electroencephalography: Basic principles, clinical applications, and related fields (Fifth edition.). Lippincott Williams & Wilkins.

Ochiai, H., Shiga, T., Hoshino, H., Horikoshi, S., Kanno, K., Wada, T., Osakabe, Y., Miura, I., & Yabe, H. (2021). Effect of oxytocin nasal spray on auditory automatic discrimination measured by mismatch negativity. Psychopharmacology, 238(7), 1781–1789. 10.1007/s00213-021-05807-w

Olkin, I., Dahabreh, I. J., & Trikalinos, T. A. (2012). GOSH – a graphical display of study heterogeneity. Research Synthesis Methods, 3(3), 214–223. 10.1002/jrsm.1053

Page, M. J., McKenzie, J. E., Bossuyt, P. M., Boutron, I., Hoffmann, T. C., Mulrow, C. D., Shamseer, L., Tetzlaff, J. M., Akl, E. A., Brennan, S. E., Chou, R., Glanville, J., Grimshaw, J. M., Hróbjartsson, A., Lalu, M. M., Li, T., Loder, E. W., Mayo-Wilson, E., McDonald, S.,… Moher, D. (2021). The PRISMA 2020 statement: An updated guideline for reporting systematic reviews. BMJ, 372, n71. 10.1136/bmj.n71

Peltola, M. J., Strathearn, L., & Puura, K. (2018). Oxytocin promotes face-sensitive neural responses to infant and adult faces in mothers. Psychoneuroendocrinology, 91, 261–270. 10.1016/j.psyneuen.2018.02.012

Perry, A., Bentin, S., Shalev, I., Israel, S., Uzefovsky, F., Bar-On, D., & Ebstein, R. P. (2010). Intranasal oxytocin modulates EEG mu/alpha and beta rhythms during perception of biological motion. Psychoneuroendocrinology, 35(10), 1446–1453. 10.1016/j.psyneuen.2010.04.011

Petereit, P., Rinn, C., Stemmler, G., & Mueller, E. M. (2019). Oxytocin reduces the link between neural and affective responses after social exclusion. Biological Psychology, 145, 224–235. 10.1016/j.biopsycho.2019.05.002

Peters, J. L., Sutton, A. J., Jones, D. R., Abrams, K. R., & Rushton, L. (2008). Contour-enhanced meta-analysis funnel plots help distinguish publication bias from other causes of asymmetry. Journal of Clinical Epidemiology, 61(10), 991–996. 10.1016/j.jclinepi.2007.11.010

Posit team. (2025). RStudio: Integrated Development Environment for R [Computer software]. Posit Software, PBC. http://www.posit.co/

Procyshyn, T. L., Dupertuys, J., & Bartz, J. A. (2024). Neuroimaging and behavioral evidence of sex-specific effects of oxytocin on human sociality. Trends in Cognitive Sciences, 28(10), 948– 961. 10.1016/j.tics.2024.06.010

Quintana, D. S. (2018). Revisiting non-significant effects of intranasal oxytocin using equivalence testing. Psychoneuroendocrinology, 87, 127–130. 10.1016/j.psyneuen.2017.10.010

Quintana, D. S. (2023). A Guide for Calculating Study-Level Statistical Power for Meta-Analyses. Advances in Methods and Practices in Psychological Science, 6(1), 25152459221147260. 10.1177/25152459221147260

Quintana, D. S., Glaser, B. D., Kang, H., Kildal, E. S. M., Audunsdottir, K., Sartorius, A. M., & Barth, C. (2024). The interplay of oxytocin and sex hormones. Neuroscience & Biobehavioral Reviews, 163, 105765. 10.1016/j.neubiorev.2024.105765

Quintana, D. S., & Guastella, A. J. (2020). An Allostatic Theory of Oxytocin. Trends in Cognitive Sciences, 24(7), 515–528. 10.1016/j.tics.2020.03.008

Quintana, D. S., Lischke, A., Grace, S., Scheele, D., Ma, Y., & Becker, B. (2021). Advances in the field of intranasal oxytocin research: Lessons learned and future directions for clinical research. Molecular Psychiatry, 26(1), 80–91. 10.1038/s41380-020-00864-7

Quintana, D. S., Outhred, T., Westlye, L. T., Malhi, G. S., & Andreassen, O. A. (2016). The impact of oxytocin administration on brain activity: A systematic review and meta-analysis protocol. Systematic Reviews, 5(1), 205. 10.1186/s13643-016-0386-2

Quintana, D. S., Westlye, L. T., Hope, S., Nærland, T., Elvsåshagen, T., Dørum, E., Rustan, Ø., Valstad, M., Rezvaya, L., Lishaugen, H., Stensønes, E., Yaqub, S., Smerud, K. T., Mahmoud, R. A., Djupesland, P. G., & Andreassen, O. A. (2017). Dose-dependent social-cognitive effects of intranasal oxytocin delivered with novel Breath Powered device in adults with autism spectrum disorder: A randomized placebo-controlled double-blind crossover trial. Translational Psychiatry, 7(5), e1136–e1136. 10.1038/tp.2017.103

Quintana, D. S., Westlye, L. T., Rustan, Ø. G., Tesli, N., Poppy, C. L., Smevik, H., Tesli, M., Røine, M., Mahmoud, R. A., Smerud, K. T., Djupesland, P. G., & Andreassen, O. A. (2015). Low-dose oxytocin delivered intranasally with Breath Powered device affects social-cognitive behavior: A randomized four-way crossover trial with nasal cavity dimension assessment. Translational Psychiatry, 5(7), e602–e602. 10.1038/tp.2015.93

R Core Team. (2024). R: A Language and Environment for Statistical Computing [Computer software]. R Foundation for Statistical Computing. https://www.R-project.org

Roberts, B. Z., Young, J. W., He, Y. V., Cope, Z. A., Shilling, P. D., & Feifel, D. (2019). Oxytocin improves probabilistic reversal learning but not effortful motivation in Brown Norway rats. Neuropharmacology, 150, 15–26. 10.1016/j.neuropharm.2019.02.028

Rohatgi, A. (2022). WebPlotDigitizer (Version 5.2) [Computer software]. https://automeris.io

Rossion, B., Joyce, C. A., Cottrell, G. W., & Tarr, M. J. (2003). Early lateralization and orientation tuning for face, word, and object processing in the visual cortex. NeuroImage, 20(3), 1609–1624. 10.1016/j.neuroimage.2003.07.010

Rubin, L. H., Carter, C. S., Bishop, J. R., Pournajafi-Nazarloo, H., Drogos, L. L., Hill, S. K., Ruocco, A. C., Keedy, S. K., Reilly, J. L., Keshavan, M. S., Pearlson, G. D., Tamminga, C. A., Gershon, E. S., & Sweeney, J. A. (2014). Reduced Levels of Vasopressin and Reduced Behavioral Modulation of Oxytocin in Psychotic Disorders. Schizophrenia Bulletin, 40(6), 1374–1384. 10.1093/schbul/sbu027

Russell, J., Maguire, S., Hunt, G. E., Kesby, A., Suraev, A., Stuart, J., Booth, J., & McGregor, I. S. (2018). Intranasal oxytocin in the treatment of anorexia nervosa: Randomized controlled trial during re-feeding. Psychoneuroendocrinology, 87, 83–92. 10.1016/j.psyneuen.2017.10.014

Rutherford, H. J. V., Guo, X. M., Graber, K. M., Hayes, N. J., Pelphrey, K. A., & Mayes, L. C. (2017). Intranasal oxytocin and the neural correlates of infant face processing in non-parent women. Biological Psychology, 129, 45–48. 10.1016/j.biopsycho.2017.08.002

Sadeghirad, B., Foroutan, F., Zoratti, M. J., Busse, J. W., Brignardello-Petersen, R., Guyatt, G., & Thabane, L. (2023). Theory and practice of Bayesian and frequentist frameworks for network meta-analysis. BMJ Evidence-Based Medicine, 28(3), 204. 10.1136/bmjebm-2022-111928

Schiller, B., Brustkern, J., Walker, M., Hamm, A., & Heinrichs, M. (2023). Oxytocin has sex-specific effects on trust and underlying neurophysiological processes. Psychoneuroendocrinology, 151, 106076. 10.1016/j.psyneuen.2023.106076

Schiller, B., Koenig, T., & Heinrichs, M. (2019). Oxytocin modulates the temporal dynamics of resting EEG networks. Scientific Reports, 9(1), 13418. 10.1038/s41598-019-49636-6

Schindler, S., Bruchmann, M., & Straube, T. (2023). Beyond facial expressions: A systematic review on effects of emotional relevance of faces on the N170. Neuroscience & Biobehavioral Reviews, 153, 105399. 10.1016/j.neubiorev.2023.105399

Schubert, E., Sander, J., Ester, M., Kriegel, H., & Xu, X. (2017). DBSCAN revisited, revisited: Why and how you should (still) use DBSCAN. ACM Transactions on Database Systems, 42, 1–21. 10.1145/3068335

Singh, F., Nunag, J., Muldoon, G., Cadenhead, K. S., Pineda, J. A., & Feifel, D. (2016). Effects of intranasal oxytocin on neural processing within a socially relevant neural circuit. European Neuropsychopharmacology, 26(3), 626–630. 10.1016/j.euroneuro.2015.12.026

Sippel, L. M., Allington, C. E., Pietrzak, R. H., Harpaz-Rotem, I., Mayes, L. C., & Olff, M. (2017). Oxytocin and Stress-related Disorders: Neurobiological Mechanisms and Treatment Opportunities. Chronic Stress, 1, 2470547016687996. 10.1177/2470547016687996

Smith, E. E., Reznik, S. J., Stewart, J. L., & Allen, J. J. B. (2017). Assessing and conceptualizing frontal EEG asymmetry: An updated primer on recording, processing, analyzing, and interpreting frontal alpha asymmetry. Rigor and Replication: Towards Improved Best Practices in Psychophysiological Research, 111, 98–114. 10.1016/j.ijpsycho.2016.11.005

Sun, Y., Wang, X., Chen, Y., Luan, Z., & Hao, R. (2025). The impact of exogenous Oxytocin on visual cortex plasticity across different stages of visual development. Scientific Reports, 15(1), 12137. 10.1038/s41598-025-96573-8

Ten Velden, F. S., Daughters, K., & De Dreu, C. K. W. (2017). Oxytocin promotes intuitive rather than deliberated cooperation with the in-group. Hormones and Behavior, 92, 164–171. 10.1016/j.yhbeh.2016.06.005

Tomescu, M. I., Van der Donck, S., Perisanu, E. M., Berceanu, A. I., Alaerts, K., Boets, B., & Carcea, I. (2024). Social functioning predicts individual changes in EEG microstates following intranasal oxytocin administration: A double-blind, cross-over randomized clinical trial. Psychophysiology, 61(8), e14581. 10.1111/psyp.14581

van den Pol, A. N. (2012). Neuropeptide Transmission in Brain Circuits. Neuron, 76(1), 98–115. 10.1016/j.neuron.2012.09.014

Viechtbauer, W. (2010). Conducting Meta-Analyses in R with the metafor Package. Journal of Statistical Software, 36(3), 1–48. 10.18637/jss.v036.i03

Viechtbauer, W., & Cheung, M. W.-L. (2010). Outlier and influence diagnostics for meta-analysis. Research Synthesis Methods, 1(2), 112–125. 10.1002/jrsm.11

Walum, H., Waldman, I. D., & Young, L. J. (2016). Statistical and Methodological Considerations for the Interpretation of Intranasal Oxytocin Studies. Oxytocin and Psychiatry: From DNA to Social Behavior, 79(3), 251–257. 10.1016/j.biopsych.2015.06.016

Weisman, O., & Feldman, R. (2013). Oxytocin administration affects the production of multiple hormones. Psychoneuroendocrinology, 38(5), 626–627. 10.1016/j.psyneuen.2013.03.004

Wickham, H. (2016). ggplot2: Elegant Graphics for Data Analysis. Springer-Verlag New York. https://ggplot2.tidyverse.org

Wigton, R., Radua, J., Allen, P., Averbeck, B., Meyer-Lindenberg, A., McGuire, P., Shergill, S. S., & Fusar-Poli, P. (2015). Neurophysiological effects of acute oxytocin administration: Systematic review and meta-analysis of placebo-controlled imaging studies. Journal of Psychiatry and Neuroscience, 40(1), E1. 10.1503/jpn.130289

Williams, T. D. (2007). Individual variation in endocrine systems: Moving beyond the ‘tyranny of the Golden Mean’. Philosophical Transactions of the Royal Society B: Biological Sciences, 363(1497), 1687–1698. 10.1098/rstb.2007.0003

Winterton, A., Westlye, L. T., Steen, N. E., Andreassen, O. A., & Quintana, D. S. (2021). Improving the precision of intranasal oxytocin research. Nature Human Behaviour, 5(1), 9–18. 10.1038/s41562-020-00996-4

Yektaeian Vaziri, A., & Makkiabadi, B. (2025). Accelerated algorithms for source orientation detection and spatiotemporal LCMV beamforming in EEG source localization. Frontiers in Neuroscience, 18. 10.3389/fnins.2024.1505017

Yen, C., Lin, C.-L., & Chiang, M.-C. (2023). Exploring the Frontiers of Neuroimaging: A Review of Recent Advances in Understanding Brain Functioning and Disorders. Life, 13(7), Article 7. 10.3390/life13071472

Zelenina, M., Kosilo, M., da Cruz, J., Antunes, M., Figueiredo, P., Mehta, M. A., & Prata, D. (2022). Temporal dynamics of intranasal oxytocin in human brain electrophysiology. Cerebral Cortex, 32(14), 3110–3126. 10.1093/cercor/bhab404

Zhang, H., Zhou, Q.-Q., Chen, H., Hu, X.-Q., Li, W.-G., Bai, Y., Han, J.-X., Wang, Y., Liang, Z.-H., Chen, D., Cong, F.-Y., Yan, J.-Q., & Li, X.-L. (2023). The applied principles of EEG analysis methods in neuroscience and clinical neurology. Military Medical Research, 10(1), 67. 10.1186/s40779-023-00502-7

Zhang, S., Zhang, Y.-D., Shi, D.-D., & Wang, Z. (2023). Therapeutic uses of oxytocin in stress-related neuropsychiatric disorders. Cell & Bioscience, 13(1), 216. 10.1186/s13578-023-01173-6

Zhang, X., Li, P., Otieno, S. C. S. A., Li, H., & Leppänen, P. H. T. (2021). Oxytocin reduces romantic rejection-induced pain in online speed-dating as revealed by decreased frontal-midline theta oscillations. Psychoneuroendocrinology, 133, 105411. 10.1016/j.psyneuen.2021.105411

Zorzos, I., Kakkos, I., Ventouras, E. M., & Matsopoulos, G. K. (2021). Advances in Electrical Source Imaging: A Review of the Current Approaches, Applications and Challenges. Signals, 2(3), Article 3. 10.3390/signals2030024

