## Supplementary materials for "Using EEG to measure the neural effects of oxytocin administration: A meta-analysis and systematic review"

### Overview of included articles

The following tables summarise study characteristics for the studies included in the two meta-analyses. Each table contain the following columns:

1. Article: Reference to the specific study
2. Population: Type of population included in the study
3. B/W: Whether the study used a between-participant (B) or within-participant (W) design
4. Total N: Total number of participants in the study, excluding drop-outs
5. Mean age (SD): Mean age (and standard deviation) of participants in the study
6. Dose in IU: Dose(s) used in International Units (IU)
7. Time after adm.: Time elapsed between administration and testing
8. Task: Brief description of task(s) given to the participants
9. EEG analysis (main outcome): EEG measurement of interest for the main outcome(s)
10. Main outcome result: Brief description of the results of the study, focusing on the main outcome

#### Social/Cognitive MA articles

| Article | Population | B/W | Total N | Mean age (SD) | Dose in IU | Time after adm. | Task | EEG analysis (main outcome) | Main outcome result |
|---|---|---|---|---|---|---|---|---|---|
| (Alaerts, Taillieu, Daniels, et al., 2021) | Neurotypical males | B | 40 | 22.34 (2.82) | 24 | 30 min. | Participants watched social and non-social emotionally evocative stimuli and gave the stimuli subjective ratings of valence. | Frontal alpha μV2/Hz asymmetry | OT amplified approach related motivational salience of stimuli with stimuli considered to be of high personal relevance to the participant. |
| (de Bruijn et al., 2017) | Neurotypical males | W | 24 | 21.50 (1.90) | 24 | 40 min. | Groups of two in a social flanker task in two contexts: joint prize (social context) where individual errors affected the pairs' chance of winning, and individual prize (individual context) where errors only affected | ERP analysis of error-related negativity (ERN) amplitudes (μV) | OT affected performance monitoring of mistakes through enhanced ERN amplitudes only in the social context. |

| | | | | | | | | | |
|---|---|---|---|---|---|---|---|---|---|
| | | | | | | | own chances of winning. | | |
| (Festante et al., 2020) | Neurotypical males | B | 35 | 20.50 (1.80) | 24 | 45 min. | Visuo-motor task where participants grasped an object and either gave it to a person (social condition) or placed it in a container (non-social condition). | ERD/ERS analysis of alpha and beta frequencies, specifically the mu rhythm, measured in db (log10 EEG power/EEG baseline) | In sensorimotor areas during the social task, OT enhanced alpha desynchronization, and did not enhance beta synchronization. There were no differences between OT and PL in the non-social condition. |
| (Moerkerke et al., 2023) | Autistic males and females | B | 68 | 10.08 (1.33) | 12 (twice daily) | 24 hours; 4 weeks | Participants viewed neutral and expressive faces at baseline, post OT administration and 4 weeks after cessation of the daily OT administrations. | Frequency tagging analysis in left (LOT) and right occipito-temporal (ROT) regions, measured in µV | OT decreased neural sensitivity towards expressive faces. |
| (Moses et al., 2024) | Neurotypical males and females | W | 37 | 23.27 (3.87) | 24 | 50 min. | Participants were presented with images of faces with different facial expressions (neutral, fearful, happy). | ERP analysis of the P100, P1, N1, N170, VPP, N2 and P2 components, measured in ms | OT modulated early (P1, N1) stages of face processing regardless of emotion, and this modulation was sustained throughout (attenuation of P2). |
| (Mu et al., 2016) | Neurotypical males | B | 60 | 22.50 (2.50) | 24 | 40 min. | Participants cooperated in pairs (social task) or individually with the computer (non-social task) to count and press a button synchronously. | Alpha band phase locking value (PLV) analysed via time frequency analysis | In the social task, OT significantly decreased time lags and enhanced alpha band PLV, reflecting interbrain neural oscillations. Enhanced alpha band PLV after OT was also seen in the non-social condition. |
| (Ochiai et al., 2021) | Neurotypical males and females | B | 40 | 27.35 (5.79) | 24 | 5-10 min. | Auditory stimuli were presented simultaneously as a movie was | ERP analysis of N1 and Mismatch Negativity (MMN) | OT significantly shortened MMN peak latency, but not amplitude, suggesting that OT promotes the |

| | | | | | | | | | |
|---|---|---|---|---|---|---|---|---|---|
| | | | | | | | screened silently. Participants were asked to disregard the auditory stimuli and focus on the movie. | components, measured in μV and ms | comparison-decision stage of auditory discrimination. |
| (Paloyelis et al., 2016) | Neurotypical males | W | 13 | 25.69 (4.85) | 24 | 45-50 min. | Participants received nociceptive stimuli before and after treatment and provided ratings of perceived pain at specific time points during the trial. | Laser-evoked potentials (LEP) analysis of N1, N2 and P2 components, measured in μV | OT reduced the peak amplitudes of N1 and N2 and increased the latency of N2, suggesting that OT affects neural components that contribute to pain perception. There was no significant effect on P2. |
| (Peltola et al., 2018) | Neurotypical females (mothers) | W | 38 | 31.92 (4.98) | 24 | 55 min. | Participants were presented, in random order, pictures of infant and adult faces on a screen. | ERP analysis of N170 amplitudes, measured in μV | OT increased the face-sensitive N170 component amplitudes when viewing both adult and infant faces. |
| (Petereit et al., 2019) | Neurotypical females | B | 58 | 23.19 (2.60) | 24 | 31 min. | Participants participated in a social exclusion/inclusion cyberball game. | ERP analysis of LPP, measured as μV*ms | OT did not significantly change neural reactions to neither social nor non-social exclusion. However, during the exclusion phases in the control group, high LPP amplitudes and strong feelings of rejection was found, but this was not found in the OT group, indicating that OT dimished this neuro-affective relationship between LPP amplitudes and feelings of rejection. |
| (Qiao et al., 2022) | Neurotypical males | W | 31 | 22.81 (2.38) | 24 | 30 min. | Participants fixated on a cross in the centre of the screen while | Frequency tagging analysis of LOT, medial occipital | OT did not modulate any of the investigated neural responses when exposed to |

| | | | | | | | | | |
|---|---|---|---|---|---|---|---|---|---|
| | | | | | | | presented images of faces and houses superimposed on top of each other. | (MO) and ROT regions, measured in Hz | the different stimuli. |
| (Rutherford et al., 2017) | Neurotypical females | W | 24 | 23.00 (3.00) | 24 | 45 min. | Participants were presented images of houses (control), 24 adults and infants on a screen. 50% of the human stimuli had a neutral expression, whereas 50% were distressed. | ERP analysis of LPP, N170 and P300 components, measured in ms | When administered OT, the P300 ERP was larger in the infant condition than in the adult condition, suggesting that OT enhances the attention towards infants. N170 and LPP were unaffected by OT administration. |
| (Santiago et al., 2024) | Neurotypical males | B | 54 | 23.60 (3.72) | 24 | 29 min. | Participants completed a Social Salience Attribution Task (sSAT), where they were presented 20 images of fearful faces (social) and 20 images of fruit (non-social). | ERP analysis of N170, P2b, P3b and LPP amplitudes (μV) | OT significantly increased N170 amplitude, but not for the amplitudes of P2b, P3b and LPP, indicating that OT affects the early stages of salience attribution processing. |
| (Schiller et al., 2020) | Neurotypical males | B | 86 | 23.70 (4.59) | 24 | 45 min. | Participants took part in a decision-making task where their own resources could be sacrificed in order to alter the game-of chance outcomes of the other in-group and out-group participants. | ERP analysis of spatio-temporal microstates A, B, C and D. | Negative social behaviour towards out-group members was eliminated in the OT condition, where longer occurrences of microstate C and microstate D were suggested as a neurophysiological explanation. |
| (Schiller et al., 2023) | Neurotypical males and females | W | 144 | 23.49 (3.59) | 24 | 40 to 50 min. | Participants took part in a trust game with monetary endowment. Participants were | ERP analysis of peak intensity (global field power, GFP) of | In response to the trustees' face, P100 was intensified in male participants after OT administration, |

| | | | | | | | | | |
|---|---|---|---|---|---|---|---|---|---|
| | | | | | | | presented a picture person and asked to decide whether they trusted the person or not. | microstates and P100 | whereas the opposite occurred with the female participants, highlighting sex-specific effects of OT. |
| (Singh et al., 2016) | Neurotypical and schizophrenic males and females | W | 32 | 37 (15); 47 (16) | 24; 48 | 45 min. | Participants viewed videos of biological (social) and non-biological (non-social) motion. | Mu power suppression in left and right sensorimotor cortex for mirror neuron system (MNS) function | OT enhanced mu suppression index (MSSI) in all groups, but dose response different across the different conditions and genders. OT reached statistical significance only for the group of schizophrenic males who received 48 IU OT, suggesting OT may remediate MNS dysregulation associated with schizophrenia, especially for males. |
| (Soriano et al., 2020) | Neurotypical males | B | 40 | 22.33 (3.17) | 24 | 30 to 45 min. | Participants were instructed to observe and pay attention to live visual stimuli. The face of a female model was shown, where the model either gazed at the participant or closed her eyes. | Power spectral density of frontal alpha asymmetry (μV/Hz) | OT modulated left-sided frontal alpha asymmetry when the participant had direct eye contact with the live model, indicating increased approach-related motivational tendencies. The treatment effect was more prominent in participants with lower self-reported social motivation. |
| (Tillman et al., 2019) | Neurotypical males | W | 21 | 25.20 (3.68) | 24 | 45 min. | Participants were presented with 70 computer-generated, grayscale faces (50% female). | ERP analysis of N170, P100 and Earlier Posterior Negativity (EPN), measured | OT reduced N170 latency to fearful faces and EPN amplitudes to fearful faces were less lateralised. There were no significant OT |

| | | | | | | | | | |
|---|---|---|---|---|---|---|---|---|---|
| | | | | | | | Each face was presented twice during the trial, once with a neutral expression and once with a fearful expression. | in µV and ms | effects on N170 amplitude or P100 latency and amplitude. |
| (Van der Donck et al., 2022) | Neurotypical males | W | 31 | 22.81 (2.38) | 24 | 30 min. | Participants were presented with faces at 6 Hz on a screen. The faces had a neutral expression, except every fifth face which were expressive (angry, happy, fearful). | Frequency tagging analysis of LOT, ROT and MO regions, measured in µV | OT did not influence neural sensitivity to facial expressions. The authors did not find that OT enhanced emotional salience or modulated social approach-avoidance tendencies. |
| (Zhang et al., 2021) | Neurotypical males and females | B | 61 | 20.27 (1.90) | 24 | 45 min. | Participants were put in a pseudo “speed date” setting where they were presented with images of the opposite sex and asked if they were interested in getting to know that person. The participants were then informed about their “speed date’s” decision. | Time frequency analysis of theta oscillations and power (µV) | Increased frontal midline theta oscillations were found during romantic rejection, and OT administration significantly decreased these oscillations. OT also decreased the connection between theta power and rejection based negative emotions. The results suggest OT has a social pain reducing effect. |
| (Zhuang et al., 2021) | Neurotypical males | W | 33 | 21.15 (2.18) | 24 | 45 min. | The study utilised a probabilistic reversal learning task with two sets of six hiragana syllables. The task consisted of a a learning phase and a | ERP analysis of feedback-related negativity (FRN), measured in µV | OT decreased FRN difference for negative minus positive feedback in the learning phase, suggesting that OT facilitates learning by rendering negative and positive feedback |

| | | | | | | | | | |
|---|---|---|---|---|---|---|---|---|---|
| | | | | | | | testing phase. | | evaluations more equivalent, rather than different. |

#### Neural MA articles

| Article | Population | B/W | Total N | Mean age (SD) | Dose in IU | Time after adm. | Task | EEG analysis (main outcome) | Main outcome result |
|---|---|---|---|---|---|---|---|---|---|
| (Alaerts, Taillieu, Prinsen, et al., 2021) | Neurotypical males | B | 96 | 22.42 (3.06) | 24 | 30 min. | Resting-state with eyes open. | Decoupling/coupling. Cross-frequency dynamics between alpha and theta bands to investigate harmonic and non-harmonic cross-frequency relationships | OT increased decoupling of theta and alpha rhythms, indicating OT plays a role in increasing the intrinsic EEG frequency structure's signal-to-noise ratio. |
| (Alaerts et al., 2024) | Autistic males and females | B | 67 | 10.49 (1.28) | 12 (twice daily) | 4 weeks | Resting-state with eyes open. | Frequency tagging analysis of harmonic and non-harmonic relationships between the alpha and theta bands | OT increased the formation of non-harmonic cross-frequency architecture, suggesting a reduction of "neural noise" and a gradual shift in the intrinsic EEG cross-frequency to the pattern observed in non-autistic children. |
| (Perry et al., 2010) | Neurotypical males | W | 23 | 25.30 (3.55) | 24 | 45 min. | The participants were presented video clips of point-light displays representing biological motion based on human walkers. The participants were asked to count the | Suppression index based on EEG power ratio logarithm for alpha and beta frequency bands, specifically the mu rhythm | OT significantly suppressed the mu rhythm in the 8-10 Hz and 15-25 Hz range. |

| | | | | | | | | | |
|---|---|---|---|---|---|---|---|---|---|
| | | | | | | | occurrences of a rare event and report this number at the end of each trial. | | |
| (Rutherford et al., 2018) | Neurotypical females | W | 23 | 23.30 (3.30) | 24 | 70 min. | Resting-state with eyes open and eyes closed. | Spectral data EEG power analysis of delta-beta coupling | OT decreased cross-frequency delta-beta coupling, especially in the eyes closed condition. |
| (Schiller et al., 2019) | Neurotypical males | B | 86 | 23.70 (4.59) | 24 | 45 min. | Resting-state with eyes open and eyes closed. | Analysis of microstates A, B, C and D. | OT significantly increased duration of all microstates and significantly reduced the occurrences of autonomic processing-related networks to favour attention-related networks. |
| (Tomescu et al., 2024) | Neurotypical males | W | 30 | 22.80 (2.40) | 24 | 45 min. | Resting-state with eyes open. | Analysis of microstates A, B, C, D, E, F and G. | On the group level, OT significantly increased the mean duration and occurrence of attention and salience microstates, while it simultaneously decreased default mode network associated microstates. |
| (Zelenina et al., 2022) | Neurotypical males | W | 19 | 27.10 (4.02) | 24 | 15 to 90 min. | Resting-state with eyes open and eyes closed, 7 time windows (including baseline) | Analysis of microstates A, B, C and D. | OT affected all microstates, both with eyes open and eyes closed. The authors suggest OT may make the brain more receptive for external, and especially social, stimuli by recruiting attentional networks. |

### Publication bias assessments

#### Social/Cognitive MA

| Test | Least variance | Largest variance |
|---|---|---|
| PET | M = 0.015, CI [0.000, 0.243] | M = 0.241, CI [0.000, 1.744] |
| PEESE | M = 0.033, CI [0.000, 0.543] | M = 1.155, CI [0.000, 5.789] |
| mean $\mu$ | -0.010, *CI* [-0.190, 0.040] | 0.027, *CI* [-0.310, 0.372] |
| Inclusion bf for effect | 0.149 | 0.413 |
| Posterior probability | 0.130 | 0.292 |

PET tests if the effect size is influenced by study precision by regressing effect sizes on their standard errors, while PEESE adjusts for small-study effects by correcting for precision-related bias via squared standard errors. PET and PEESE estimates close to zero with wide confidence intervals indicate little to no bias, as is evident in the studies with the least variance, whereas larger estimates and narrower confidence intervals indicate a potential presence of bias, as seen with the studies with larger variances.

Adjusting for bias reduced the effect size for both variance groups, indicating that observed effects may be due to publication bias or small-study effects, and that the "true" effect may be negligible. While the mean effect size for the studies with the largest variance is slightly higher, the confidence intervals are much larger, indicating greater uncertainty.

Inclusion bayes factor for the effect and the posterior probability suggest weak evidence for a true effect across the studies and that a null-model is favoured.

#### Neural MA

| Test | Least variance | Largest variance |
|---|---|---|
| PET | M = 0.06, CI [0.00, 0.72] | M = 0.24, CI [0.00, 2.17] |
| PEESE | M = 0.15, CI [0.00, 2.36] | M = 1.155, CI [0.000, 5.789] |
| mean $\mu$ | 0.13, *CI [-0.05 to 0.21]* | 0.27, *CI [-0.10 to 0.75]* |
| Inclusion bf for effect | 0.20 | 1.81 |
| Posterior probability | 0.16 | 0.65 |

The possibility of publication or small study bias was supported by the model-averaged PET estimates, suggesting that small-study effects or biases may be influencing the overall results. Additionally, the PEESEs corrected for small-study effects by adjusting the overall effect size for the inflation seen in smaller studies.

The Robust Bayesian meta-analysis adjusted effect size was much lower than seen in the multilevel meta-analysis ($\mu = 0.31$), suggesting that publication bias or small-study effects may have inflated the effect size in the multilevel meta-analysis.

The inclusion Bayes factor for the effect and the posterior probability indicated some evidence for a true effect across studies with the largest variances, while the studies with the least variances showed support for a null model.

#### Heterogeneity measures

We calculated both Cochran's Q and $I^2$. Cochran's Q tests are commonly used for total residual heterogeneity, testing if the observed variability in effect sizes exceeds what would be expected from random sampling error. It is however sensitive to the number of studies included, as it may overestimate heterogeneity in large samples. It may also fail to detect heterogeneity if the sample is small (Borenstein et al., 2009). $I^2$ is derived from Cochran's Q, but instead of being an absolute measure, it quantifies the *proportion* of total variance that can be attributed to true heterogeneity as opposed to sampling error, thus providing a measure of inconsistency across studies. Since it is a relative measure, $I^2$ is less affected by sample size compared to Cochran's Q (Higgins & Thompson, 2002).

GOSH plots were also computed. GOSH plots can identify outliers, subgroups or other patterns in the data that may not be detected by traditional tests of heterogeneity and thus indicates variability that is unaccounted for.

### Forest plots

#### Social/Cognitive MA

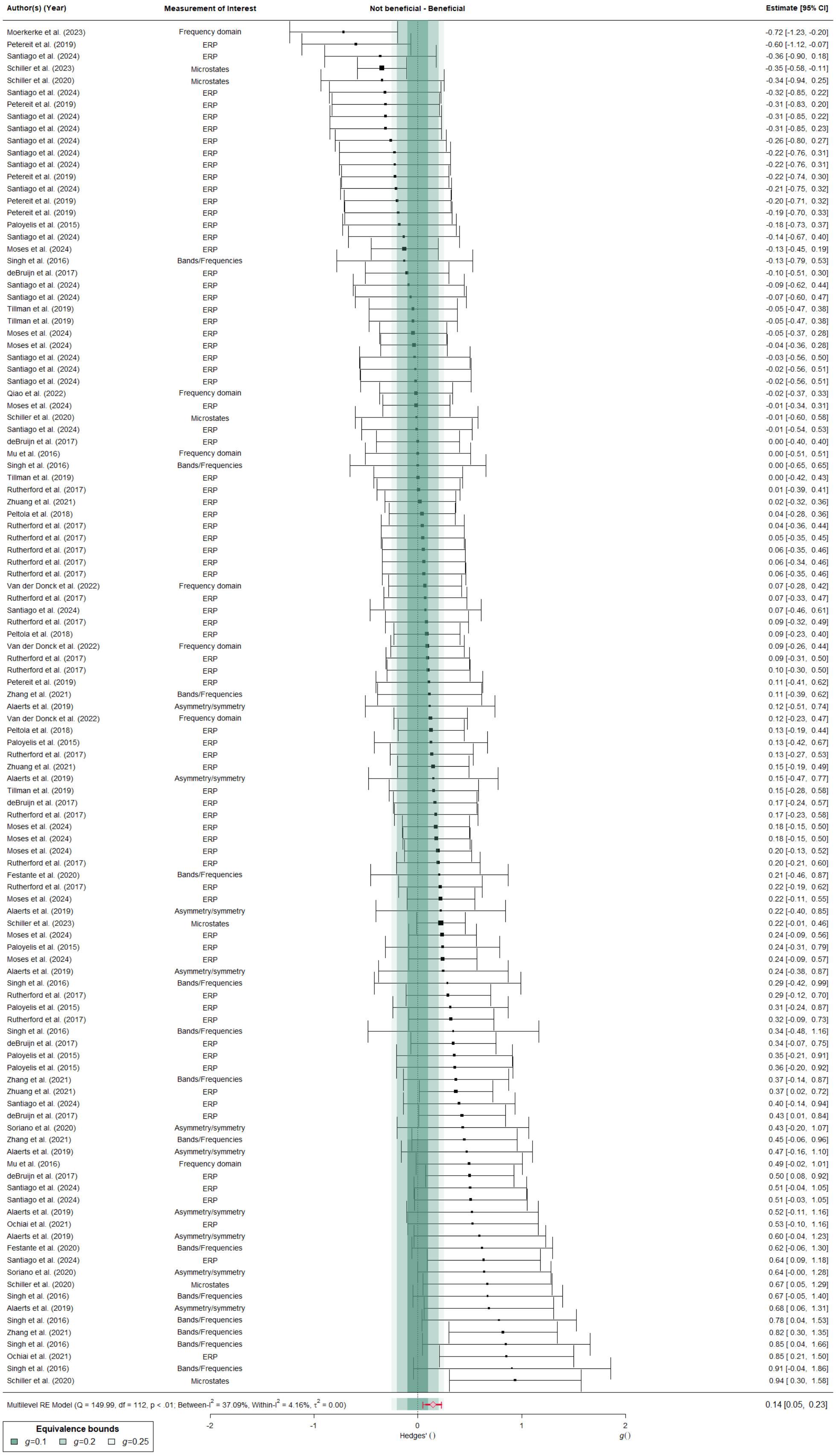

*Forest plot for the Cognitive MA, derived from a multilevel random-effects model applied to data from 21 studies, encompassing a total of 113 extracted effect sizes. Shades of green represent equivalence bounds (Hedges' g of 0.1, 0.2, and 0.25). Each individual effect size is shown as a filled square, with its size indicating the weight of the point estimate. Thin lines represent the 95% confidence intervals. The red diamond at the bottom shows the summary effect size along with its 95% CI.*

#### Neural MA

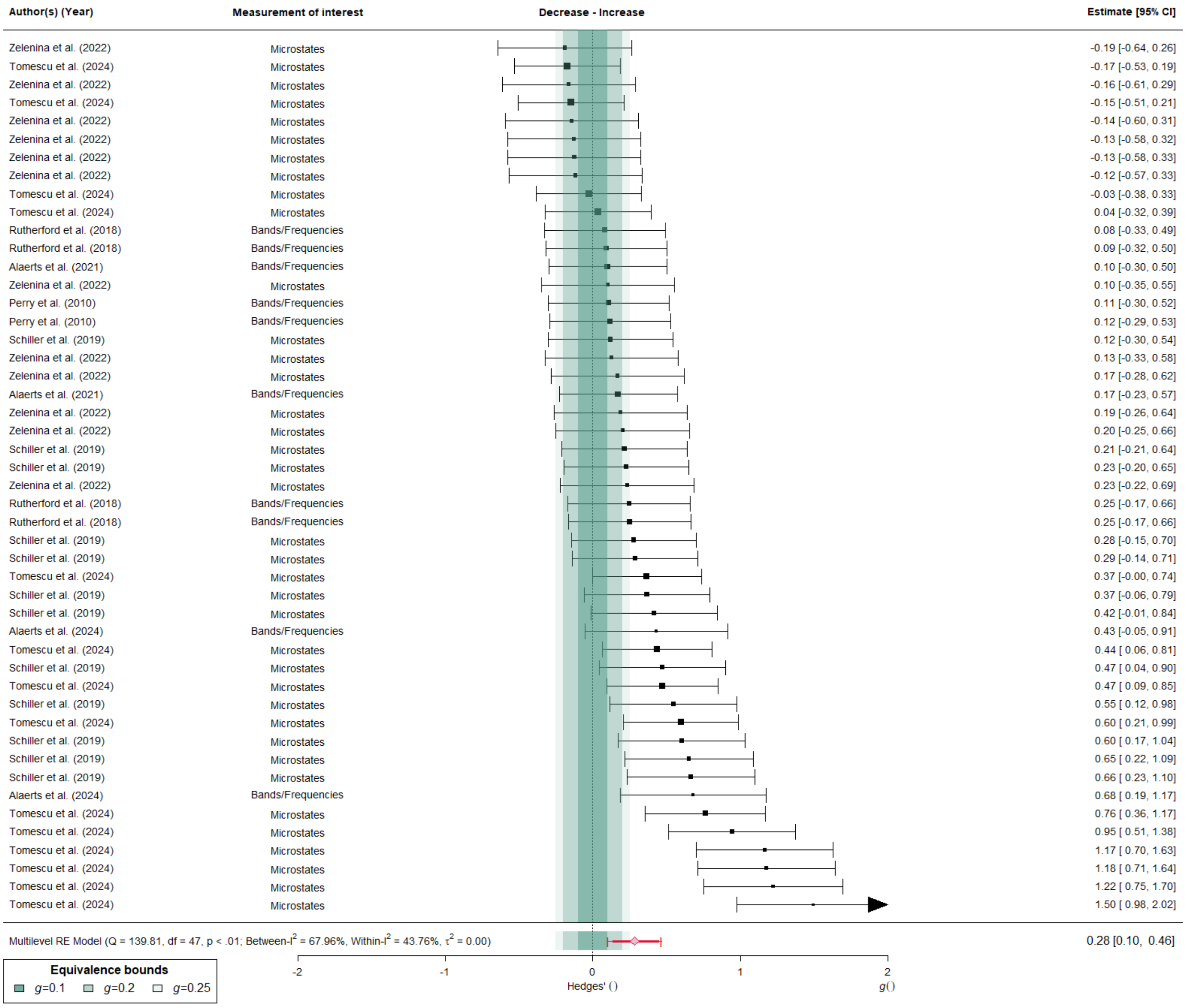

*Forest plot for the Neural MA based on the data from a multilevel random-effects model performed on 7 studies with a total of 48 extracted effect sizes. Shades of green represent equivalence bounds (Hedges' g of 0.1, 0.2, and 0.25). Individual effect sizes are shown as filled squares, with the size reflecting the weight of the point estimate. The 95% confidence intervals are indicated by thin lines. The far-right column displays the effect sizes and their associated 95% CIs for each individual extracted effect size. The red diamond at the bottom represents the summary effect size and its corresponding 95% CI.*

### Influence Diagnostics, Leave-One-Out Analyses and GOSH diagnostics

This section includes figures created when conducting influence diagnostics and Leave-One-Out (LOO) analysis, as well as GOSH diagnostics outputs.

#### Influence diagnostics plots

This section includes a table for each meta-analysis provide an overview of study IDs, alongside accompanying influence diagnostics plots.

Below are influence diagnostics plots for the meta-analysis subsets with the least and the largest variances. Each panel visualizes a different diagnostic statistic across studies. Dotted horizontal lines represent reference thresholds or medians. Red dots indicate studies flagged as particularly influential or potential outliers. The plots include the following: studentized residuals (rstudent), change in fitted values (dffits), Cook's distance (cook.d), covariance ratios (cov.r), changes in heterogeneity (tau2.del), change in model fit (QE.del), study leverage (hat), and study weight (weight).

##### Social/Cognitive MA

| ID | Study |
|---|---|
| 1 | (Alaerts, Taillieu, Daniels, et al., 2021) |
| 2 | (de Bruijn et al., 2017) |
| 3 | (Festante et al., 2020) |
| 4 | (Moerkerke et al., 2023) |
| 5 | (Mu et al., 2016) |
| 6 | (Ochiai et al., 2021) |
| 7 | (Paloyelis et al., 2016) |
| 8 | (Peltola et al., 2018) |
| 9 | (Petereit et al., 2019) |
| 10 | (Qiao et al., 2022) |
| 11 | (Santiago et al., 2024) |
| 12 | (Schiller et al., 2023) |
| 13 | (Soriano et al., 2020) |
| 14 | (Tillman et al., 2019) |
| 15 | (Van der Donck et al., 2022) |
| 16 | (Zhuang et al., 2021) |
| 17 | (Zhang et al., 2021) |
| 18 | (Schiller et al., 2020) |
| 19 | (Moses et al., 2024) |
| 20 | (Rutherford et al., 2017) |
| 21 | (Singh et al., 2016) |

#### Data with the least variance

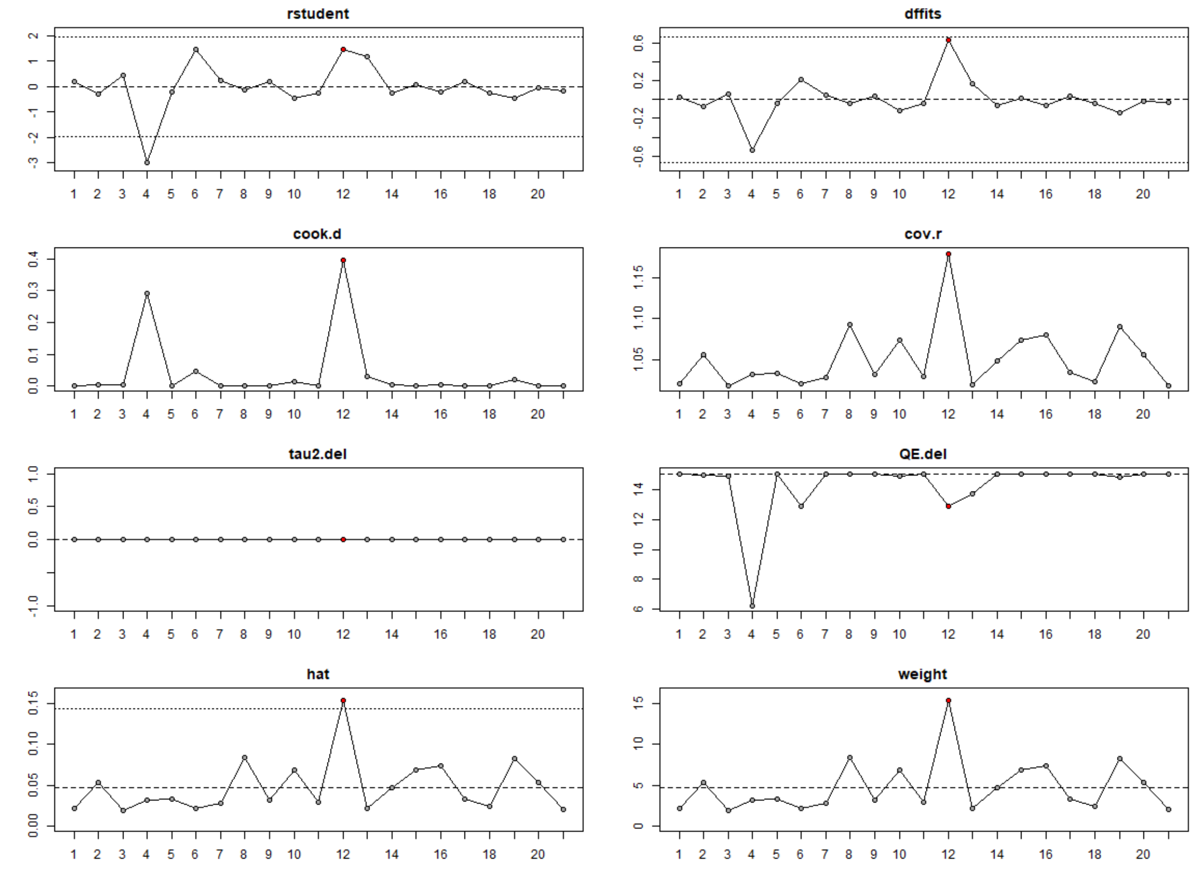

#### Data with the largest variance

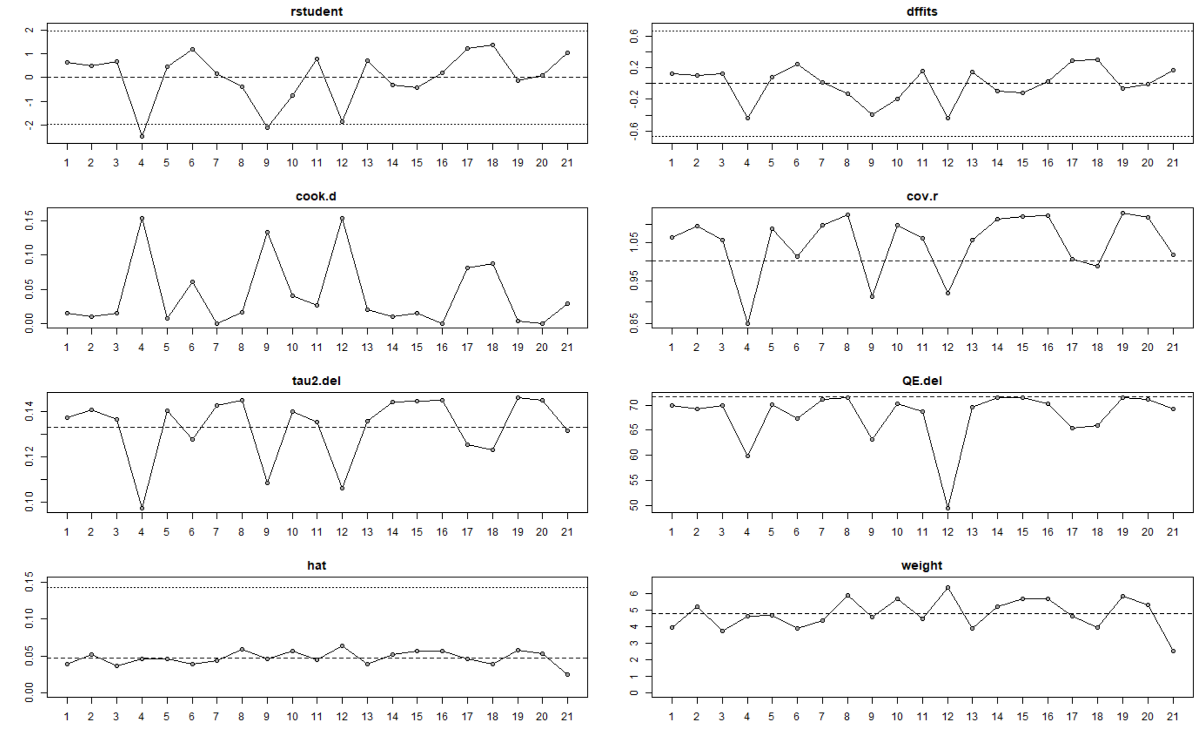

#### Neural MA

| ID | Study |
|---|---|
| 1 | (Alaerts, Taillieu, Prinsen, et al., 2021) |
| 2 | (Alaerts et al., 2024) |
| 3 | (Perry et al., 2010) |
| 4 | (Schiller et al., 2019) |
| 5 | (Zelenina et al., 2022) |
| 6 | (Tomescu et al., 2024) |
| 7 | (Rutherford et al., 2018) |

**Data with the least variance**

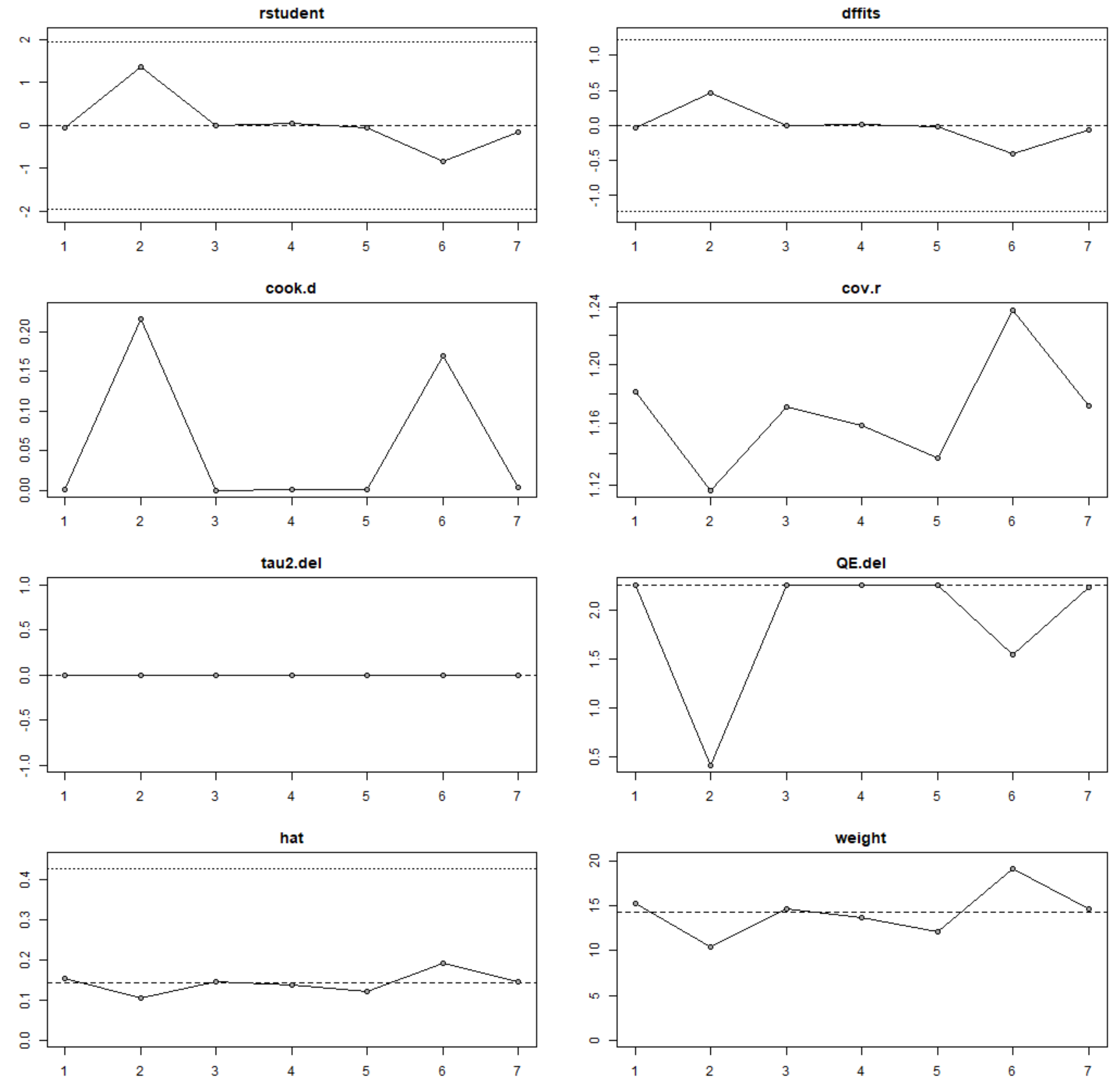

**Data with the largest variance**

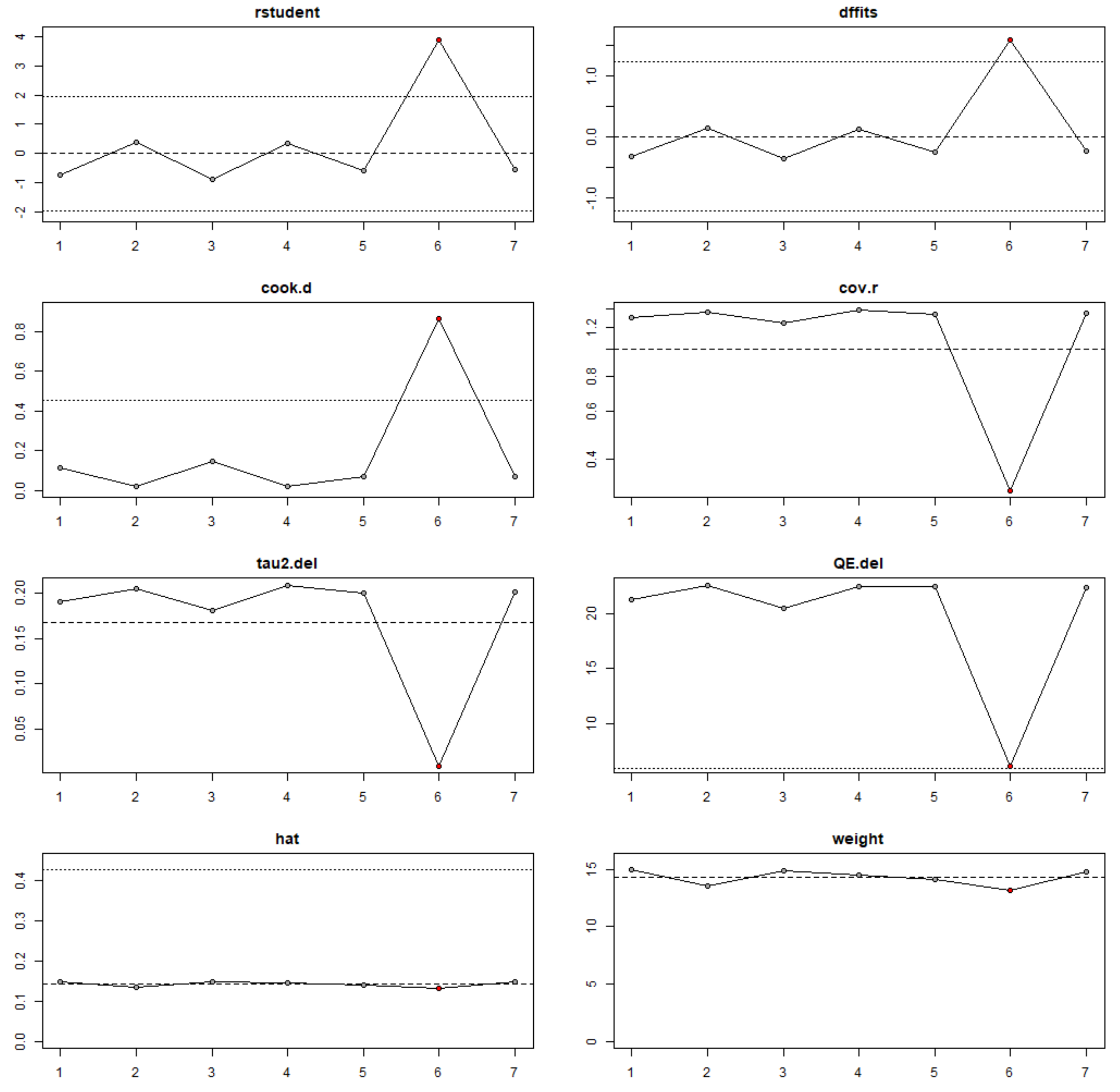

#### Leave-One-Out analysis plots:

Two plots are generated for each meta-analysis. Note that the plots here display individual effect sizes, not individual studies.

Left plot: Visualises change in the estimated intercept coefficient when each effect size is excluded. Each point represents the model's intercept estimate excluding one effect size. The dashed red line indicates the intercept from the full model including all effect sizes.

Right plot: Visualises change in the residual heterogeneity statistic (QE) after excluding each effect size. Each point represents the QE value from a model excluding one effect size. The dashed red line indicates the QE from the full model including all effect sizes.

#### Social/Cognitive MA:

##### Leave-One-Out analysis

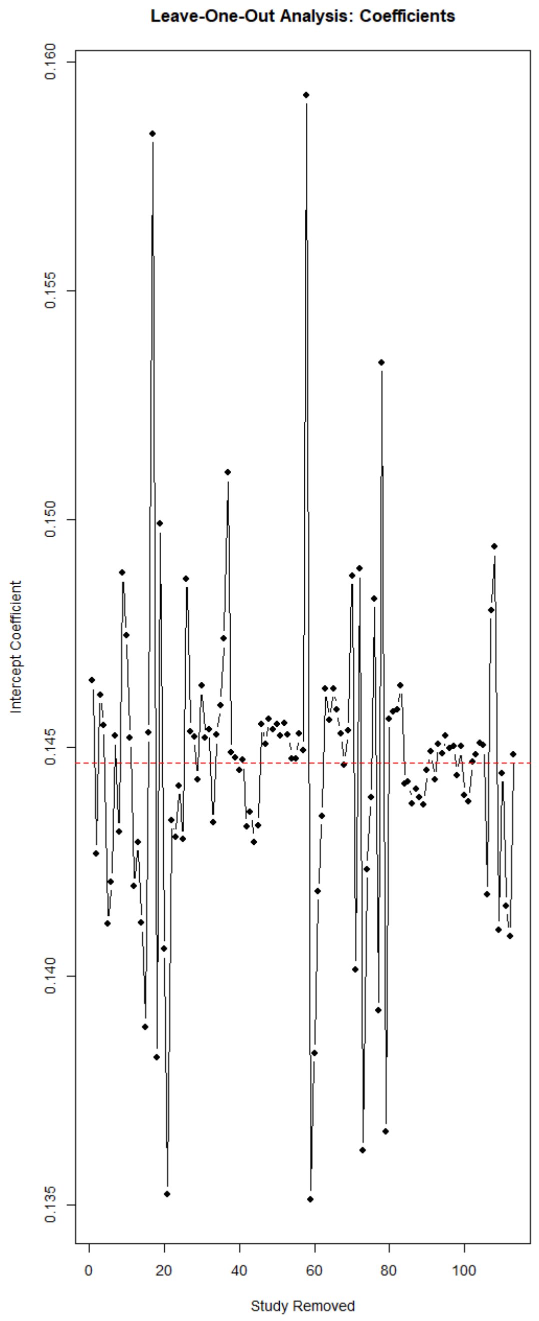

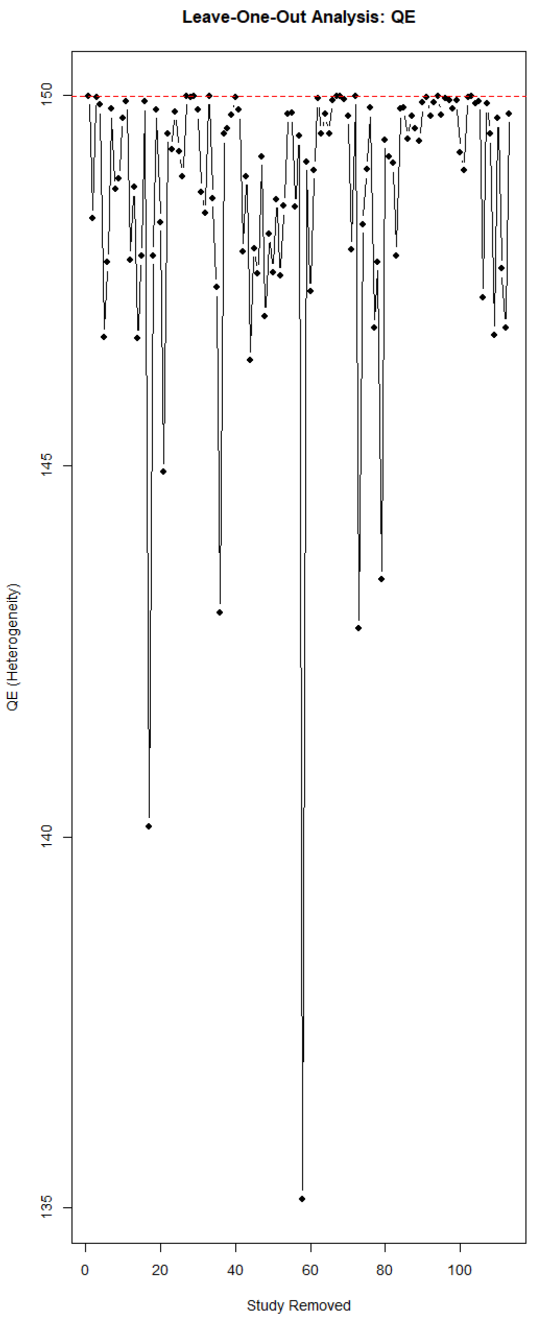

#### Neural MA

**Leave-One-Out analysis**

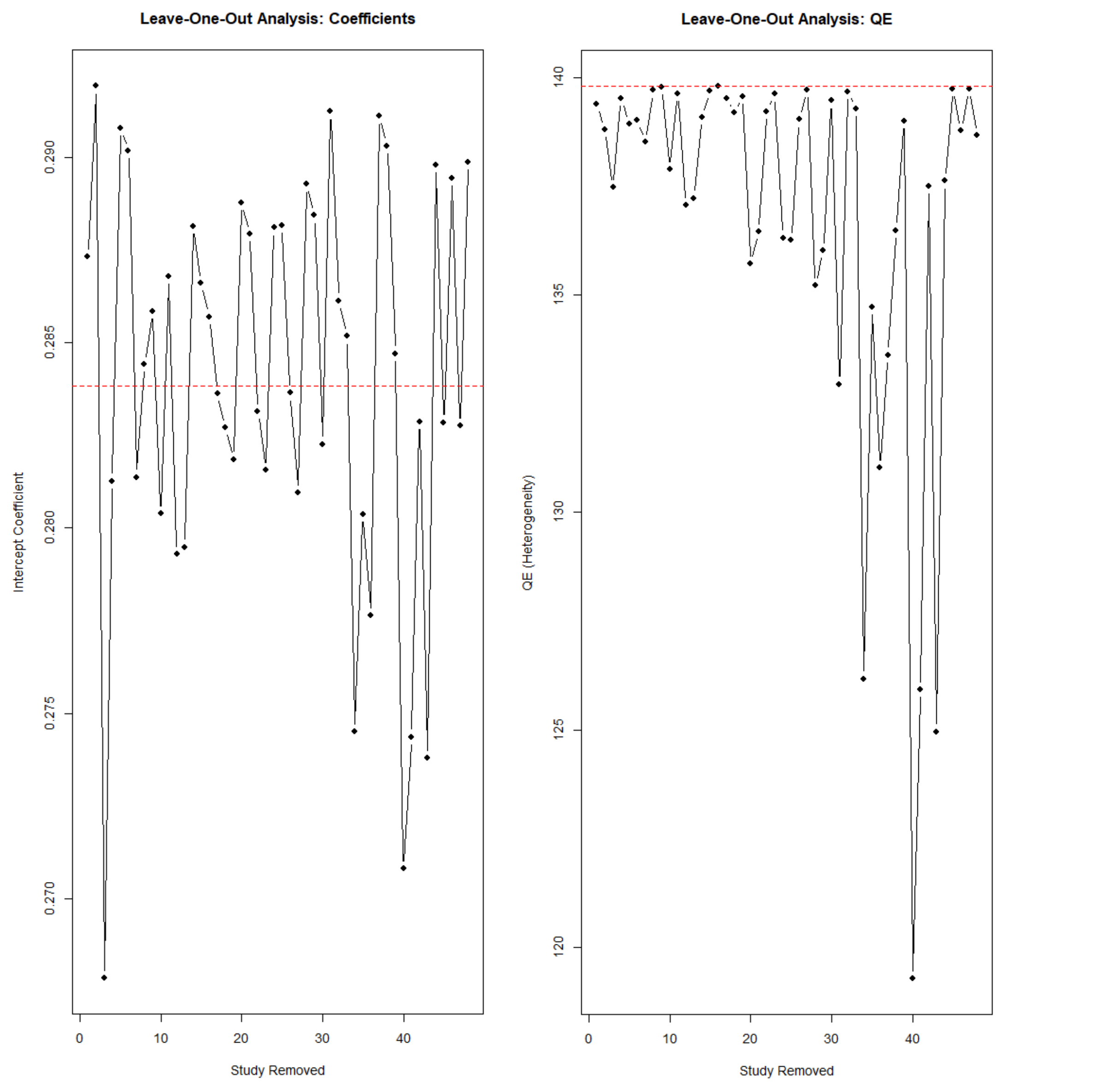

#### GOSH diagnostics

This section lists all effect sizes listed as potential outliers in GOSH diagnostics by the clustering algorithms k-means, density reachability and connectivity clustering (DBSCAN) and Gaussian Mixture Models (GMM).

K-means partition the GOSH plot into *k* clusters where each point belongs to the cluster with the nearest mean, detecting clusters of subsets yielding similar effect sizes and heterogeneity values. K-means clusters (centers) were set to 2 for both meta-analyses.

DBSCAN finds clusters based on density, detecting outliers as noise. The parametres epsilon (ε) and MinPts refer to the maximum distance between two points to be considered neighbours and the minimum number of points to form a cluster core, respectively. ε was set to 0.08 for both meta-analyses. MinPts parametres were set to 100 and 40 for the Social/Cognitive MA and the Neural MA, respectively.

GMM assumes the data is generated from several Gaussian distributions, where each cluster is modelled as a multivariate normal distribution. GMM can detect distinct underlying clusters of models with similar effect sizes and $\tau^2$ estimates.

#### Social/Cognitive MA

| | Output | Parametres (where applicable) |
|---|---|---|
| **K-means** | 58 | Centers = 2 |
| **DBSCAN** | 17, 58 | ε = 0.08; MinPts = 100 |
| **GMM** | 58, 73, 79, 17, 14, 21, 44, 36 | NA |
| **ID (study)** | 14 (de Bruijn et al., 2017)<br>17 (Moerkerke et al., 2023)<br>21 (Ochiai et al., 2021)<br>36 (Petereit et al., 2019)<br>44 (Santiago et al., 2024)<br>58 (Schiller et al., 2023)<br>73 (Zhang et al., 2021)<br>79 (Schiller et al., 2020) | |

#### Neural MA

| | Output | Parametres (where applicable) |
|---|---|---|
| **K-means** | 34, 40, 41, 43 | Centers = 2 |
| **DBSCAN** | 34, 40, 41, 43, 17, 28, 42, 19, 31 | ε = 0.08; MinPts = 40 |
| **GMM** | 28, 31, 37, 40, 34, 41, 43 | NA |
| **ID (study)** | 17 (Schiller et al., 2019)<br>19, 28 (Zelenina et al., 2022)<br>31, 34, 40, 41, 42, 43 (Tomescu et al., 2024) | |

### Equivalence testing

The plots display confidence intervals from equivalence tests (TOST) for individual studies (not individual effects) at three SESOI thresholds: ±0.1, ±0.2 and ±0.25.

The lower (LL) and upper (UL) bounds of the 90% CIs used in the TOST procedure for each study are grouped by SESOI threshold and whether the point represents the lower or upper bound of the CI.

The horizontal dashed red lines denote the SESOI threshold values (0.1, 0.2, and 0.25, respectively).

### Social/Cognitive MA

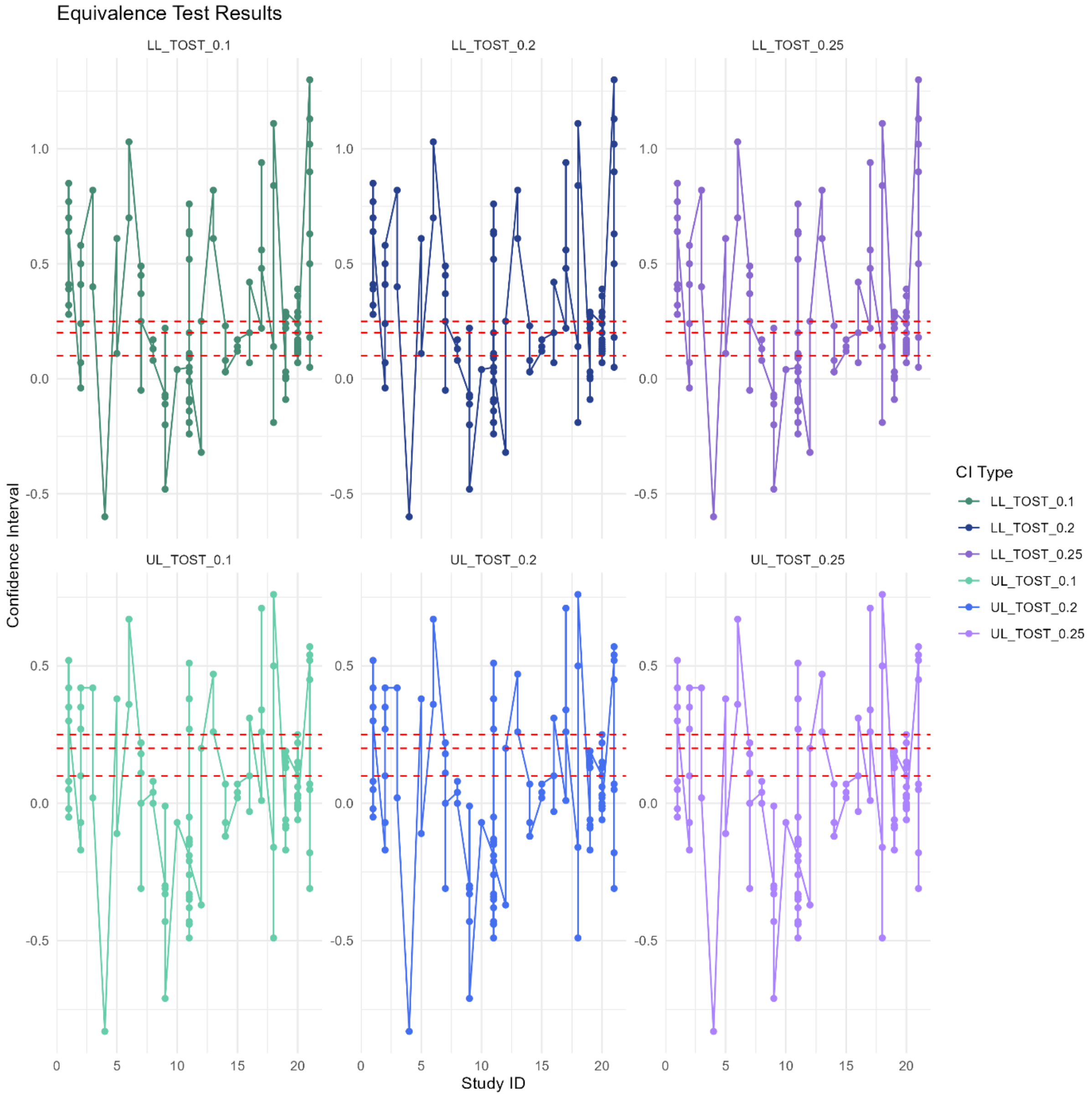

### Neural MA

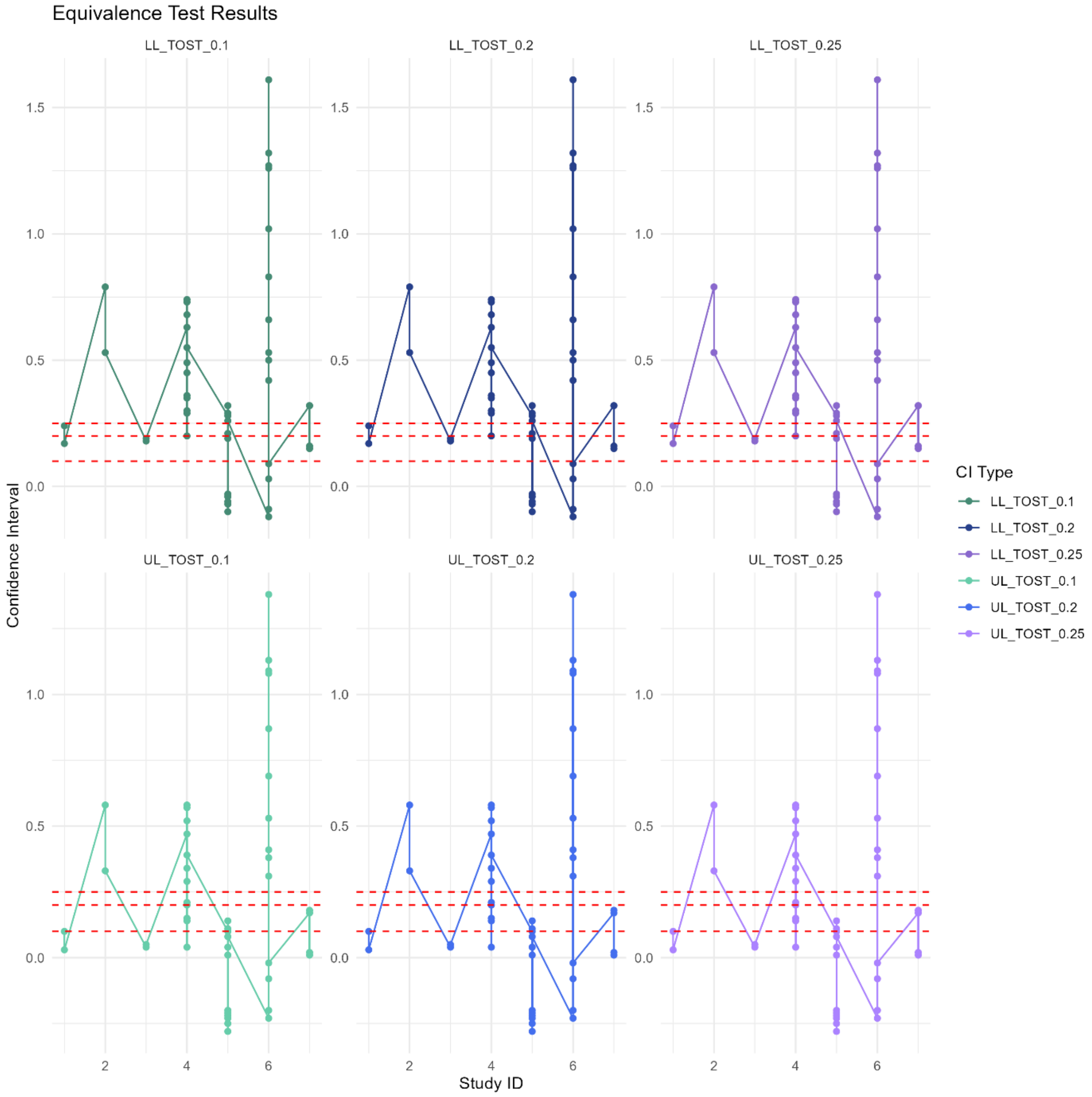

Equivalence Test Results
LL_TOST_0.1
LL_TOST_0.2
LL_TOST_0.25
UL_TOST_0.1
UL_TOST_0.2
UL_TOST_0.25
Confidence Interval
Study ID
CI Type
LL_TOST_0.1
LL_TOST_0.2
LL_TOST_0.25
UL_TOST_0.1
UL_TOST_0.2
UL_TOST_0.25

### Moderator analyses

#### Social/Cognitive MA

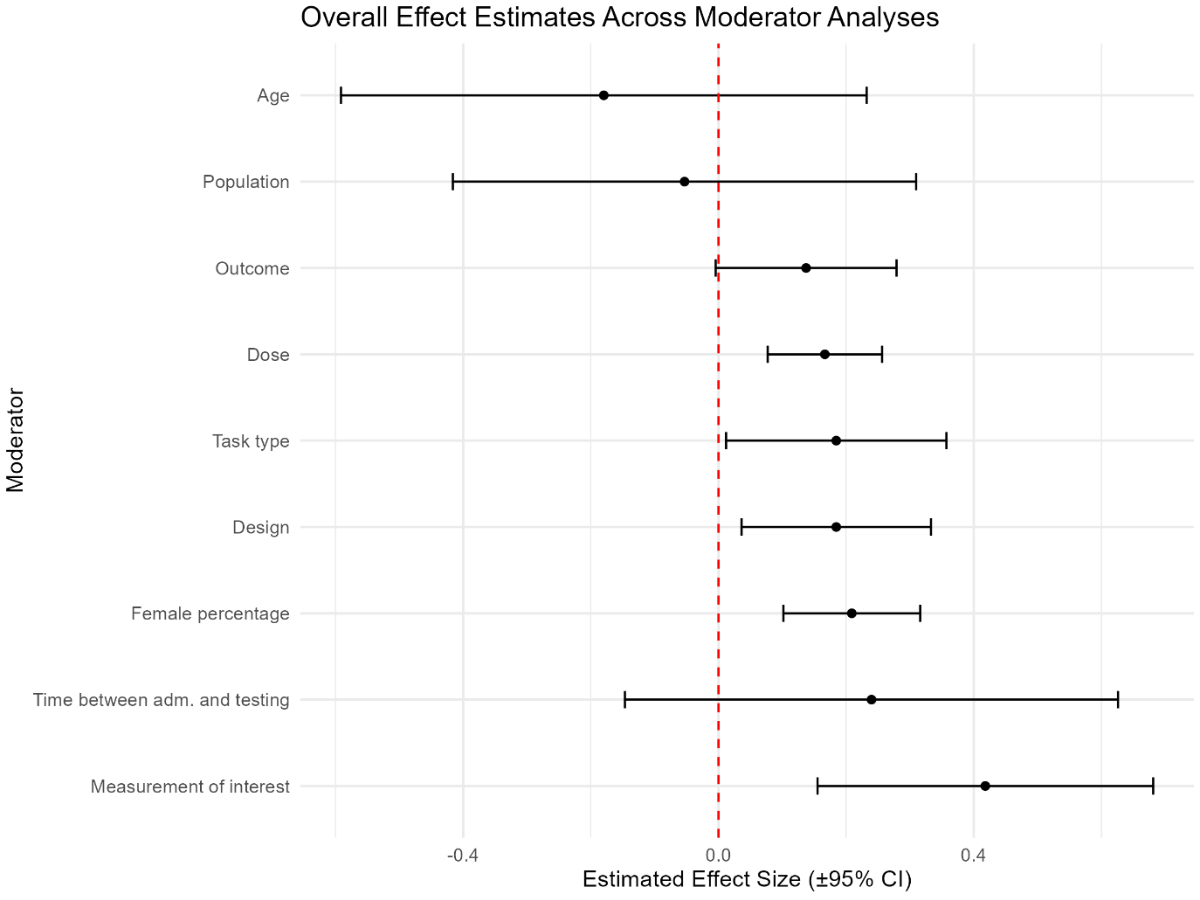

#### Non-signficant moderators

For significant moderators, please refer to the main text.

| Moderator | Effect (Hedge's *g)* | Residual heterogeneity | Test of moderator |
|---|---|---|---|
| **Outcome** | | QE(df = 111) = 149.5368, p = 0.0087 | QM(df = 1) = 0.0210, p = 0.8848 |
| Non-social<br>Social | 0.137<br>-0.1266 | | |
| **Task type** | | QE(df = 110) = 146.2674, p = 0.0118 | QM(df = 2) = 0.2706, p = 0.8735 |
| Behavioural<br>Visual and behavioural<br>Visual | 0.1846<br>-0.2361<br>0.2417 | | |
| **Time in minutes between administration and testing (only single dose studies)** | | QE(df = 110) = 139.8398, p = 0.0288 | QM(df = 1) = 0.1698, p = 0.6803 |
| Per 1 minute increase | -0.0020 | | |

| | | | |
|---|---|---|---|
| **Population** | | QE(df = 111) = 149.6775, p = 0.0085 | QM(df = 1) = 1.2245, p = 0.2685 |
| Neurotypical<br>Clinical | -0.0531<br>0.2609 | | |
| **Age** | | QE(df = 111) = 146.8575, p = 0.0128 | QM(df = 1) = 2.4952, p = 0.1142 |
| Per 1 year increase in mean age | 0.0134 | | |

*Overview of non-significant moderator analyses for social and non-social outcomes. Listed effects for all variables are real effects, i.e. the sum after subtracting the intercept value from the moderator. For each moderator analysis in the table, the first listed effect is the intercept. Significant residual heterogeneity is not highlighted.*

#### Neural MA

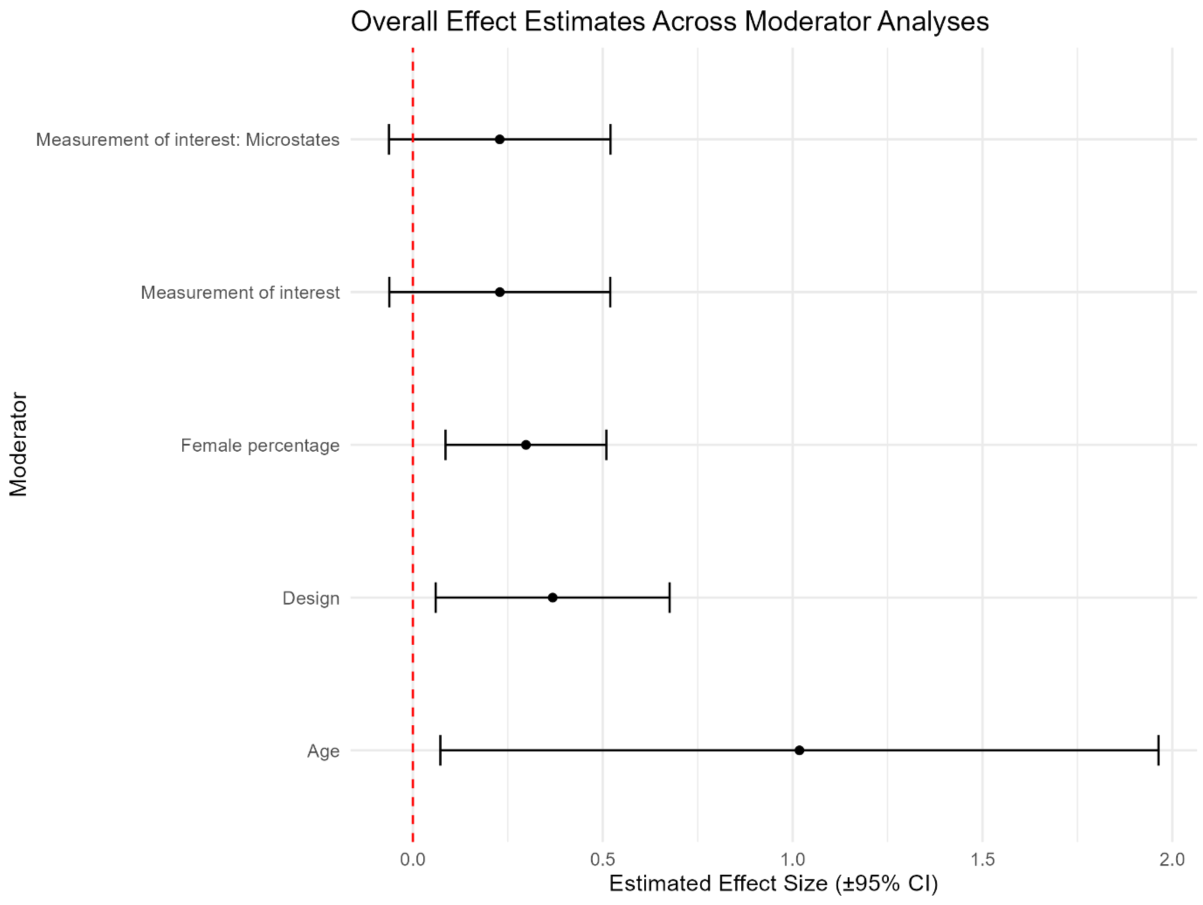

##### Non-signficant moderators

In addition to the pre-specified moderator analyses, a moderator analysis on outcomes where microstate outcomes were divided in groups (duration, occurrence, coverage, contribution) was also conducted based on literature pointing to the heterogeneity of these outcomes (Michel & Koenig, 2018), despite two of these groups (contribution, coverage) only including four effect sizes. Neither the test of moderator nor any of the moderators reached significance, and there was still significant residual heterogeneity.

| Moderator | Effect (Hedges' *g)* | Residual heterogeneity | Test of moderator |
|---|---|---|---|
| **Measurement of interest** | | QE(df = 46) = 137.3322, p < .0001 | QM(df = 1) = 0.2323, p = 0.6298 |
| Frequency bands<br>Microstates | 0.2289<br>0.3237 | | |
| **Measurement of interest: Separate microstate groups** | | QE(df = 43) = 128.6513, p < .0001 | QM(df = 4) = 0.9730, p = 0.9139 |
| Frequency bands<br>Microstates: Contribution<br>Microstates: Coverage<br>Microstates: Duration<br>Microstates: Occurrence | 0.2286<br>-0.2052<br>-0.1713<br>-0.1599<br>-0.0725 | | |
| **Age** | | QE(df = 46) = 118.3252, p < .0001 | QM(df = 1) = 1.7695, p = 0.1834 |
| Per 1 year increase in mean age | -0.0297 | | |

*Overview of non-significant moderator analyses for neural outcomes. Listed effects for all variables are real effects, i.e. the sum after subtracting the intercept value from the moderator. For each moderator analysis in the table, the first listed effect is the intercept. Significant residual heterogeneity is not highlighted.*

### Outlier removals

#### Social/Cognitive MA

| | Full model | Model with outcome 58 removed |
|---|---|---|
| **Model fit** | | |
| AIC<br>BIC | 32.4280<br>40.5835 | 22.3496<br>30.4782 |
| **Heterogeneity** | | |
| QE | 149.9877 | 135.1191 |
| *p*-value | 0.0096 | 0.0596 |
| **Estimate** | | |
| Hedges' *g* | 0.1447 | 0.1593 |
| *p*-value | 0.0016 | 0.0003 |

| **Robust Bayesian Meta-Analysis (RoBMA)** | | | | |
|---|---|---|---|---|
| | **Full model** | | **Outcome 34, 40, 41 and 43 removed** | |
| | **Least** | **Largest** | **Least** | **Largest** |
| **Inclusion BF** | | | | |
| Effect | 0.15 | 0.41 | 0.15 | 0.53 |
| Heterogeneity | 0.29 | 413.44 | 0.29 | 93.23 |
| Bias | 1.21 | 9.57 | 1.21 | 6.07 |
| **Mean estimates** | | | | |

| | | | | |
|---|---|---|---|---|
| PET | 0.02 | 0.24 | 0.02 | 0.25 |
| PEESE | 0.03 | 1.16 | 0.03 | 0.89 |
| *μ* | -0.01 | 0.03 | -0.01 | 0.07 |

*Comparison of results of the multilevel meta-analysis (top) and RoBMA (bottom) results for neural outcomes. The results from with all eligible studies included is shown at the left, whereas the results with outliers removed is presented to the right.*

With the one outlier removed, the Egger's regression test for funnel plot asymmetry was not significant (t(110) = -0.35, p = 0.7294), with a bias estimate of -1.8142 (SE = 5.2305).

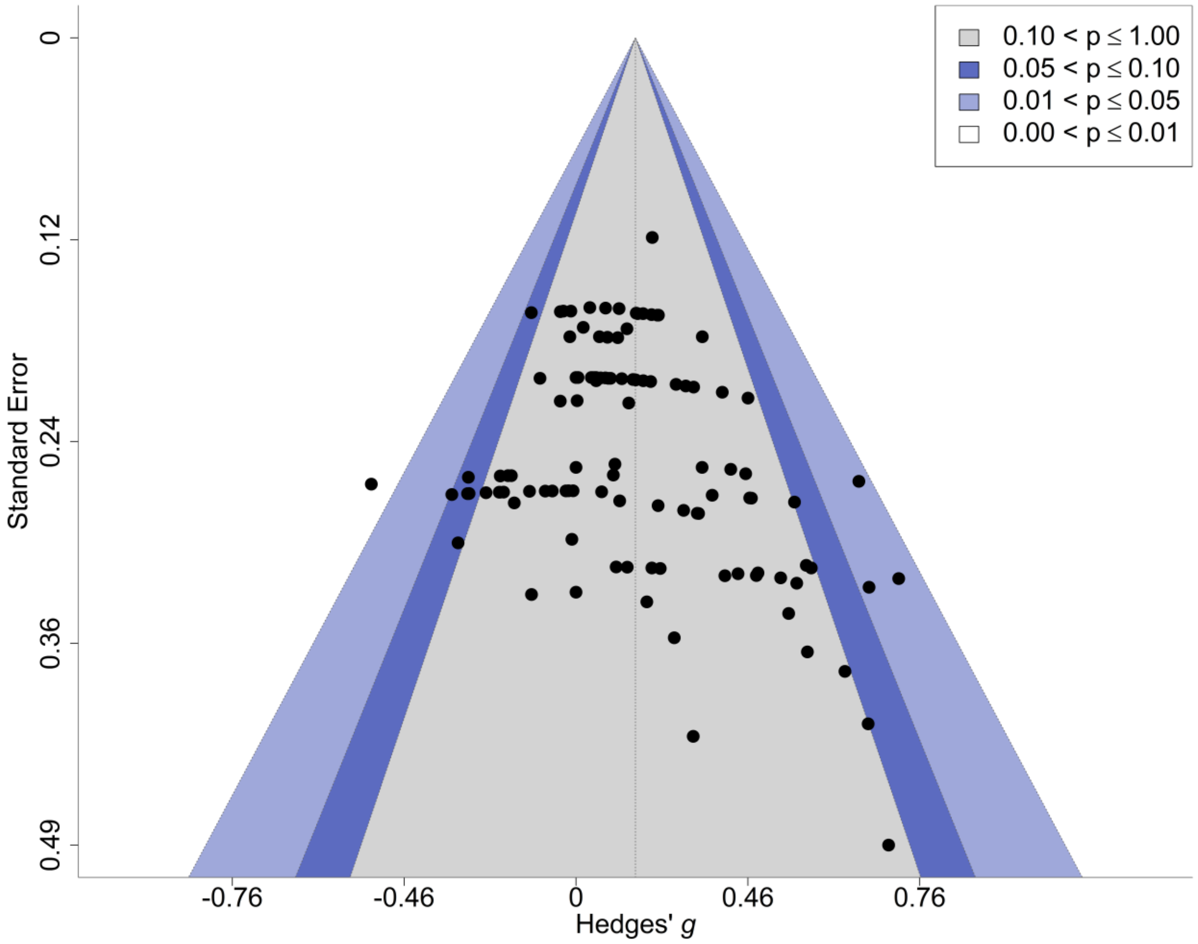

#### Neural MA

| | Full model | Model with outcome 34, 40, 41 and 43 removed |
|---|---|---|
| **Model fit** | | |
| AIC | 42.4251 | 12.9873 |
| BIC | 47.9755 | 18.2709 |
| **Heterogeneity** | | |
| QE | 139.8086 | 73.6844 |
| *p*-value | 0.0001 | 0.0025 |
| **Estimate** | | |
| Hedges' *g* | 0.2838 | 0.2323 |

| | | |
|---|---|---|
| *p*-value | 0.0021 | 0.0008 |

| Robust Bayesian Meta-Analysis (RoBMA) | | | | |
|---|---|---|---|---|
| | **Full model** | | **Outcome 34, 40, 41 and 43 removed** | |
| | **Least** | **Largest** | **Least** | **Largest** |
| **Inclusion BF** | | | | |
| Effect | 0.20 | 1.18 | 0.52 | 0.44 |
| Heterogeneity | 0.38 | 9.15 | 0.36 | 0.41 |
| Bias | 1.17 | 1.29 | 3.70 | 2.33 |
| **Mean estimates** | | | | |
| PET | 0.06 | 0.24 | 0.01 | 0.14 |
| PEESE | 0.15 | 0.43 | 0.05 | 0.41 |
| $\mu$ | 0.01 | 0.27 | -0.11 | 0.04 |

*Comparison of results of the multilevel meta-analysis (top) and RoBMA (bottom) results for neural outcomes. The results from with all eligible studies included is shown at the left, whereas the results with outliers removed is presented to the right.*

Egger's regression test for funnel plot asymmetry was non-significant, $t(32) = -0.55$, $p = 0.58$, with a negative bias estimate of -16.89 (SE = 30.49). This non-significant result suggests that, after excluding the outliers, there is no strong evidence of publication bias or small-study effects. The negative bias estimate value may indicate that smaller studies show more negative effect sizes, but this is not a robust finding given the large SE. The $tau^2$ value was 2341.16, indicating that there was still substantial residual heterogeneity among studies.

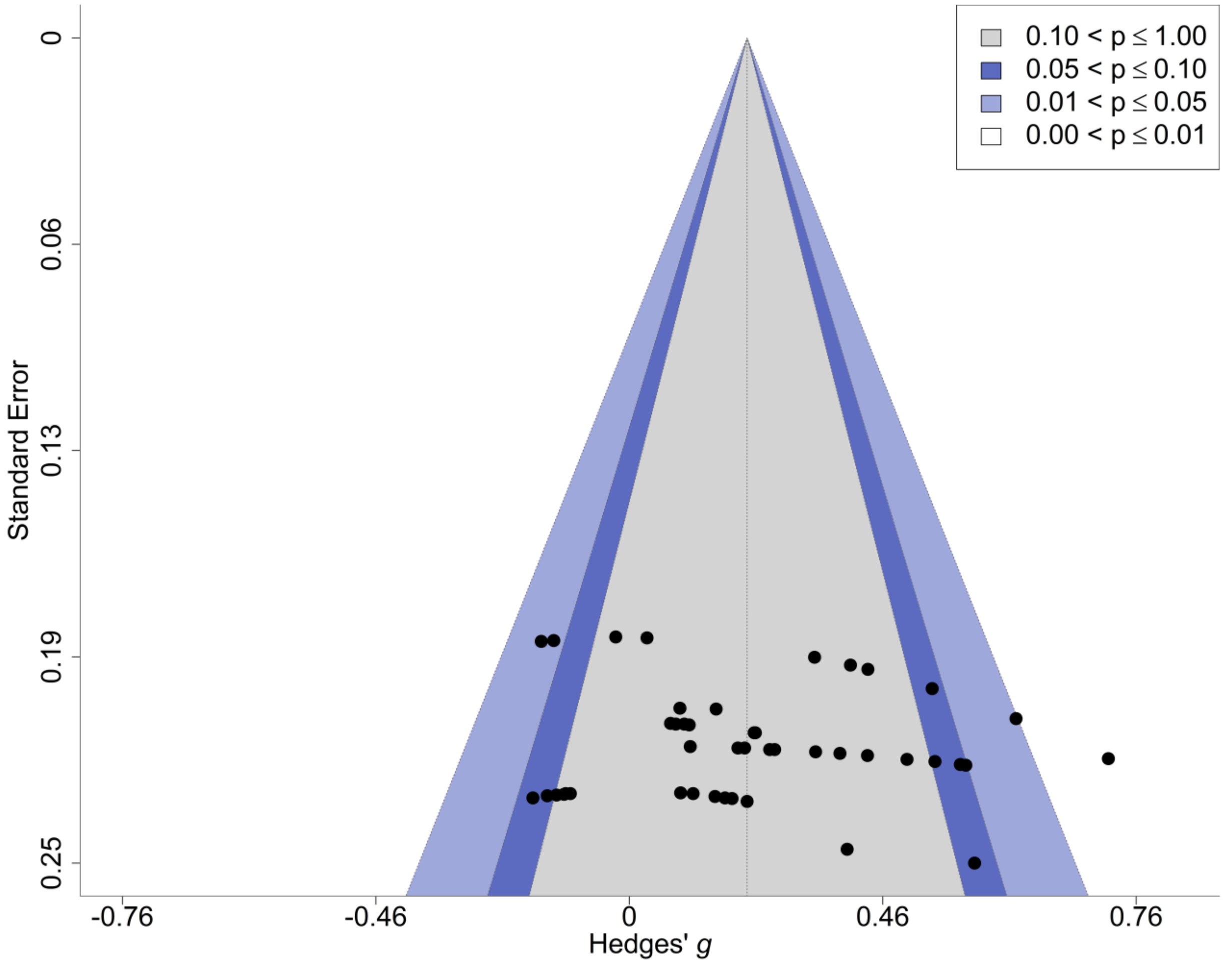
0.10 < p ≤ 1.00
0.05 < p ≤ 0.10
0.01 < p ≤ 0.05
0.00 < p ≤ 0.01
Standard Error
0
0.06
0.13
0.19
0.25
-0.76
-0.46
0
0.46
0.76
Hedges' g

### Protocol deviations

| Pre-printed protocol | Article |
|---|---|
| Defined two hypotheses | In the paper, these were rephrased and incorporated into text.<br><br>The hypotheses were defined before conducting the first literature search. One hypothesis mentioned EEG as a valid tool to detect modulations of neural activity after oxytocin administration. This was rephrased as the initial wording was unclear and gave the impression that EEG as a tool would be tested, instead of investigating the many ways in which EEG could be used to investigate oxytocin administration's effect on neural activity.<br><br>One hypothesis was that oxytocin administration would exert larger effects on neural activity in clinical than in neurotypical populations. However, the literature searched revealed that there simply weren't enough eligible studies to be able to make this comparison. |
| ESDist package would be used | Not used |
| Author contributions | Changes were made in terms of which author, besides ED, would do the data extraction, and the percentage of articles that would be screened by this author. |

### Initial literature search

dDatabase(s): **Ovid MEDLINE(R) ALL** 1946 to June 19, 2023
Search Strategy:

| # | Searches | Results |
|---|---|---|
| 1 | Oxytocin/ | 21538 |
| 2 | (ocytocin or oxytocin or pitocin or syntocinon or disipidin or endopituitrin or fetusin or hypophysin or hypophysine or mipareton or neoxyn or opn300 or orasthin or orastina or oxitocin or oxitone or oxoject or oxystin or oxytan or oxytocina or oxytocine or pareton or partacon or partocon or partolact or partoxin or physormon or piton or pituilobine or pitupartin or solvoxine or synpitan or tranoxy or utedrin or uteracon or uterason or vagitocin or xitocin or Oksytocin).tw,kw,kf. | 26589 |
| 3 | 1 or 2 | 30745 |
| 4 | Electroencephalography/ | 160998 |
| 5 | (eeg or electroencephalogram or electroencephalograms or electroencephalography or encephalography).tw,kw,kf. | 121127 |
| 6 | 4 or 5 | 197860 |
| 7 | Evoked Potentials/ | 55967 |
| 8 | ("n1 wave" or "n1 waves" or "n2 wave" or "n2 waves" or "n3 wave" or "n3 waves" or "n4 wave" or "n4 waves" or "p2 wave" or "p2 waves" or "p50 wave" or "p50 waves").tw,kw,kf. | 533 |
| 9 | (evoked adj3 (potential or potentials or discharge or response)).tw,kw,kf. | 62943 |
| 10 | (event adj3 related adj3 (potential or potentials or desynchronisation or desynchronization)).tw,kw,kf. | 25481 |
| 11 | or/7-10 | 118928 |
| 12 | Evoked Potentials, Visual/ | 17320 |
| 13 | (visual adj3 evoked adj3 (potential or response or responses)).tw,kw,kf. | 5349 |
| 14 | 12 or 13 | 19184 |
| 15 | 11 or 14 | 125229 |
| 16 | 6 or 15 | 284066 |
| 17 | 3 and 16 | 249 |

Database(s): **Embase Classic+Embase** 1947 to 2023 June 19
Search Strategy:

| # | Searches | Results |
|---|---|---|
| 1 | Oxytocin/ | 42272 |
| 2 | (ocytocin or oxytocin or pitocin or syntocinon or disipidin or endopituitrin or fetusin or hypophysin or hypophysine or mipareton or neoxyn or opn300 or orasthin or orastina or oxitocin or oxitone or oxoject or oxystin or oxytan or oxytocina or oxytocine or pareton or partacon or partocon or partolact or partoxin or physormon or piton or pituilobine or pitupartin or solvoxine or synpitan or tranoxy or utedrin or uteracon or uterason or vagitocin or xitocin or Oksytocin).tw,kw,kf. | 35502 |
| 3 | 1 or 2 | 48723 |
| 4 | Electroencephalography/ | 144297 |
| 5 | (eeg or electroencephalogram or electroencephalograms or electroencephalography or encephalography).tw,kw,kf. | 189812 |
| 6 | 4 or 5 | 256434 |
| 7 | evoked response/ | 47714 |
| 8 | ("n1 wave" or "n1 waves" or "n2 wave" or "n2 waves" or "n3 wave" or "n3 waves" or "n4 wave" or "n4 waves" or "p2 wave" or "p2 waves" or "p50 wave" or "p50 waves").tw,kw,kf. | 754 |
| 9 | (evoked adj3 (potential or potentials or discharge or response)).tw,kw,kf. | 85055 |
| 10 | (event adj3 related adj3 (potential or potentials or desynchronisation or desynchronization)).tw,kw,kf. | 31495 |
| 11 | or/7-10 | 138756 |
| 12 | visual evoked potential/ | 6241 |
| 13 | (visual adj3 evoked adj3 (potential or response or responses)).tw,kw,kf. | 7735 |
| 14 | 12 or 13 | 12103 |
| 15 | 11 or 14 | 141472 |
| 16 | 6 or 15 | 367816 |
| 17 | 3 and 16 | 389 |

Database(s): **APA PsycInfo** 1806 to June Week 2 2023
Search Strategy:

| # | Searches | Results |
|---|---|---|
| 1 | Oxytocin/ | 4003 |
| 2 | (ocytocin or oxytocin or pitocin or syntocinon or disipidin or endopituitrin or fetusin or hypophysin or hypophysine or mipareton or neoxyn or opn300 or orasthin or orastina or oxitocin or oxitone or oxoject or oxystin or oxytan or oxytocina or oxytocine or pareton or partacon or partocon or partolact or partoxin or physormon or piton or pituilobine or pitupartin or solvoxine or synpitan or tranoxy or utedrin or uteracon or uterason or vagitocin or xitocin or Oksytocin).ti,ab,id. | 5478 |
| 3 | 1 or 2 | 5497 |
| 4 | Electroencephalography/ | 31836 |
| 5 | (eeg or electroencephalogram or electroencephalograms or electroencephalography or encephalography).ti,ab,id. | 51595 |
| 6 | 4 or 5 | 53483 |
| 7 | Evoked Potentials/ or Visual Evoked Potentials/ | 29909 |
| 8 | ("n1 wave" or "n1 waves" or "n2 wave" or "n2 waves" or "n3 wave" or "n3 waves" or "n4 wave" or "n4 waves" or "p2 wave" or "p2 waves" or "p50 wave" or "p50 waves").ti,ab,id. | 202 |
| 9 | (evoked adj3 (potential or potentials or discharge or response)).ti,ab,id. | 19406 |
| 10 | (event adj3 related adj3 (potential or potentials or desynchronisation or desynchronization)).ti,ab,id. | 23329 |
| 11 | (visual adj3 evoked adj3 (potential or response or responses)).ti,ab,id. | 1887 |
| 12 | or/7-11 | 47558 |
| 13 | 6 or 12 | 90962 |
| 14 | 3 and 13 | 74 |

Search Name: **COCHRANE**

Date Run: 20/06/2023 20:18:08

Comment:

ID Search Hits

#2 (ocytocin OR oxytocin OR pitocin OR syntocinon OR disipidin OR endopituitrin OR fetusin OR hypophysin OR hypophysine OR mipareton OR neoxyn OR opn300 OR orasthin OR orastina OR oxitocin OR oxitone OR oxoject OR oxystin OR oxytan OR oxytocina OR oxytocine OR pareton OR partacon OR partocon OR partolact OR partoxin OR physormon OR piton OR pituilobine OR pitupartin OR solvoxine OR synpitan OR tranoxy OR utedrin OR uteracon OR uterason OR vagitocin OR xitocin):ti,ab,kw 5903

#3 (eeg OR electroencephalogram OR electroencephalograms OR electroencephalography OR encephalography):ti,ab,kw 13944

#4 (n1 wave OR n1 waves OR n2 wave OR n2 waves OR n3 wave OR n3 waves OR n4 wave OR n4 waves OR p2 wave OR p2 waves OR p50 wave OR p50 waves OR wave n1 OR wave n2 OR wave n3 OR wave n4 OR wave p2 OR wave p50 OR waves n1 OR waves n2 OR waves n3 OR waves n4 OR waves p2 OR waves p50):ti,ab,kw 252

#5 (evoked) NEAR/3 (potential OR potentials OR discharge OR response):ti,ab,kw 6612

#6 (event) NEAR/3 (related) NEAR/3 (potential OR potentials OR desynchronisation OR desynchronization):ti,ab,kw 2213

#7 (visual) NEAR/3 (evoked) NEAR/3 (potential OR response OR responses):ti,ab,kw 500

#8 #3 or #4 or #5 or #6 or #7 19333

#9 #2 and #8 91

**CINAHL**

Tuesday, June 20, 2023 12:52:48 PM

### Query Limiters/Expanders Last Run Via Results

S3 S1 AND S2 Expanders - Apply equivalent subjects

Search modes - Boolean/Phrase Interface - EBSCOhost Research Databases

Search Screen - Advanced Search

Database - CINAHL 18

S2 ( (MH "Evoked Potentials+") OR (MH "Evoked Potentials, Visual") ) OR (MH "Electroencephalography") ) OR ( (evoked) N3 (potential OR potentials OR discharge OR response) ) OR ( (event related) N3 (potential OR potentials OR desynchronisation OR desynchronization) ) AND ( (visual evoked) N3 (potential OR response OR responses) ) AND ( "n1 wave" or "n1 waves" or "n2 wave" or "n2 waves" or "n3 wave" or "n3 waves" or "n4 wave" or "n4 waves" or "p2 wave" or "p2 waves" or "p50 wave" or "p50 waves" ) Expanders - Apply equivalent subjects

Search modes - Boolean/Phrase Interface - EBSCOhost Research Databases

Search Screen - Advanced Search

Database - CINAHL 32,271

S1 (MH "Oxytocin") OR ( ocytocin or oxytocin or pitocin or syntocinon or disipidin or endopituitrin or fetusin or hypophysin or hypophysine or mipareton or neoxyn or opn300 or orasthin or orastina or oxitocin or oxitone or oxoject or oxystin or oxytan or oxytocina or oxytocine or pareton or partacon or partocon or partolact or partoxin or physormon or piton or pituilobine or pitupartin or solvoxine or synpitan or tranoxy or utedrin or uteracon or uterason or vagitocin or xitocin or Oksytocin ) Expanders - Apply equivalent subjects

Search modes - Boolean/Phrase Interface - EBSCOhost Research Databases

Search Screen - Advanced Search

Database - CINAHL 4,28

**SCOPUS**

Tuesday, June 20, 2023

( TITLE-ABS-KEY ( ocytocin OR oxytocin OR pitocin OR syntocinon OR disipidin OR endopituitrin OR fetusin OR hypophysin OR hypophysine OR mipareton OR neoxyn OR opn300 OR orasthin OR orastina OR oxitocin OR oxitone OR oxoject OR oxystin OR oxytan OR oxytocina OR oxytocine OR pareton OR partacon OR partocon OR partolact OR partoxin OR physormon OR piton OR pituilobine OR pitupartin OR solvoxine OR synpitan

OR tranoxy OR utedrin OR uteracon OR uterason OR vagitocin OR xitocin OR oksytocin ) ) AND ( ( TITLE-ABS-KEY ( eeg OR electroencephalogram OR electroencephalograms OR electroencephalography OR encephalography ) OR TITLE-ABS-KEY ( "n1 wave" OR "n1 waves" OR "n2 wave" OR "n2 waves" OR "n3 wave" OR "n3 waves" OR "n4 wave" OR "n4 waves" OR "p2 wave" OR "p2 waves" OR "p50 wave" OR "p50 waves" ) OR TITLE-ABS-KEY ( ( evoked ) W/3 ( potential OR potentials OR discharge OR response ) ) OR TITLE-ABS-KEY ( ( "event related" ) W/3 ( potential OR potentials OR desynchronisation OR desynchronization ) ) OR TITLE-ABS-KEY ( ( "visual evoked" ) W/3 ( potential OR response OR responses ) ) ) )

Results: 699

**WEB OF SCIENCE**

Tuesday, June 20, 2023

Query #1

ocytocin OR oxytocin OR pitocin OR syntocinon OR disipidin OR endopituitrin OR fetusin OR hypophysin OR hypophysine OR mipareton OR neoxyn OR opn300 OR orasthin OR orastina OR oxitocin OR oxitone OR oxoject OR oxystin OR oxytan OR oxytocina OR oxytocine OR pareton OR partacon OR partocon OR partolact OR partoxin OR physormon OR piton OR pituilobine OR pitupartin OR solvoxine OR synpitan OR tranoxy OR utedrin OR uteracon OR uterason OR vagitocin OR xitocin (Topic)

Query #2

"n1 wave" OR "evoked potential" OR "event related potential" OR "n1 waves" OR "evoked potentials" OR "event related potentials" OR "n2 wave" OR "evoked discharge" OR "event related desynchronisation" OR "n2 waves" OR "evoked potentials" OR "event related desynchronization" OR "n3 wave" OR "evoked response" OR "n3 waves" OR "n4 wave" OR "n4 waves" OR "p2 wave" OR "p2 waves" OR "p50 wave" OR "p50 waves" OR "wave n1" OR "wave n2" OR "wave n3" OR "wave n4" OR "wave p2" OR "wave p50" OR "waves n1" OR "waves n2" OR "waves n3" OR "waves n4" OR "waves p2" OR "waves p50" (Topic) or (evoked) NEAR/3 (potential OR potentials OR discharge OR response) (Topic) or ("event related") NEAR/3 (potential OR potentials OR desynchronisation OR desynchronization) (Topic) or ("visual evoked") NEAR/3 (potential OR response OR responses) (Topic) or eeg OR electroencephalogram OR electroencephalograms OR electroencephalography OR encephalography (Topic)

#1 AND #2

Results: 299

Epistemonikos

(title:((title:(ocytocin OR oxytocin OR pitocin OR syntocinon OR disipidin OR endopituitrin OR fetusin OR hypophysin OR hypophysine OR mipareton OR neoxyn OR opn300 OR orasthin OR orastina OR oxitocin OR oxitone OR oxoject OR oxystin OR oxytan OR oxytocina OR oxytocine OR pareton OR partacon OR partocon OR partolact OR partoxin OR physormon OR pitocin OR piton OR pituilobine OR pitupartin OR solvoxine OR synpitan OR syntocinon OR tranoxy OR utedrin OR uteracon OR uterason OR vagitocin OR xitocin) OR abstract:(ocytocin OR oxytocin OR pitocin OR syntocinon OR disipidin OR endopituitrin OR fetusin OR hypophysin OR hypophysine OR mipareton OR neoxyn OR opn300 OR orasthin OR orastina OR oxitocin OR oxitone OR oxoject OR oxystin OR oxytan OR oxytocina OR oxytocine OR pareton OR partacon OR partocon OR partolact OR partoxin OR physormon OR pitocin OR piton OR pituilobine OR pitupartin OR solvoxine OR synpitan OR syntocinon OR tranoxy OR utedrin OR uteracon OR uterason OR vagitocin OR xitocin)) AND (title:("eeg" OR "electroencephalogram" OR "electroencephalograms" OR "electroencephalography" OR "encephalography" OR "n1 wave" OR "evoked potential" OR "event related potential" OR "n1 waves" OR "evoked potentials" OR "event related potentials" OR "n2 wave" OR "evoked discharge" OR "event related desynchronisation" OR "n2 waves" OR "evoked potentials" OR "event related desynchronization" OR "n3 wave" OR "evoked response" OR "n3 waves" OR "n4 wave" OR "n4 waves" OR "p2 wave" OR "p2 waves" OR "p50 wave" OR "p50 waves" OR "wave n1" OR "wave n2" OR "wave n3" OR "wave n4" OR "wave p2" OR "wave p50" OR "waves n1" OR "waves n2" OR "waves n3" OR "waves n4" OR "waves p2" OR "waves p50" OR "visual evoked potential*" OR "visual evoked response" OR "visual evoked responses") OR abstract:("eeg" OR "electroencephalogram" OR "electroencephalograms" OR "electroencephalography" OR "encephalography" OR "n1 wave" OR "evoked potential" OR "event related potential" OR "n1 waves" OR "evoked potentials" OR "event related potentials" OR "n2 wave" OR "evoked discharge" OR "event related desynchronisation" OR "n2 waves" OR "evoked potentials" OR "event related desynchronization" OR "n3 wave" OR "evoked response" OR "n3 waves" OR "n4 wave" OR "n4 waves" OR "p2 wave" OR "p2 waves" OR "p50 wave" OR "p50 waves" OR "wave n1" OR "wave n2" OR "wave n3" OR "wave n4" OR "wave p2" OR "wave p50" OR "waves n1" OR "waves n2" OR "waves n3" OR "waves n4" OR "waves p2" OR "waves p50" OR "visual evoked potential*" OR "visual evoked response" OR "visual evoked responses"))) OR abstract:((title:(ocytocin OR oxytocin OR pitocin OR syntocinon OR disipidin OR endopituitrin OR fetusin OR hypophysin OR hypophysine OR mipareton OR neoxyn OR opn300 OR orasthin OR orastina OR oxitocin OR oxitone OR oxoject OR oxystin OR oxytan OR oxytocina OR oxytocine OR pareton OR partacon OR partocon OR partolact OR partoxin OR physormon OR pitocin OR piton OR pituilobine OR pitupartin OR solvoxine OR synpitan OR syntocinon OR tranoxy OR utedrin OR uteracon OR uterason OR vagitocin OR xitocin) OR abstract:(ocytocin OR oxytocin OR pitocin OR syntocinon OR disipidin OR endopituitrin OR fetusin OR hypophysin OR hypophysine OR mipareton OR neoxyn OR opn300 OR orasthin OR orastina OR oxitocin OR oxitone OR oxoject OR oxystin OR oxytan OR oxytocina OR oxytocine OR pareton OR partacon OR partocon OR partolact OR partoxin OR physormon OR pitocin OR piton OR pituilobine OR pitupartin OR solvoxine OR synpitan OR syntocinon OR tranoxy OR utedrin OR uteracon OR uterason OR vagitocin OR xitocin)) AND (title:("eeg" OR "electroencephalogram" OR "electroencephalograms" OR "electroencephalography" OR "encephalography" OR "n1 wave" OR "evoked potential" OR "event related potential" OR "n1 waves" OR "evoked potentials" OR "event related potentials" OR "n2 wave" OR "evoked discharge" OR "event related desynchronisation" OR "n2 waves" OR "evoked potentials" OR "event related desynchronization" OR "n3 wave" OR "evoked response" OR "n3 waves" OR "n4 wave" OR "n4 waves" OR "p2 wave" OR "p2 waves" OR "p50 wave" OR "p50 waves" OR "wave n1" OR "wave n2" OR "wave n3" OR "wave n4" OR "wave p2" OR "wave p50" OR "waves n1" OR "waves n2" OR "waves n3" OR "waves n4" OR "waves p2" OR "waves p50" OR "visual evoked potential*" OR "visual evoked response" OR "visual evoked responses") OR abstract:("eeg" OR "electroencephalogram" OR "electroencephalograms" OR "electroencephalography" OR "encephalography" OR "n1 wave" OR "evoked potential" OR "event related potential" OR "n1 waves" OR "evoked potentials" OR "event related potentials" OR "n2 wave" OR "evoked discharge" OR "event related desynchronisation" OR "n2 waves" OR "evoked potentials" OR "event related desynchronization" OR "n3 wave" OR "evoked response" OR "n3 waves" OR "n4 wave" OR "n4 waves" OR "p2 wave" OR "p2 waves" OR "p50 wave" OR "p50 waves" OR "wave n1" OR "wave n2" OR "wave n3" OR "wave n4" OR "wave p2" OR "wave p50" OR "waves n1" OR "waves n2" OR "waves n3" OR "waves n4" OR "waves p2" OR "waves p50" OR "visual evoked potential*" OR "visual evoked response" OR "visual evoked responses"))))

Results: 13

### Second literature search

Database(s): **Ovid MEDLINE(R) ALL** 1946 to July 22, 2024
Search Strategy:

| # | Searches | Results |
|---|---|---|
| 1 | Oxytocin/ | 22003 |
| 2 | (ocytocin or oxytocin or pitocin or syntocinon or disipidin or endopituitrin or fetusin or hypophysin or hypophysine or mipareton or neoxyn or opn300 or orasthin or orastina or oxitocin or oxitone or oxoject or oxystin or oxytan or oxytocina or oxytocine or pareton or partacon or partocon or partolact or partoxin or physormon or piton or pituilobine or pitupartin or solvoxine or synpitan or tranoxy or utedrin or uteracon or uterason or vagitocin or xitocin or Oksytocin).tw,kw,kf. | 27571 |
| 3 | 1 or 2 | 31735 |
| 4 | Electroencephalography/ | 165652 |
| 5 | (eeg or electroencephalogram or electroencephalograms or electroencephalography or encephalography).tw,kw,kf. | 128888 |
| 6 | 4 or 5 | 206520 |
| 7 | Evoked Potentials/ | 56825 |
| 8 | ("n1 wave" or "n1 waves" or "n2 wave" or "n2 waves" or "n3 wave" or "n3 waves" or "n4 wave" or "n4 waves" or "p2 wave" or "p2 waves" or "p50 wave" or "p50 waves").tw,kw,kf. | 552 |
| 9 | (evoked adj3 (potential or potentials or discharge or response)).tw,kw,kf. | 64807 |
| 10 | (event adj3 related adj3 (potential or potentials or desynchronisation or desynchronization)).tw,kw,kf. | 26738 |
| 11 | or/7-10 | 122122 |
| 12 | Evoked Potentials, Visual/ | 17596 |
| 13 | (visual adj3 evoked adj3 (potential or response or responses)).tw,kw,kf. | 5587 |
| 14 | 12 or 13 | 19551 |
| 15 | 6 or 11 or 14 | 294498 |
| 16 | 3 and 15 | 262 |
| 17 | limit 16 to yr="2023 -Current" | 15 |

Database(s): **Embase Classic+Embase** 1947 to 2024 July 22
Search Strategy:

| # | Searches | Results |
|---|---|---|
| 1 | Oxytocin/ | 43840 |
| 2 | (ocytocin or oxytocin or pitocin or syntocinon or disipidin or endopituitrin or fetusin or hypophysin or hypophysine or mipareton or neoxyn or opn300 or orasthin or orastina or oxitocin or oxitone or oxoject or oxystin or oxytan or oxytocina or oxytocine or pareton or partacon or partocon or partolact or partoxin or physormon or piton or pituilobine or pitupartin or solvoxine or synpitan or tranoxy or utedrin or uteracon or uterason or vagitocin or xitocin or Oksytocin).tw,kw,kf. | 36536 |
| 3 | 1 or 2 | 50493 |
| 4 | Electroencephalography/ | 155183 |
| 5 | (eeg or electroencephalogram or electroencephalograms or electroencephalography or encephalography).tw,kw,kf. | 198877 |
| 6 | 4 or 5 | 269263 |
| 7 | evoked response/ | 49050 |
| 8 | ("n1 wave" or "n1 waves" or "n2 wave" or "n2 waves" or "n3 wave" or "n3 waves" or "n4 wave" or "n4 waves" or "p2 wave" or "p2 waves" or "p50 wave" or "p50 waves").tw,kw,kf. | 775 |
| 9 | (evoked adj3 (potential or potentials or discharge or response)).tw,kw,kf. | 86942 |
| 10 | (event adj3 related adj3 (potential or potentials or desynchronisation or desynchronization)).tw,kw,kf. | 32520 |
| 11 | or/7-10 | 142116 |
| 12 | visual evoked potential/ | 6996 |
| 13 | (visual adj3 evoked adj3 (potential or response or responses)).tw,kw,kf. | 7968 |
| 14 | 12 or 13 | 12876 |
| 15 | 6 or 11 or 14 | 382890 |
| 16 | 3 and 15 | 408 |
| 17 | limit 16 to yr="2023 -Current" | 28 |

Database(s): **APA PsycInfo** 1806 to July Week 3 2024
Search Strategy:

| # | Searches | Results |
|---|---|---|
| 1 | Oxytocin/ | 4224 |
| 2 | (ocytocin or oxytocin or pitocin or syntocinon or disipidin or endopituitrin or fetusin or hypophysin or hypophysine or mipareton or neoxyn or opn300 or orasthin or orastina or oxitocin or oxitone or oxoject or oxystin or oxytan or oxytocina or oxytocine or pareton or partacon or partocon or partolact or partoxin or physormon or piton or pituilobine or pitupartin or solvoxine or synpitan or tranoxy or utedrin or uteracon or uterason or vagitocin or xitocin or Oksytocin).ti,ab,id. | 5750 |
| 3 | 1 or 2 | 5770 |
| 4 | Electroencephalography/ | 34016 |
| 5 | (eeg or electroencephalogram or electroencephalograms or electroencephalography or encephalography).ti,ab,id. | 54565 |
| 6 | 4 or 5 | 56533 |
| 7 | Evoked Potentials/ or Visual Evoked Potentials/ | 31016 |
| 8 | ("n1 wave" or "n1 waves" or "n2 wave" or "n2 waves" or "n3 wave" or "n3 waves" or "n4 wave" or "n4 waves" or "p2 wave" or "p2 waves" or "p50 wave" or "p50 waves").ti,ab,id. | 207 |
| 9 | (evoked adj3 (potential or potentials or discharge or response)).ti,ab,id. | 19864 |
| 10 | (event adj3 related adj3 (potential or potentials or desynchronisation or desynchronization)).ti,ab,id. | 24279 |
| 11 | (visual adj3 evoked adj3 (potential or response or responses)).ti,ab,id. | 1932 |
| 12 | or/7-11 | 49091 |
| 13 | 6 or 12 | 94887 |
| 14 | 3 and 13 | 81 |
| 15 | limit 14 to yr="2023 -Current" | 9 |

Search Name:

Date Run: 23/07/2024 08:40:38

Comment:

ID Search Hits

#1 (ocytocin OR oxytocin OR pitocin OR syntocinon OR disipidin OR endopituitrin OR fetusin OR hypophysin OR hypophysine OR mipareton OR neoxyn OR opn300 OR orasthin OR orastina OR oxitocin OR oxitone OR oxoject OR oxystin OR oxytan OR oxytocina OR oxytocine OR pareton OR partacon OR partocon OR partolact OR partoxin OR physormon OR piton OR pituilobine OR pitupartin OR solvoxine OR synpitan OR tranoxy OR utedrin OR uteracon OR uterason OR vagitocin OR xitocin):ti,ab,kw 6380

#2 eeg OR electroencephalogram OR electroencephalograms OR electroencephalography OR encephalography 16107

#3 n1 wave OR n1 waves OR n2 wave OR n2 waves OR n3 wave OR n3 waves OR n4 wave OR n4 waves OR p2 wave OR p2 waves OR p50 wave OR p50 waves OR wave n1 OR wave n2 OR wave n3 OR wave n4 OR wave p2 OR wave p50 OR waves n1 OR waves n2 OR waves n3 OR waves n4 OR waves p2 OR waves p50 408

#4 (evoked) NEAR/3 (potential OR potentials OR discharge OR response) 7405

#5 (event) NEAR/3 (related) NEAR/3 (potential OR potentials OR desynchronisation OR desynchronization) 2400

#6 (visual) NEAR/3 (evoked) NEAR/3 (potential OR response OR responses) 561

#7 #2 or #3 or #4 or #5 or #6 22061

#8 #1 and #7 105

Scopus 23.07.2024

( TITLE-ABS-KEY ( ocytocin OR oxytocin OR pitocin OR syntocinon OR disipidin OR endopituitrin OR fetusin OR hypophysin OR hypophysine OR mipareton OR neoxyn OR opn300 OR orasthin OR orastina OR oxitocin OR oxitone OR oxoject OR oxystin OR oxytan OR oxytocina OR oxytocine OR pareton OR partacon OR partocon OR partolact OR partoxin OR physormon OR piton OR pituilobine OR pitupartin OR solvoxine OR synpitan OR tranoxy OR utedrin OR uteracon OR uterason OR vagitocin OR xitocin OR oksytocin ) ) AND ( ( TITLE-ABS-KEY ( eeg OR electroencephalogram OR electroencephalograms OR electroencephalography OR encephalography OR "n1 wave" OR "n1 waves" OR "n2 wave" OR "n2 waves" OR "n3 wave" OR "n3 waves" OR "n4 wave" OR "n4 waves" OR "p2 wave" OR "p2 waves" OR "p50 wave" OR "p50 waves" ) ) OR ( TITLE-ABS-KEY ( ( ( evoked ) W/3 ( potential OR potentials OR discharge OR response ) ) ) ) OR ( TITLE-ABS-KEY ( ( ( "event related" ) W/3 ( potential OR potentials OR desynchronisation OR desynchronization ) ) ) ) OR ( TITLE-ABS-KEY ( ( ( "visual evoked" ) W/3 ( potential OR response OR responses ) ) ) ) ) AND PUBYEAR > 2022 AND PUBYEAR < 2025

**CINAHL**

23.07.2024

( ( (MH "Evoked Potentials+") OR (MH "Evoked Potentials, Visual") ) OR (MH "Electroencephalography") ) OR ( (evoked) N3 (potential OR potentials OR discharge OR response) ) OR ( (event related) N3 (potential OR potentials OR desynchronisation OR desynchronization) ) or ( (visual evoked) N3 (potential OR response OR responses) ) or ( "n1 wave" or "n1 waves" or "n2 wave" or "n2 waves" or "n3 wave" or "n3 waves" or "n4 wave" or "n4 waves" or "p2 wave" or "p2 waves" or "p50 wave" or "p50 waves" ) ) AND ( (MH "Oxytocin") OR ( ocytocin or oxytocin or pitocin or syntocinon or disipidin or endopituitrin or fetusin or hypophysin or hypophysine or mipareton or neoxyn or opn300 or orasthin or orastina or oxitocin or oxitone or oxoject or oxystin or oxytan or oxytocina or oxytocine or pareton or partacon or partocon or partolact or partoxin or physormon or piton or pituilobine or pitupartin or solvoxine or synpitan or tranoxy or utedrin or uteracon or uterason or vagitocin or xitocin or Oksytocin ) )

**WEB OF SCIENCE**

Tuesday, July 23, 2024

Query #1

ocytocin OR oxytocin OR pitocin OR syntocinon OR disipidin OR endopituitrin OR fetusin OR hypophysin OR hypophysine OR mipareton OR neoxyn OR opn300 OR orasthin OR orastina OR oxitocin OR oxitone OR oxoject OR oxystin OR oxytan OR oxytocina OR oxytocine OR pareton OR partacon OR partocon OR partolact OR partoxin OR physormon OR piton OR pituilobine OR pitupartin OR solvoxine OR synpitan OR tranoxy OR utedrin OR uteracon OR uterason OR vagitocin OR xitocin (Topic)

Query #2

"n1 wave" OR "evoked potential" OR "event related potential" OR "n1 waves" OR "evoked potentials" OR "event related potentials" OR "n2 wave" OR "evoked discharge" OR "event related desynchronisation" OR "n2 waves" OR "evoked potentials" OR "event related desynchronization" OR "n3 wave" OR "evoked response" OR "n3 waves" OR "n4 wave" OR "n4 waves" OR "p2 wave" OR "p2 waves" OR "p50 wave" OR "p50 waves" OR "wave n1" OR "wave n2" OR "wave n3" OR "wave n4" OR "wave p2" OR "wave p50" OR "waves n1" OR "waves n2" OR "waves n3" OR "waves n4" OR "waves p2" OR "waves p50" (Topic) or (evoked) NEAR/3 (potential OR potentials OR discharge OR response) (Topic) or ("event related") NEAR/3 (potential OR potentials OR desynchronisation OR desynchronization) (Topic) or ("visual evoked") NEAR/3 (potential OR response OR responses) (Topic) or eeg OR electroencephalogram OR electroencephalograms OR electroencephalography OR encephalography (Topic)

#1 AND #2

Results: 299

**EPISTEMONIKOS**

23/07/2024

ocytocin OR oxytocin OR pitocin OR syntocinon OR disipidin OR endopituitrin OR fetusin OR hypophysin OR hypophysine OR mipareton OR neoxyn OR opn300 OR orasthin OR orastina OR oxitocin OR oxitone OR oxoject OR oxystin OR oxytan OR oxytocina OR oxytocine OR pareton OR partacon OR partocon OR partolact OR partoxin OR physormon OR pitocin OR piton OR pituilobine OR pitupartin OR solvoxine OR synpitan OR syntocinon OR tranoxy OR utedrin OR uteracon OR uterason OR vagitocin OR xitocin) OR abstract:(ocytocin OR oxytocin OR pitocin OR syntocinon OR disipidin OR endopituitrin OR fetusin OR hypophysin OR hypophysine OR mipareton OR neoxyn OR opn300 OR orasthin OR orastina OR oxitocin OR oxitone OR oxoject OR oxystin OR oxytan OR oxytocina OR oxytocine OR pareton OR partacon OR partocon OR partolact OR partoxin OR physormon OR pitocin OR piton OR pituilobine OR pitupartin OR solvoxine OR synpitan OR syntocinon OR tranoxy OR utedrin OR uteracon OR uterason OR vagitocin OR xitocin

AND

"eeg" OR "electroencephalogram" OR "electroencephalograms" OR "electroencephalography" OR "encephalography" OR "n1 wave" OR "evoked potential" OR "event related potential" OR "n1 waves" OR "evoked potentials" OR "event related potentials" OR "n2 wave" OR "evoked discharge" OR "event related desynchronisation" OR "n2 waves" OR "evoked potentials" OR "event related desynchronization" OR "n3 wave" OR "evoked response" OR "n3 waves" OR "n4 wave" OR "n4 waves" OR "p2 wave" OR "p2 waves" OR "p50 wave" OR "p50 waves" OR "wave n1" OR "wave n2" OR "wave n3" OR "wave n4" OR "wave p2" OR "wave p50" OR "waves n1" OR "waves n2" OR "waves n3" OR "waves n4" OR "waves p2" OR "waves p50" OR "visual evoked potential*" OR "visual evoked response" OR "visual evoked responses"

### References

Alaerts, K., Moerkerke, M., Daniels, N., Zhang, Q., Grazia, R., Steyaert, J., Prinsen, J., & Boets, B. (2024). Chronic oxytocin improves neural decoupling at rest in children with autism: An exploratory RCT. *Journal of Child Psychology and Psychiatry*, *65*(10), 1311–1326. https://doi.org/10.1111/jcpp.13966

Alaerts, K., Taillieu, A., Daniels, N., Soriano, J. R., & Prinsen, J. (2021). Oxytocin enhances neural approach towards social and non-social stimuli of high personal relevance. *Scientific Reports*, *11*(1), 23589. https://doi.org/10.1038/s41598-021-02914-8

Alaerts, K., Taillieu, A., Prinsen, J., & Daniels, N. (2021). Tracking transient changes in the intrinsic neural frequency architecture: Oxytocin facilitates non-harmonic relationships between alpha and theta rhythms in the resting brain. *Psychoneuroendocrinology*, *133*, 105397. https://doi.org/10.1016/j.psyneuen.2021.105397

de Bruijn, E. R. A., Ruissen, M. I., & Radke, S. (2017). Electrophysiological correlates of oxytocin-induced enhancement of social performance monitoring. *Social Cognitive and Affective Neuroscience*, *12*(10), 1668–1677. https://doi.org/10.1093/scan/nsx094

Festante, F., Ferrari, P. F., Thorpe, S. G., Buchanan, R. W., & Fox, N. A. (2020). Intranasal oxytocin enhances EEG mu rhythm desynchronization during execution and observation of social action: An exploratory study. *Psychoneuroendocrinology*, *111*, 104467. https://doi.org/10.1016/j.psyneuen.2019.104467

Higgins, J. P. T., & Thompson, S. G. (2002). Quantifying heterogeneity in a meta-analysis. *Statistics in Medicine*, *21*(11), 1539–1558. https://doi.org/10.1002/sim.1186

Moerkerke, M., Daniels, N., Van der Donck, S., Tibermont, L., Tang, T., Debbaut, E., Bamps, A., Prinsen, J., Steyaert, J., Alaerts, K., & Boets, B. (2023). Can repeated intranasal oxytocin administration affect reduced neural sensitivity towards expressive faces in autism? A randomized controlled trial. *Journal of Child Psychology and Psychiatry*, *64*(11), 1583–1595. https://doi.org/10.1111/jcpp.13850

Moses, E., Nelson, N., Taubert, J., & Pegna, A. J. (2024). Oxytocin differentially modulates the early neural responses to faces and non-social stimuli. *Social Cognitive and Affective Neuroscience*, *19*(1), nsae010. https://doi.org/10.1093/scan/nsae010

Mu, Y., Guo, C., & Han, S. (2016). Oxytocin enhances inter-brain synchrony during social coordination in male adults. *Social Cognitive and Affective Neuroscience*, *11*(12), 1882–1893. https://doi.org/10.1093/scan/nsw106

Ochiai, H., Shiga, T., Hoshino, H., Horikoshi, S., Kanno, K., Wada, T., Osakabe, Y., Miura, I., & Yabe, H. (2021). Effect of oxytocin nasal spray on auditory automatic discrimination measured by mismatch negativity. *Psychopharmacology*, *238*(7), 1781–1789. https://doi.org/10.1007/s00213-021-05807-w

Paloyelis, Y., Krahé, C., Maltezos, S., Williams, S. C., Howard, M. A., & Fotopoulou, A. (2016). The Analgesic Effect of Oxytocin in Humans: A Double-Blind, Placebo-Controlled Cross-Over Study Using Laser-Evoked Potentials. *Journal of Neuroendocrinology*, *28*(4). https://doi.org/10.1111/jne.12347

Peltola, M. J., Strathearn, L., & Puura, K. (2018). Oxytocin promotes face-sensitive neural responses to infant and adult faces in mothers. *Psychoneuroendocrinology*, *91*, 261–270. https://doi.org/10.1016/j.psyneuen.2018.02.012

Perry, A., Bentin, S., Shalev, I., Israel, S., Uzefovsky, F., Bar-On, D., & Ebstein, R. P. (2010). Intranasal oxytocin modulates EEG mu/alpha and beta rhythms during perception of biological motion. *Psychoneuroendocrinology*, *35*(10), 1446–1453. https://doi.org/10.1016/j.psyneuen.2010.04.011

Petereit, P., Rinn, C., Stemmler, G., & Mueller, E. M. (2019). Oxytocin reduces the link between neural and affective responses after social exclusion. *Biological Psychology*, *145*, 224–235. https://doi.org/10.1016/j.biopsycho.2019.05.002

Qiao, Z., Van der Donck, S., Moerkerke, M., Dlhosova, T., Vettori, S., Dzhelyova, M., van Winkel, R., Alaerts, K., & Boets, B. (2022). Frequency-Tagging EEG of Superimposed Social and

Non-Social Visual Stimulation Streams Provides No Support for Social Salience Enhancement after Intranasal Oxytocin Administration. *Brain Sciences*, *12*(9). https://doi.org/10.3390/brainsci12091224

Rutherford, H. J. V., Guo, X. M., Graber, K. M., Hayes, N. J., Pelphrey, K. A., & Mayes, L. C. (2017). Intranasal oxytocin and the neural correlates of infant face processing in non-parent women. *Biological Psychology*, *129*, 45–48. https://doi.org/10.1016/j.biopsycho.2017.08.002

Rutherford, H. J. V., Guo, X. M., Wu, J., Graber, K. M., Hayes, N. J., Pelphrey, K. A., & Mayes, L. C. (2018). Intranasal oxytocin decreases cross-frequency coupling of neural oscillations at rest. *International Journal of Psychophysiology*, *123*, 143–151. https://doi.org/10.1016/j.ijpsycho.2017.09.017

Santiago, A. F., Kosilo, M., Cogoni, C., Diogo, V., Jerónimo, R., & Prata, D. (2024). Oxytocin modulates neural activity during early perceptual salience attribution. *Psychoneuroendocrinology*, *161*, 106950. https://doi.org/10.1016/j.psyneuen.2023.106950

Schiller, B., Brustkern, J., Walker, M., Hamm, A., & Heinrichs, M. (2023). Oxytocin has sex-specific effects on trust and underlying neurophysiological processes. *Psychoneuroendocrinology*, *151*, 106076. https://doi.org/10.1016/j.psyneuen.2023.106076

Schiller, B., Domes, G., & Heinrichs, M. (2020). Oxytocin changes behavior and spatio-temporal brain dynamics underlying inter-group conflict in humans. *European Neuropsychopharmacology*, *31*, 119–130. https://doi.org/10.1016/j.euroneuro.2019.12.109

Schiller, B., Koenig, T., & Heinrichs, M. (2019). Oxytocin modulates the temporal dynamics of resting EEG networks. *Scientific Reports*, *9*(1), 13418. https://doi.org/10.1038/s41598-019-49636-6

Singh, F., Nunag, J., Muldoon, G., Cadenhead, K. S., Pineda, J. A., & Feifel, D. (2016). Effects of intranasal oxytocin on neural processing within a socially relevant neural circuit. *European Neuropsychopharmacology*, *26*(3), 626–630. https://doi.org/10.1016/j.euroneuro.2015.12.026

Soriano, J. R., Daniels, N., Prinsen, J., & Alaerts, K. (2020). Intranasal oxytocin enhances approach-related EEG frontal alpha asymmetry during engagement of direct eye contact. *Brain Communications*, *2*(2), fcaa093. https://doi.org/10.1093/braincomms/fcaa093

Tillman, R., Gordon, I., Naples, A., Rolison, M., Leckman, J. F., Feldman, R., Pelphrey, K. A., & McPartland, J. C. (2019). Oxytocin Enhances the Neural Efficiency of Social Perception. *Frontiers in Human Neuroscience*, *Volume 13-2019*. https://www.frontiersin.org/journals/human-neuroscience/articles/10.3389/fnhum.2019.00071

Tomescu, M. I., Van der Donck, S., Perisanu, E. M., Berceanu, A. I., Alaerts, K., Boets, B., & Carcea, I. (2024). Social functioning predicts individual changes in EEG microstates following intranasal oxytocin administration: A double-blind, cross-over randomized clinical trial. *Psychophysiology*, *61*(8), e14581. https://doi.org/10.1111/psyp.14581

Van der Donck, S., Moerkerke, M., Dlhosova, T., Vettori, S., Dzhelyova, M., Alaerts, K., & Boets, B. (2022). Monitoring the effect of oxytocin on the neural sensitivity to emotional faces via frequency-tagging EEG: A double-blind, cross-over study. *Psychophysiology*, *59*(7), e14026. https://doi.org/10.1111/psyp.14026

Zelenina, M., Kosilo, M., da Cruz, J., Antunes, M., Figueiredo, P., Mehta, M. A., & Prata, D. (2022). Temporal dynamics of intranasal oxytocin in human brain electrophysiology. *Cerebral Cortex*, *32*(14), 3110–3126. https://doi.org/10.1093/cercor/bhab404

Zhang, X., Li, P., Otieno, S. C. S. A., Li, H., & Leppänen, P. H. T. (2021). Oxytocin reduces romantic rejection-induced pain in online speed-dating as revealed by decreased frontal-midline theta oscillations. *Psychoneuroendocrinology*, *133*, 105411. https://doi.org/10.1016/j.psyneuen.2021.105411

Zhuang, Q., Zhu, S., Yang, X., Zhou, X., Xu, X., Chen, Z., Lan, C., Zhao, W., Becker, B., Yao, S., & Kendrick, K. M. (2021). Oxytocin-induced facilitation of learning in a probabilistic task is associated with reduced feedback- and error-related negativity potentials. *Journal of Psychopharmacology*, *35*(1), 40–49. https://doi.org/10.1177/0269881120972347
